# General-purpose large language models can achieve physician-level accuracy in complex medical data extraction

**DOI:** 10.64898/2026.06.06.26354838

**Authors:** Manu Rajeev, Ananthu Narayan

**Affiliations:** Department of Hepatology, Institute of Liver and Biliary Sciences, New Delhi 110 070, India

## Abstract

**Background:** Unstructured data represent about 80% of total electronic health records (EHR) data. Structuring this free text is essential for advancing clinical research, including cohort selection for trials, retrospective studies, and the development of disease registries. While manual chart review (MCR) remains the gold standard for extracting this clinical data, the process is inherently slow, resource-intensive, and susceptible to errors from human fatigue. We evaluated the extraction accuracy, safety, and efficiency of the HeLIX (Hepatology Logic-Integrated Extraction) framework, a Large Language Model (LLM) protocol using Google Gemini 3 Pro, compared to a gold-standard Manual Chart Review (MCR).

**Methods:** A prospective validation study was conducted using 50 high-complexity, simulated hepatology discharge summaries designed to replicate the real-world heterogeneity of EHRs. The HeLIX framework employed a Zero-Shot, Structured Chain-of-Thought (CoT) prompting strategy enforced by a three-layer architecture: Clinical Reasoning Trace, Schema Enforcement, and Evidence Verification. The model extracted 45 distinct clinical variables. Performance was benchmarked against a consensus MCR.

**Results:** Across 2,250 evaluated data points, the model achieved an overall Extraction Accuracy of 99.24% (95% CI: 98.8%–99.5%), with perfect concordance in 35/45 (77.8%) variables. For binary diagnostic variables, the model demonstrated an overall F1-score of 0.98, Recall of 0.99 and substantial inter-rater reliability (Cohen’s κ = 0.97). Hallucinations were exceptionally rare (2/2250; 0.08%). Critical errors affecting clinical management occurred in only 2 instances (<0.1% of total data), both involving etiological misattribution in complex multifactorial diagnoses. The AI workflow was 13.4-fold faster and 95.1% more cost-effective than manual extraction.

**Conclusion:** The HeLIX framework demonstrates physician-level accuracy and reliability in extracting complex hepatology data. It offers a scalable, efficient, and economical alternative to manual chart review. Such frameworks could accelerate clinical research, enabling healthcare systems globally to build comprehensive patient registries for a fraction of the traditional cost.

## INTRODUCTION

Electronic health records (EHRs) store vast quantities of longitudinal patient data, yet a significant proportion remains locked within unstructured narrative text.^1,2^ Converting this free text into structured data is essential for a wide spectrum of healthcare domains, ranging from retrospective studies, development of disease registries, and patient selection for clinical trials to administrative tasks such as billing, auditing, and quality improvement. Currently, the gold standard for generating such high-quality datasets is through manual chart review (MCR).^3–5^ However, this process is resource-intensive, slow, and prone to errors due to human fatigue.^6–8^

Technological approaches to this problem have centered on the application of Natural Language Processing (NLP).^9,10^ NLP is a branch of artificial intelligence (AI) that focuses on the interaction between computers and human language to automatically analyze and derive meaning from complex text.^11^ NLP techniques have advanced significantly in the recent years, evolving from rigid rule-based systems to transformer-based Large Language Models (LLMs). Modern LLMs have been shown to demonstrate efficacy in identifying clearly stated diagnoses, medications, or procedural entities from short-text segments.^12–14^ These capabilities have been further enhanced by recent advancements in prompt engineering, which have refined the ability of these models to reason and extract medical data.^15^

Despite these advances, current LLM applications have been reported to have certain limitations. They often struggle when required to extract data from complex clinical narratives that require judgement rather than mere retrieval.^16–20^ This is particularly evident in complex sub-specialties such as hepatology and gastroenterology.^10,18,21–23^ Discharge summaries in these fields contain complicated clinical parameters, non-standard abbreviations and disjointed timelines that render simple keyword searches inefficient.^24^ Another limitation of LLMs is its potential to “hallucinate” (the generation of plausible but factually incorrect information). Clinicians remain hesitant to rely on automated extraction due to this risk of data fabrication, which could compromise the integrity of research data.^25,26^

To address these limitations, we developed and validated the Hepatology Logic-Integrated Extraction (HeLIX) protocol. This study assessed the protocol’s performance using the commercial LLM Google Gemini 3 Pro on a dataset deliberately injected with linguistic noise and clinical ambiguity to simulate real-world hepatology discharge summaries. This novel framework utilized a “Three-Layer Reasoning Architecture” comprising Clinical Reasoning, Schema Enforcement, and Evidence Verification. We aimed to demonstrate that constraining the model with an explicit chain-of-thought reasoning, structured outputs and evidence citation achieved extraction accuracy comparable to the gold-standard Manual Chart Review (MCR).

## METHODS

### Study Design and Setting

We conducted a single-center, prospective validation study to evaluate the extraction accuracy, safety, and operational efficiency of an LLM-based framework in data extraction. We developed a structured prompting framework, termed HeLIX (Hepatology Logic-Integrated Extraction), to standardize the interaction with the Large Language Model (LLM). The study compared the performance of the HeLIX framework against a Manual Chart Review (MCR) in extracting 45 complex clinical variables from 50 unstructured hepatology discharge summaries.

### Dataset Generation

To circumvent privacy limitations associated with Protected Health Information (PHI) and to ensure ground-truth control, we generated a cohort of 50 high-complexity, simulated discharge summaries **(Supplementary Material S1)**. Unlike standard training datasets, these summaries were designed to replicate the heterogeneity of real-world electronic health records (EHRs). The cohort covered a diverse spectrum of hepatobiliary etiologies ranging from compensated cirrhosis to hyperacute liver failure and hepatocellular carcinoma (HCC). A total of 45 distinct clinical variables across five domains were defined for extraction from each summary **(Figure 1A-B)**.

**Figure 1.**
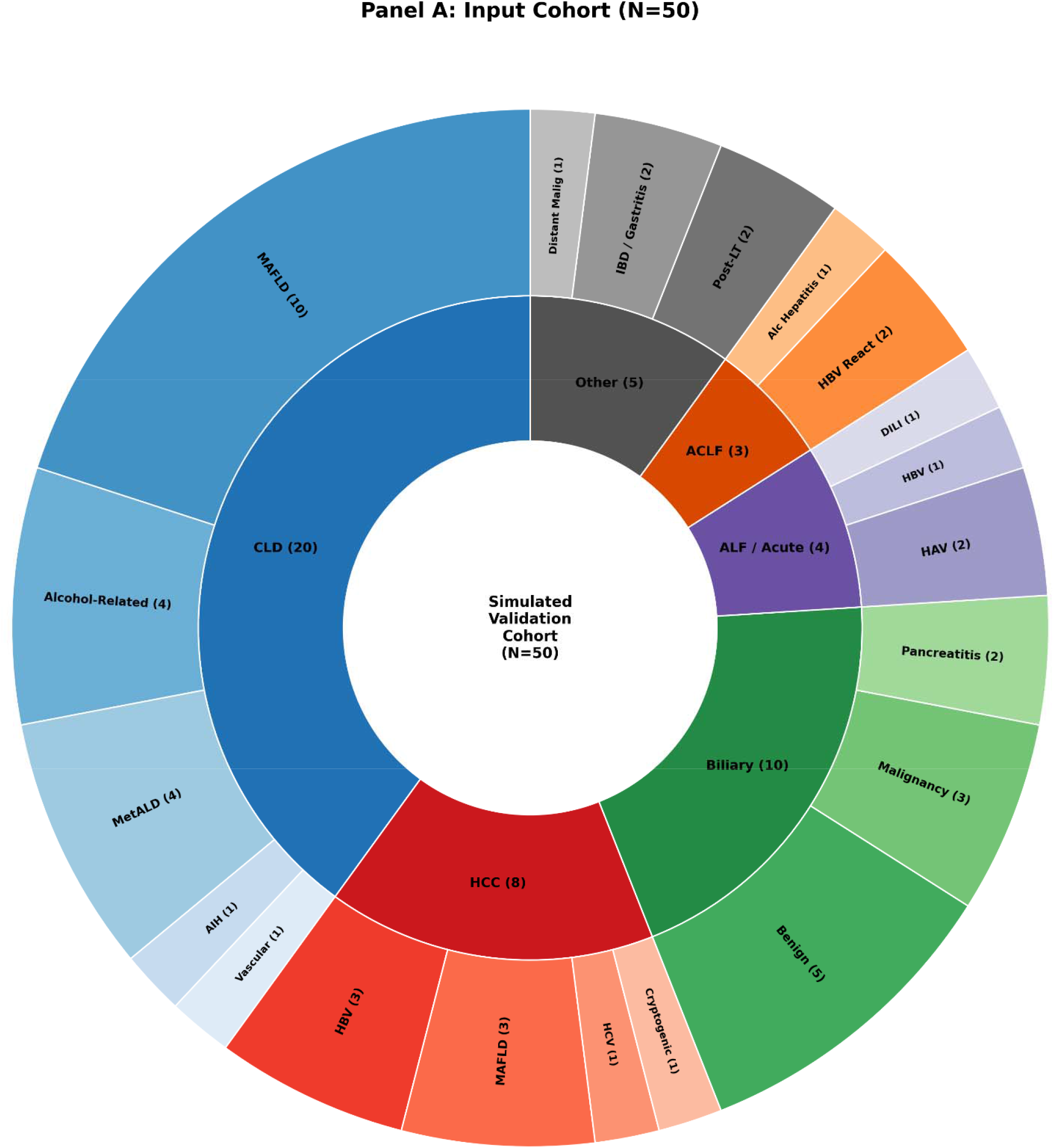
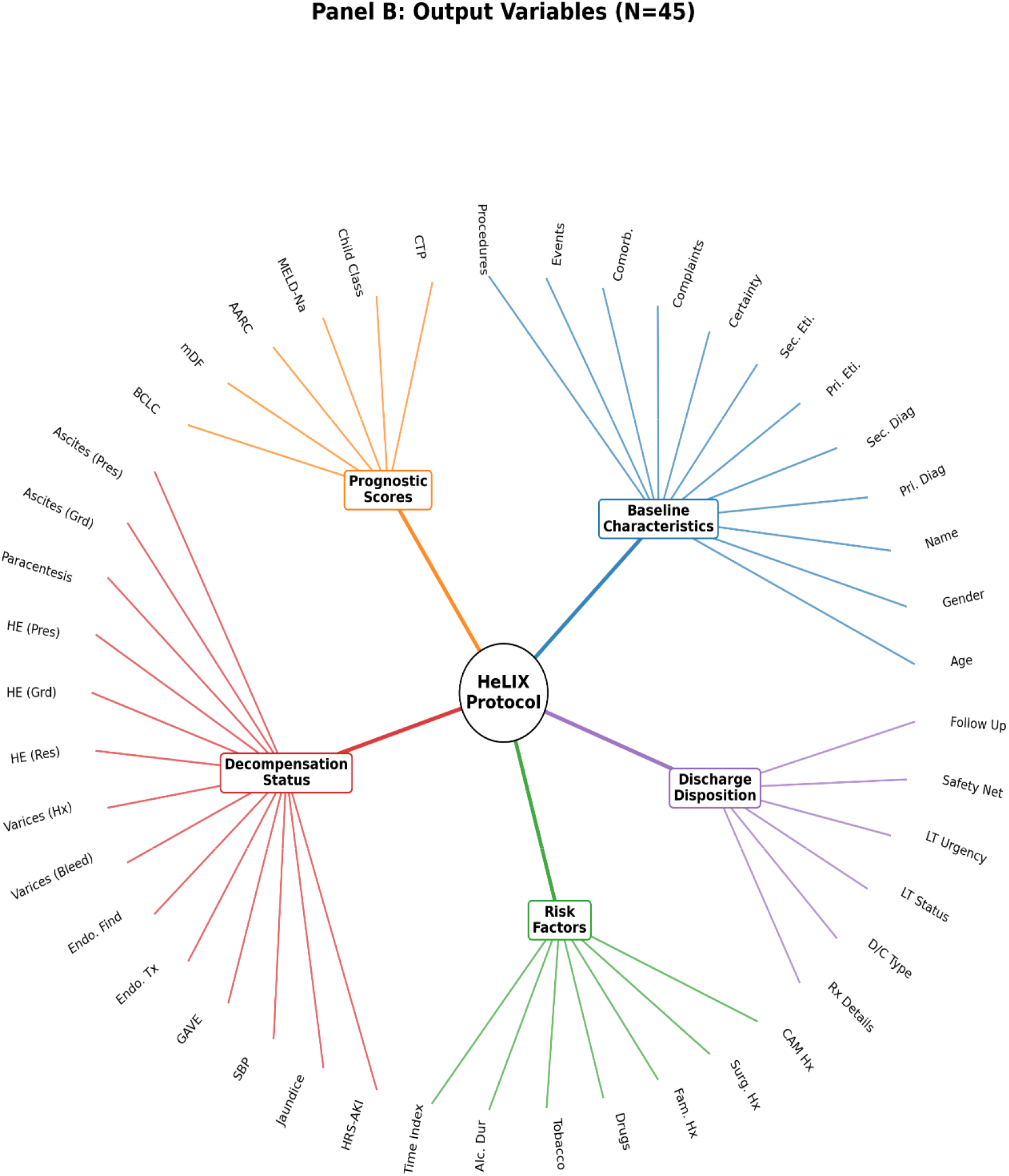
Distribution of Clinical Etiologies and the Panel of 45 Variables Extracted Across the Simulated Validation Cohort (N=50). **(A)** Nest Pie Chart illustrating the disease spectrum of the simulated validation cohort (N=50), stratified by etiology (inner ring) and sub-etiology (outer ring). **(B)** Dendrogram illustrating the 45 clinical variables extracted by the HeLIX protocol. AARC: APASL ACLF Research Consortium; ACLF: Acute-on-Chronic Liver Failure; AIH: Autoimmune Hepatitis; AKI: Acute Kidney Injury; ALF: Acute Liver Failure; BCLC: Barcelona Clinic Liver Cancer; CAM: Complementary and Alternative Medicine; CLD: Chronic Liver Disease; CTP: Child-Turcotte-Pugh; DILI: Drug-Induced Liver Injury; GAVE: Gastric Antral Vascular Ectasia; HAV: Hepatitis A Virus; HBV: Hepatitis B Virus; HCC: Hepatocellular Carcinoma; HCV: Hepatitis C Virus; HE: Hepatic Encephalopathy; HRS: Hepatorenal Syndrome; LT: Liver Transplant; MAFLD: Metabolic Dysfunction–Associated Fatty Liver Disease; mDF: Maddrey Discriminant Function; MELD-Na: Model for End-Stage Liver Disease-Sodium; MetALD: Metabolic Alcohol-Associated Liver Disease.

To evaluate the model, a “Noise Injection Protocol” was applied to the dataset, introducing three layers of complexity to simulate real-world discharge summaries. First, **Linguistic Noise** was introduced involving intentional typographical errors, non-standard abbreviations (e.g., “HE” for Hepatic Encephalopathy, “EVL” for Banding), non-standard dosing (1-1-1-1) and inconsistent date formats. Drugs were documented using a mix of generic and regional brand names (e.g., Rifaximin vs. Rifagut). Second, **semantic complexity** was introduced wherein data points were deliberately dispersed across non-contiguous sections of the summary to test the model’s context window and long-context retrieval capabilities. This included documenting ascites solely within a procedural note (e.g., “200cc fluid removed”) while the physical exam noted a “soft abdomen; tympanic note with no free fluid”, or burying etiology-defining investigations (e.g., “HBsAg Reactive”) in notes far from the primary diagnosis list. Third, **clinical ambiguity** was introduced requiring clinical judgement over simple keyword extraction. This included differentiating “trace pedal edema” from pathological ascites or distinguishing Hepatorenal Syndrome (HRS-AKI) from Prerenal Azotemia based on documented response to volume expansion (e.g. creatinine improvement post-albumin). The model was also required to resolve conflicting information, such as prioritizing objective biomarkers (e.g., elevated PEth levels) over subjective patient reports of alcohol abstinence.

### The HeLIX Protocol

The HeLIX Protocol was executed using Google Gemini 3 Pro (Google DeepMind, Mountain View, CA; February 2026 Build), deployed via the Google AI Studio environment (https://aistudio.google.com/). The protocol leveraged the model’s expanded context window of 1 million tokens, thus overcoming the truncation errors of previous studies.^27–29^ The Temperature was set to 0.7, as validated in previous studies.^26,30,31^ Nucleus Sampling (Top-P) was maintained at 0.95.

To accommodate the high variability of hepatology discharge summaries, we employed a Zero-Shot Prompting strategy **(Supplementary Material S2)**. This eliminates the need for ‘Few-Shot’ examples or case-specific fine-tuning, enabling immediate deployment by end-users with limited technical expertise.^32–36^ Additionally, we integrated a Structured Chain-of-Thought (CoT) reasoning. This not only improves extraction accuracy but also allows clinicians to audit the model’s decision-making process using clinical logic rather than technical code.^15,37^ The system prompt underwent a design phase using a separate set of 10 simulated discharge summaries. This phase optimized the JSON schema instructions and CoT triggers. These 10 cases were excluded from the final validation cohort of 50 summaries to prevent bias.

A Three-Layer Reasoning Architecture was devised

#### Layer 1 - Clinical Reasoning

By employing a “Clinical Reasoning Trace”, a form of Chain-of-Thought (CoT) prompting, the protocol created a reasoning step where the model resolved temporal conflicts (e.g., converting relative dates to calendar years) and clinical ambiguities before committing to a final output. Heuristic and CoT prompting have been demonstrated to outperform simple zero-shot prompts in clinical NLP tasks.^15,27,31,38^ Without an enforced reasoning structure, general-purpose models cannot leverage their full inferential capabilities.^34,35^

#### Layer 2 – Schema Enforcement

The second layer, Schema Enforcement, restricted the extracted output into a JSON (JavaScript Object Notation) schema covering 45 variables. By constraining the model to produce data in rigid formats, LLMs have been shown to better organize information, reduce hallucinations, and facilitate downstream data processing.^27,39–41^ To prevent hallucinations, the model was explicitly instructed to output “N/A” if a variable was not textually present. Additionally, the Schema Enforcement layer was used to leverage the parametric knowledge of the LLM to generate uniform outputs for prescriptions. This was achieved by adding an instructional constraint within the JSON schema {“Generic_Name”: “String (Convert Brand to Generic)”} to convert diverse proprietary drug names into their International Nonproprietary Names (INN). This obviated the need for the complex external lookup pipelines (such as MedXN20 and RxNorm mapping) or specific fine-tuning traditionally used in rule-based NLP.^41^

#### Layer 3 – Evidence Verification

The third layer, Evidence Verification, addressed the inherent opacity of deep learning models.^2^ The model was required to extract the specific verbatim text (substring) that supported its decision, allowing reviewers to verify the source of any data point. This requirement has been shown to contribute to the mitigation of hallucinations.^17,26,27,42,43^ Additionally, the model was mandated to expose the Clinical Reasoning Trace (Layer 1) to accelerate auditing and error analyses.

The reference standard (Ground Truth) was established through a manual chart review of all 50 summaries by two independent physicians with hepatology experience (MR, AN), who remained blinded to the model outputs. To ensure independence, human extraction was done prior to the AI analysis.

### Outcome Measures

The primary endpoints evaluated were the extraction performance (Sensitivity, Specificity, Precision, Overall Accuracy and F1-scores) and reliability (Cohen’s Kappa). Overall accuracy was defined as the percentage of total data points where the AI output perfectly matched the manual chart review. For each binary variable, true positives, true negatives, false positives, and false negatives were defined by direct comparison of AI-extracted values against manual chart review (Human = Yes/No), and standard confusion-matrix–based metrics were calculated. The F1-score was calculated to assess the harmonic mean of precision (positive predictive value) and recall (sensitivity). Inter-rater reliability was assessed using Cohen’s Kappa (κ) to quantify agreement beyond chance.

Secondary endpoints focused on error characterization and model efficiency. Discrepancies were manually audited and classified using an **Error Taxonomy**: Type A (Omission/False Negative), Type B (Hallucination/False Positive), Type C (Value Error), Type D (Attribution), and Type E (Temporal). Errors were further stratified by clinical severity: Level 1 (Minor) for formatting or semantic variations without clinical consequence; Level 2 (Moderate) for significant errors that preserved the primary diagnosis and management; and Level 3 (Critical) for failures that would fundamentally alter patient diagnosis, staging, prognosis, or management. Additionally, we performed a qualitative error analysis for each error to identify the specific linguistic or reasoning deficits.

Finally, we conducted an efficiency analysis. Time-to-extraction was measured by comparing the median time-to-extraction for the AI workflow versus manual chart review. A commercial feasibility model was constructed using Google Vertex AI pricing (February 2026 rates: $2.00 USD per 1 million input tokens and $12.00 USD per 1 million output tokens) to estimate deployment costs in a real-world hospital setting, benchmarked against institutional clinician billing rates.

### Statistical Analysis

Categorical variables were reported as counts and percentages. Continuous variables were reported as medians with Interquartile Ranges (IQR). Confidence Intervals (95% CI) for accuracy were calculated using the Wilson Score Interval method. For binary variables, any “N/A” outputs from the model (indicating absence of documentation) were converted to “No” (negative) prior to analysis of confusion matrix. All analyses were performed using Microsoft Excel (v.16.0) and Python (Pandas library).

## RESULTS

### Accuracy of AI-Based Data Extraction

Across 2,250 datapoints (45 variables across 50 hepatology discharge summaries), the LLM achieved an overall extraction accuracy of 99.24% (95% CI: 98.8% – 99.5%) when compared against manual chart review **(Figure 2)**.

**Figure 2.**
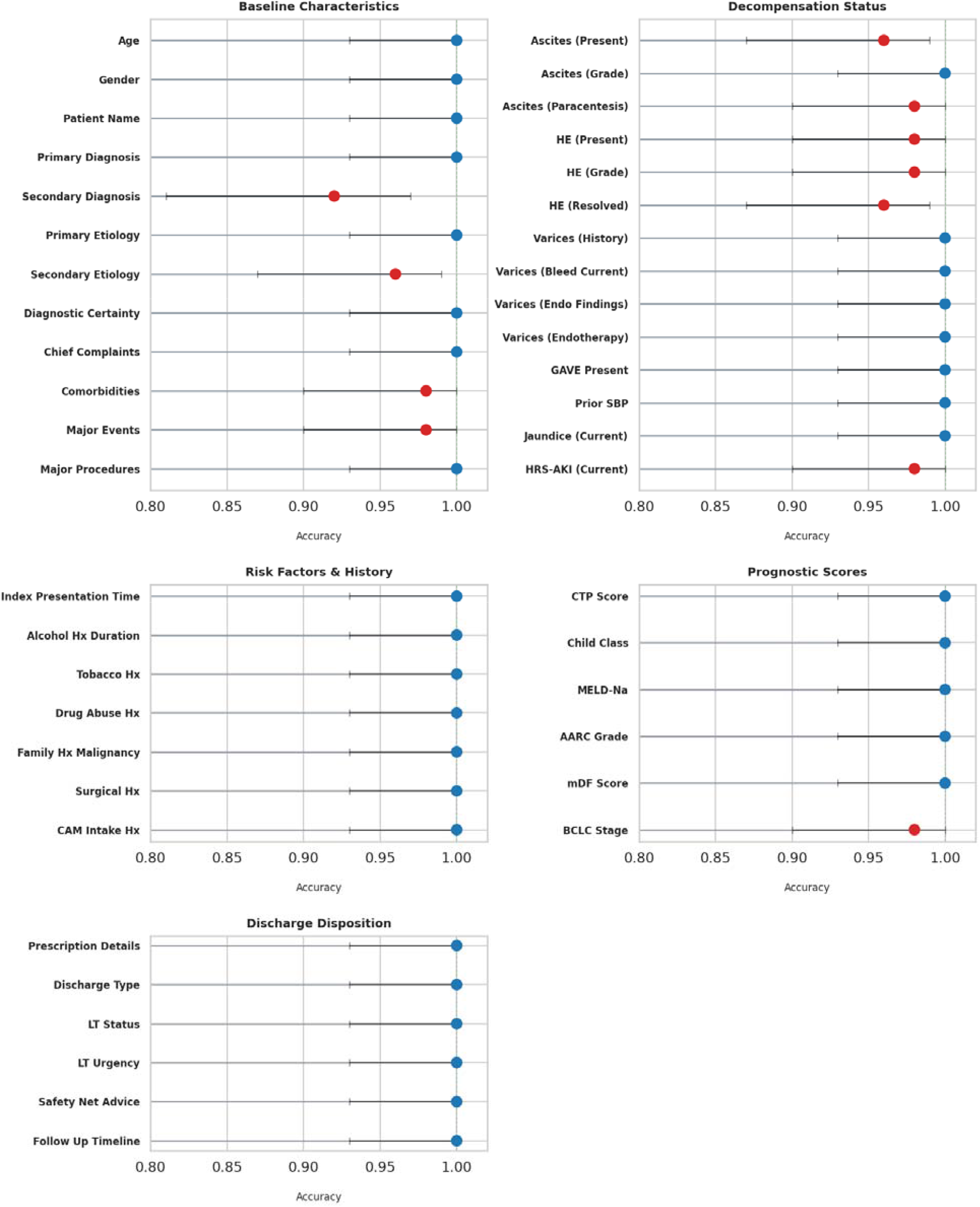
Diagnostic Accuracy and 95% Confidence Intervals for Automated Extraction Across 45 Clinical Variables (n=2,250)

The model achieved 100% accuracy across 35 of the 45 variables (77.8%), capturing all demographic identifiers, composite prognostic scores (CTP, MELD-Na), and primary discharge diagnoses. Errors were noted in a subset of 10 variables where accuracy ranged from 92.0% to 98.0%. The most numerous errors were observed in Secondary Diagnoses (92.0%) and HE Resolution Status (96.0%), reflecting the difficulty in extracting semantically complex fields.

Additionally, the evaluation revealed 6 instances of AI-identified omissions (Type F), where the model correctly identified valid clinical findings (e.g., buried comorbidities) that were not documented during human chart review. These cases were documented separately (Type F Errors) and were not included in the error counts. Consequently, the reference standard was updated, and these data points were reclassified as True Positives.

### Diagnostic Performance on Binary Clinical Variables

Across 15 predefined binary clinical variables, the LLM demonstrated consistently high diagnostic performance, with F1-scores ranging from 0.91 to 1.00 (Overall 0.98) **(Table 1)**.

**Table 1.** Confusion Matrix and Performance Metrics for 15 Binary Clinical Variables. Balanced Accuracy is calculated as (Sensitivity + Specificity) / 2. CAM: Complementary and Alternative Medicine; GAVE: Gastric Antral Vascular Ectasia; HE: Hepatic Encephalopathy; HRS-AKI: Hepatorenal Syndrome-Acute Kidney Injury; SBP: Spontaneous Bacterial Peritonitis.

| Clinical Variable | TP | TN | FP | FN | Precision | Sensitivity | Specificity | F1-Score | Balanced Accuracy |
| --- | --- | --- | --- | --- | --- | --- | --- | --- | --- |
| Ascites - Present in Current Admission | 25 | 22 | 3 | 0 | 0.89 | 1.00 | 0.88 | 0.94 | 0.94 |
| Ascites - Paracentesis Required | 6 | 43 | 0 | 1 | 1.00 | 0.86 | 1.00 | 0.92 | 0.93 |
| HE - Present in Current Admission | 5 | 44 | 1 | 0 | 0.83 | 1.00 | 0.98 | 0.91 | 0.99 |
| HE - Resolved in Current Admission | 10 | 38 | 2 | 0 | 0.83 | 1.00 | 0.95 | 0.91 | 0.98 |
| Variceal Status - History of Bleed | 14 | 36 | 0 | 0 | 1.00 | 1.00 | 1.00 | 1.00 | 1.00 |
| Variceal Status - Bleed in Current Admission | 6 | 44 | 0 | 0 | 1.00 | 1.00 | 1.00 | 1.00 | 1.00 |
| Endoscopy - GAVE Present | 7 | 43 | 0 | 0 | 1.00 | 1.00 | 1.00 | 1.00 | 1.00 |
| Prior SBP | 2 | 48 | 0 | 0 | 1.00 | 1.00 | 1.00 | 1.00 | 1.00 |
| Jaundice in Current Admission | 24 | 26 | 0 | 0 | 1.00 | 1.00 | 1.00 | 1.00 | 1.00 |
| HRS-AKI in Current Admission | 7 | 42 | 1 | 0 | 0.88 | 1.00 | 0.98 | 0.93 | 0.99 |
| Tobacco History | 7 | 43 | 0 | 0 | 1.00 | 1.00 | 1.00 | 1.00 | 1.00 |
| Drug Abuse History | 2 | 48 | 0 | 0 | 1.00 | 1.00 | 1.00 | 1.00 | 1.00 |
| Family History of Malignancy | 2 | 48 | 0 | 0 | 1.00 | 1.00 | 1.00 | 1.00 | 1.00 |
| CAM Intake History | 4 | 46 | 0 | 0 | 1.00 | 1.00 | 1.00 | 1.00 | 1.00 |
| Safety Net Advice | 40 | 10 | 0 | 0 | 1.00 | 1.00 | 1.00 | 1.00 | 1.00 |
| **Overall (Pooled Analysis)** | **161** | **581** | **7** | **1** | **0.96** | **0.99** | **0.99** | **0.98** | **0.99** |

The model demonstrated near-perfect Recall (Sensitivity) across all variables (mean Recall: 0.99), and notably, a high Specificity of 0.99. This resulted in a Mean Balanced Accuracy [(Sensitivity + Specificity)/2] of 0.991. Precision (Positive Predictive Value) was similarly high at 0.96 **(Figure 3)**.

**Figure 3.**
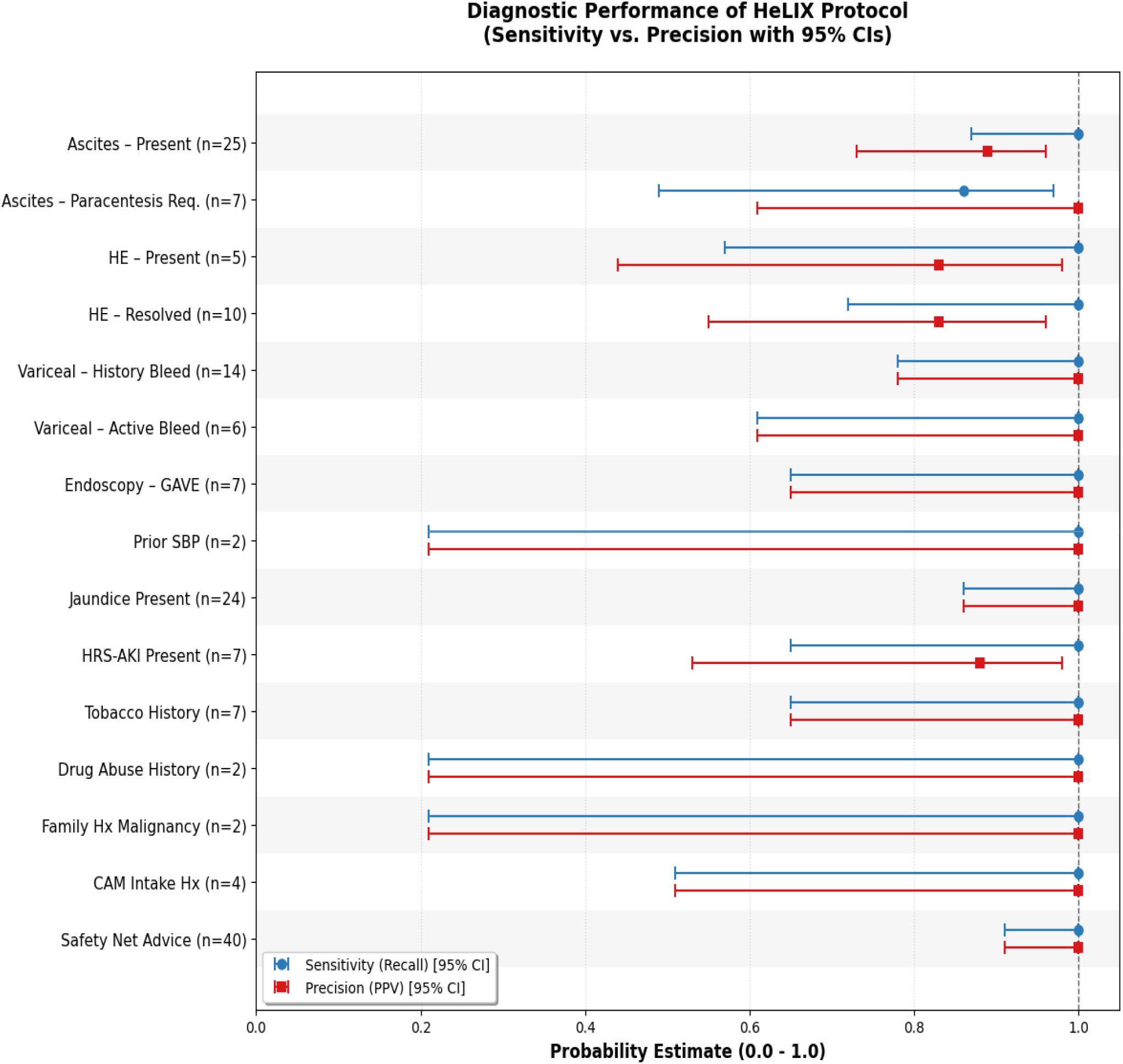
Diagnostic performance (Sensitivity and Precision) with 95% Confidence Intervals. Variables are annotated with the total number of positive cases (n).

Minor reductions in Precision (0.83 – 0.89) were observed in complex narratives involving Hepatic Encephalopathy and Ascites, where the model displayed a tendency towards over-diagnose (False Positives) rather than omit. When pooled across variables, Cohen’s κ was 0.97, indicating almost perfect agreement between AI-based extraction and physician manual chart review beyond chance.

### Error Taxonomy and Clinical Severity

Among 2,250 evaluated data points, a total of 17 mismatches (0.76%) were identified **(Table 2)**.

**Table 2.** Failure Mode Analysis and Clinical Severity of Errors in in AI Extractions Against the Manual Chart Review (N= 17)

| Classification Category | Description | Frequency (n) | Proportion (%) |
| --- | --- | --- | --- |
| I. Error Taxonomy |  |  |  |
| Type A | Omission (False Negative) | 5 | 29.4% |
| Type B | Hallucination (False Positive) | 2 | 11.8% |
| Type C | Value Mismatch | 0 | 0.0% |
| Type D | Attribution Error | 9 | 52.9% |
| Type E | Temporal Error | 1 | 5.9% |
|  | Total Errors | 17 | 100.0% |
| II. Clinical Severity |  |  |  |
| Level 1 | Minor: Insignificant variations; no clinical impact | 3 | 17.6% |
| Level 2 | Moderate: Significant; preserved primary diagnosis | 12 | 70.6% |
| Level 3 | Critical: Altered patient diagnosis or management | 2 | 11.8% |
|  | Total Errors | 17 | 100.0% |

Stratification by clinical severity showed that the majority of errors were Level 1 (Minor, 17.6%) or Level 2 (Moderate, 70.6%), and did not compromise the primary diagnosis. Level 3 (Critical) errors, defined as failures with potential to alter clinical management, were rare (n=2, 11.8%). In both instances of critical errors, the model incorrectly identified incidental findings (Cholelithiasis) or historical exposures (CAM) as the secondary etiological factor of disease, along with the primary etiology (Gallbladder Carcinoma and HBV, respectively) **(Table 3)**.

**Table 3.**
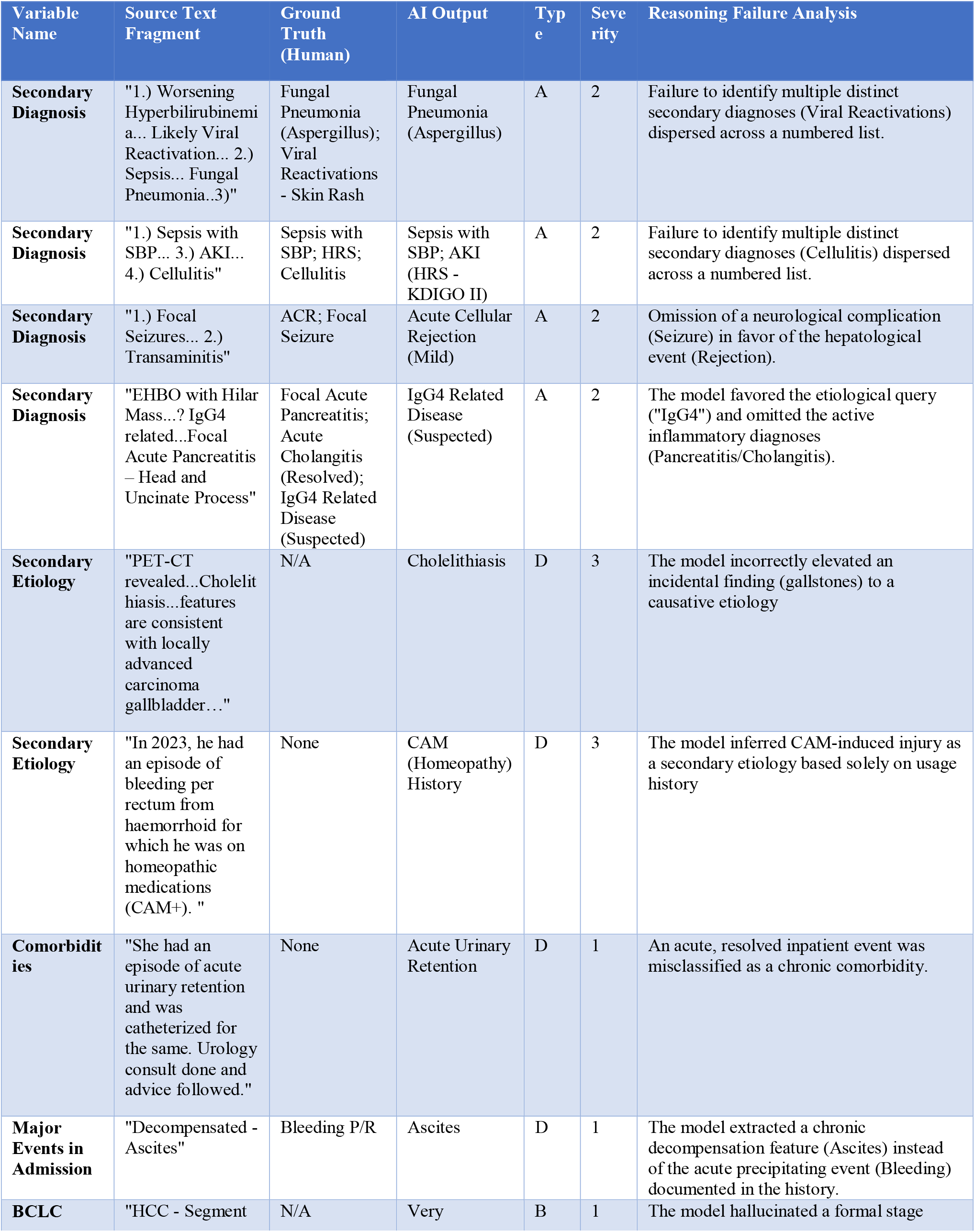
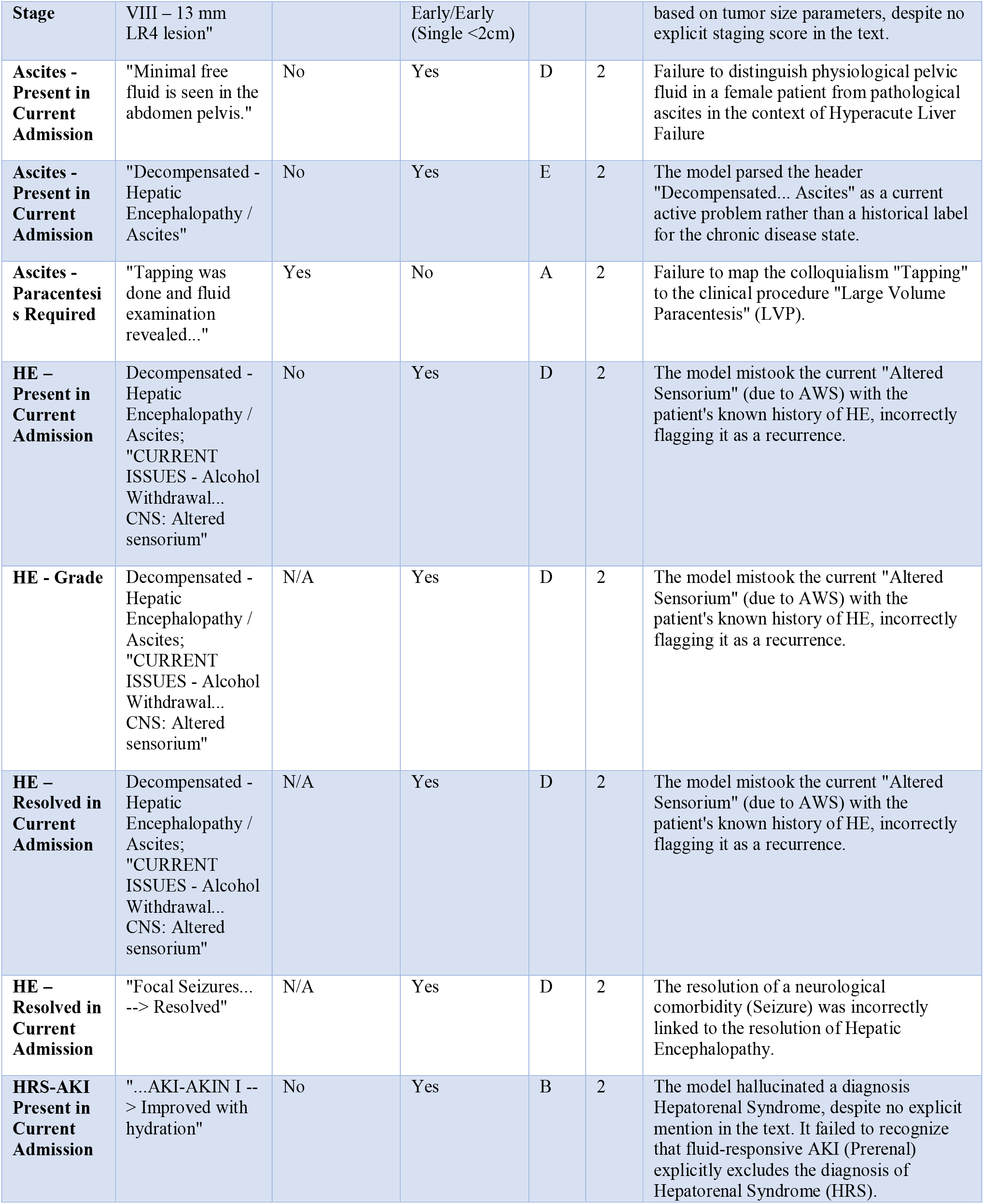
Qualitative Error Analysis of Errors in AI Extractions Against the Manual Chart Review (N= 17) Error Types: Type A: Omission; Type B: Hallucination; Type C: Value Error; Type D: Attribution; Type E: Temporal. Severity: Level 1: Minor; Level 2: Moderate; Level 3: Critical. ACR: Acute Cellular Rejection; AKI: Acute Kidney Injury; AWS: Alcohol Withdrawal Syndrome; BCLC: Barcelona Clinic Liver Cancer; CAM: Complementary and Alternative Medicine; CNS: Central Nervous System; HRS: Hepatorenal Syndrome; SBP: Spontaneous Bacterial Peritonitis.

When classified by error mechanism, the predominant errors were Attributional (Type D, 52.9%) and Omission (Type A, 29.4%). Notably, Hallucinations (Type B) that involve the generation of non-existent data were exceptionally rare, occurring in two instances (11.8% of errors; 0.08% of total data points).

### Efficiency and Resource Utilization

While manual expert extraction required a median of 10.5 minutes (632.5 seconds) per summary (IQR: 498–740 seconds), the AI workflow reduced this to a median of 46.5 seconds (IQR: 38–56 seconds), corresponding to a 92.5% reduction in extraction time and a 13.4-fold increase in speed **(Table 4)**.

**Table 4.** Comparative Analysis of Temporal Efficiency, Economic Cost, and Throughput between Manual Physician Review and Automated AI Extraction (N=50) AI: Artificial Intelligence; IQR: Interquartile Range (Q1–Q3); USD: United States Dollar.

| Metric | Manual Chart Review | AI Model |
| --- | --- | --- |
| Temporal Efficiency |  |  |
| Median Processing Time (IQR) | 10.54 min (8.3 – 12.3) | 0.78 min (0.63 – 0.93) |
| Total Cohort Time (N=50) | 8.74 hours | 0.65 hours |
| Economic Analysis (USD) |  |  |
| Unit Cost per Summary | \$0.76 | \$0.04 |
| Total Cohort Cost (N=50) | \$38.12 | \$1.88 |
| Tokens Consumed |  |  |
| Total Token Load Processed | — | 308,988 Tokens |
| Mean Tokens per Summary | — | 6,180 Tokens |

Economic modeling estimated the total cost of manual extraction as □3,495.22 ($38.12 USD), compared with an AI computational cost of □172.50 ($1.88 USD). This represents a 95.1% reduction in resource expenditure.

## DISCUSSION

In this validation study, the Hepatology Logic-Integrated Extraction (HeLIX) protocol achieved an overall accuracy of 99.24% while increasing extraction velocity by 13-fold and reducing costs by 95.1% compared to manual chart review. These findings are particularly significant given the inherent complexity of hepatology documentation. The datasets contained disjointed timelines and semantic ambiguity, often requiring the distinction between active and historical decompensation events, or the differentiation between chronic bleeding secondary to Gastric Antral Vascular Ectasia (GAVE) and acute variceal bleeding. This demonstrated that modern LLMs are capable of parsing high-complexity clinical text. Furthermore, the protocol successfully extracted 45 distinct clinical variables per summary, for a total of 2,250 data points. This represents a marked increase over standard protocols that used LLMs in recent literature, which were often limited to 10–15 isolated variables.^13,14,44^ This showed that the model’s long context window effectively handled dense, longitudinal data. Consequently, this enables the framework to offer a scalable method for generating comprehensive disease registries.

The overall accuracy of HeLIX is comparable with recent literature. Ntinopoulos et al.^14^ reported accuracies exceeding 98% using advanced models like GPT-4 and Claude 3 Opus on simulated medical notes. Similarly, Far et al.^44^ achieved 99.3% accuracy in extracting Vibration Controlled Transient Elastography (VCTE) stiffness values using GPT-4o. These findings demonstrate a marked improvement over benchmarks from older (now deprecated) models like GPT-3.5, as reported by Ge et al.^12^ for hepatocellular carcinoma imaging (89%) and Huang et al.^13^ for lung cancer pathology (89%).

While Manual Chart Review (MCR) remains the gold standard for registry generation, it is prone to cognitive fatigue, which increases error rates during repetitive tasks.^5,6^ Furthermore, it is highly constrained by costs and resource demands.^5–8,11,45^ These barriers create a systemic disparity in research and health informatics: resource-limited nations are often unable to establish high-quality registries due to the expense of physician-led chart review.^46–52^ The use of LLMs offer a scalable economic alternative, rendering the development of large-scale registries feasible regardless of geographical or financial constraints.

The HeLIX protocol demonstrated a 95.1% reduction in cost compared to manual chart review (MCR). This is consistent with findings in recent literature, including Far et al.^44^ ($0.012–0.014 per note) and Choi et al.^53^ ($909.30 for manual review vs. $95.40 for LLM extraction). Automated extraction has been demonstrated to be profoundly more economical than clinical labor, and could fundamentally alter the economics of data collection.^7,54,55^ By lowering the barrier to entry, automated extraction facilitates the *de novo* creation of large-scale registries. This enables resource-limited nations to generate robust patient registries without the need for expensive GPU infrastructure.^56–58^

Our findings also indicate that a general-purpose LLM, when constrained by a reasoning architecture and structured output, can achieve high accuracy. The debate concerning the optimal strategy for adapting LLMs to the medical domain currently centers on the choice between prompt engineering and fine-tuning.^59,60^ The above results challenge the prevailing assumption that fine-tuning is necessary for high-performance extraction.^61^ The protocol’s high accuracy and ability to minimize hallucinations eliminate the need for extensive computational resources and labeled training datasets required by fine-tuning, thereby addressing the critical limitations of current NLP methodologies.^1,13,40^ In addition to its high cost, fine-tuning inherently yields a static knowledge base. Consequently, updating this knowledge base (e.g., when a new drug enters the market) requires retraining and further computational costs. In contrast, the prompt-based approach allows for immediate knowledge updates by simply editing the prompt, offering greater adaptability.

This study has certain limitations. The primary limitation is the use of simulated discharge summaries. NLP models may perform differently on real-world data, which often contain higher levels of noise and ambiguity.^17^ Our dataset lacked Optical Character Recognition (OCR) artifacts and dictation errors that are common in real-world EHRs. Secondly, the study is confined to hepatology. Domain adaptation remains a challenge, and the generalizability of the HeLIX protocol to other medical specialties requires further investigation. However, we anticipate that the underlying prompt engineering techniques build a transferable framework likely to produce similar results in other specialties. Thirdly, the use of a cloud-based model necessitates strict de-identification of patient data to comply with privacy regulations.^62,63^ Fourthly, a reliance on a commercial API introduces the risk of model drift.^17^ Finally, despite the visible logical trace provided by the CoT and Evidence Verification layers, the underlying neural parameters remain opaque, a limitation inherent to current deep learning models.^64^

Our findings demonstrate that general-purpose LLMs, when constrained by strategic prompting, can extract medical data with high accuracy, obviating the need for resource-intensive manual chart review. Such frameworks could accelerate clinical research, enabling healthcare systems globally to build comprehensive patient registries for a fraction of the traditional cost.

## Data Availability

All data produced in the present work are contained in the manuscript.

## REFERENCES

1. Li I, Pan J, Goldwasser J, et al. Neural Natural Language Processing for unstructured data in electronic health records: A review. Comput Sci Rev. 2022;46:100511. doi:10.1016/j.cosrev.2022.100511

2. Xiao C, Choi E, Sun J. Opportunities and challenges in developing deep learning models using electronic health records data: a systematic review. J Am Med Inform Assoc JAMIA. 2018;25(10):1419–1428. doi:10.1093/jamia/ocy068

3. Kreimeyer K, Foster M, Pandey A, et al. Natural language processing systems for capturing and standardizing unstructured clinical information: A systematic review. J Biomed Inform. 2017;73:14–29. doi:10.1016/j.jbi.2017.07.012

4. Iroju OG, Olaleke JO. A Systematic Review of Natural Language Processing in Healthcare. Int J Inf Technol Comput Sci. 7(8):44.

5. Melton GB, Hripcsak G. Automated Detection of Adverse Events Using Natural Language Processing of Discharge Summaries. J Am Med Inform Assoc. 2005;12(4):448–457. doi:10.1197/jamia.M1794

6. Murff HJ, Patel VL, Hripcsak G, Bates DW. Detecting adverse events for patient safety research: a review of current methodologies. J Biomed Inform. 2003;36(1):131–143. doi:10.1016/j.jbi.2003.08.003

7. Agatstein K. Chart Review Is Dead; Long Live Chart Review: How Artificial Intelligence Will Make Human Review of Medical Records Obsolete, One Day. Popul Health Manag. 2023;26(6):438–440. doi:10.1089/pop.2023.0227

8. Wu ST, Sohn S, Ravikumar KE, et al. Automated chart review for asthma cohort identification using natural language processing: an exploratory study. Ann Allergy Asthma Immunol. 2013;111(5):364–369. doi:10.1016/j.anai.2013.07.022

9. Hou JK, Imler TD, Imperiale TF. Current and Future Applications of Natural Language Processing in the Field of Digestive Diseases. Clin Gastroenterol Hepatol. 2014;12(8):1257–1261. doi:10.1016/j.cgh.2014.05.013

10. Omar M, Nassar S, SharIf K, Glicksberg BS, Nadkarni GN, Klang E. Emerging applications of NLP and large language models in gastroenterology and hepatology: a systematic review. Front Med. 2025;11. doi:10.3389/fmed.2024.1512824

11. Pons E, Braun LMM, Hunink MGM, Kors JA. Natural Language Processing in Radiology: A Systematic Review. Radiology. 2016;279(2):329–343. doi:10.1148/radiol.16142770

12. Ge J, Li M, Delk MB, Lai JC. A comparison of large language model versus manual chart review for extraction of data elements from the electronic health record. medRxiv. Preprint posted online September 4, 2023:2023.08.31.23294924. doi:10.1101/2023.08.31.23294924

13. Huang J, Yang DM, Rong R, et al. A critical assessment of using ChatGPT for extracting structured data from clinical notes. Npj Digit Med. 2024;7(1):106. doi:10.1038/s41746-024-01079-8

14. Ntinopoulos V, Biefer HRC, Tudorache I, et al. Large language models for data extraction from unstructured and semi-structured electronic health records: a multiple model performance evaluation. BMJ Health Care Inform. 2025;32(1). doi:10.1136/bmjhci-2024-101139

15. Sivarajkumar S, Kelley M, Samolyk-Mazzanti A, Visweswaran S, Wang Y. An Empirical Evaluation of Prompting Strategies for Large Language Models in Zero-Shot Clinical Natural Language Processing: Algorithm Development and Validation Study. JMIR Med Inform. 2024;12:e55318. doi:10.2196/55318

16. Rodrigues T, Teixeira Lopes C. Harnessing Large Language Models for Clinical Information Extraction: A Systematic Literature Review. ACM Trans Comput Healthc. 2025;6(4):47:1–47:35. doi:10.1145/3744660

17. Hager P, Jungmann F, Holland R, et al. Evaluation and mitigation of the limitations of large language models in clinical decision-making. Nat Med. 2024;30(9):2613–2622. doi:10.1038/s41591-024-03097-1

18. Sheikhalishahi S, Miotto R, Dudley JT, Lavelli A, Rinaldi F, Osmani V. Natural Language Processing of Clinical Notes on Chronic Diseases: Systematic Review. JMIR Med Inform. 2019;7(2):e12239. doi:10.2196/12239

19. Chapman WW, Nadkarni PM, Hirschman L, D’Avolio LW, Savova GK, Uzuner O. Overcoming barriers to NLP for clinical text: the role of shared tasks and the need for additional creative solutions. J Am Med Inform Assoc. 2011;18(5):540–543. doi:10.1136/amiajnl-2011-000465

20. Kimia AA, Savova G, Landschaft A, Harper MB. An Introduction to Natural Language Processing: How You Can Get More From Those Electronic Notes You Are Generating. Pediatr Emerg Care. 2015;31(7):536. doi:10.1097/PEC.0000000000000484

21. Spasic I, Nenadic G. Clinical Text Data in Machine Learning: Systematic Review. JMIR Med Inform. 2020;8(3):e17984. doi:10.2196/17984

22. Stammers M, Ramgopal B, Owusu Nimako A, et al. A foundation systematic review of natural language processing applied to gastroenterology & hepatology. BMC Gastroenterol. 2025;25(1):58. doi:10.1186/s12876-025-03608-5

23. Klang E, Sourosh A, Nadkarni GN, Sharif K, Lahat A. Evaluating the role of ChatGPT in gastroenterology: a comprehensive systematic review of applications, benefits, and limitations. Ther Adv Gastroenterol. 2023;16:17562848231218618. doi:10.1177/17562848231218618

24. Nehme F, Feldman K. Evolving Role and Future Directions of Natural Language Processing in Gastroenterology. Dig Dis Sci. 2021;66(1):29–40. doi:10.1007/s10620-020-06156-y

25. Roustan D, Bastardot F. The Clinicians’ Guide to Large Language Models: A General Perspective With a Focus on Hallucinations. Interact J Med Res. 2025;14(1):e59823. doi:10.2196/59823

26. Chung P, Swaminathan A, Goodell AJ, et al. Verifying Facts in Patient Care Documents Generated by Large Language Models Using Electronic Health Records. NEJM AI. 2025;3(1):AIdbp2500418. doi:10.1056/AIdbp2500418

27. Feng R, Brennan KA, Azizi Z, et al. Engineering of Generative Artificial Intelligence and Natural Language Processing Models to Accurately Identify Arrhythmia Recurrence. Circ Arrhythm Electrophysiol. 2025;18(1):e013023. doi:10.1161/CIRCEP.124.013023

28. Liu NF, Lin K, Hewitt J, et al. Lost in the Middle: How Language Models Use Long Contexts. arXiv. Preprint posted online November 20, 2023:2307.03172. doi:10.48550/arXiv.2307.03172

29. Lee D, Vaid A, Menon KM, et al. Using Large Language Models to Automate Data Extraction From Surgical Pathology Reports: Retrospective Cohort Study. JMIR Form Res. 2025;9(1):e64544. doi:10.2196/64544

30. Far AT, Bastani A, Lee A, et al. Evaluating the positive predictive value of code-based identification of cirrhosis and its complications utilizing GPT-4. Hepatology. 2025;81(6):1753. doi:10.1097/HEP.0000000000001115

31. Spitzer P, Hendriks D, Rudolph J, et al. The effect of medical explanations from large language models on diagnostic decisions in radiology. medRxiv. Preprint posted online March 6, 2025:2025.03.04.25323357. doi:10.1101/2025.03.04.25323357

32. Kojima T, Gu SS, Reid M, Matsuo Y, Iwasawa Y. Large language models are zero-shot reasoners. In: Proceedings of the 36th International Conference on Neural Information Processing Systems. NIPS ‘22. Curran Associates Inc.; 2022:22199–22213.

33. Reynolds L, McDonell K. Prompt Programming for Large Language Models: Beyond the Few-Shot Paradigm. In: Extended Abstracts of the 2021 CHI Conference on Human Factors in Computing Systems. CHI EA ‘21. Association for Computing Machinery; 2021:1–7. doi:10.1145/3411763.3451760

34. Wan X, Sun R, Dai H, Arik SO, Pfister T. Better Zero-Shot Reasoning with Self-Adaptive Prompting. arXiv. Preprint posted online May 23, 2023:2305.14106. doi:10.48550/arXiv.2305.14106

35. Wang L, Xu W, Lan Y, et al. Plan-and-Solve Prompting: Improving Zero-Shot Chain-of-Thought Reasoning by Large Language Models. arXiv. Preprint posted online May 26, 2023:2305.04091. doi:10.48550/arXiv.2305.04091

36. Dang H, Mecke L, Lehmann F, Goller S, Buschek D. How to Prompt? Opportunities and Challenges of Zero- and Few-Shot Learning for Human-AI Interaction in Creative Applications of Generative Models. arXiv. Preprint posted online September 3, 2022:2209.01390. doi:10.48550/arXiv.2209.01390

37. Miao J, Thongprayoon C, Suppadungsuk S, Krisanapan P, Radhakrishnan Y, Cheungpasitporn W. Chain of Thought Utilization in Large Language Models and Application in Nephrology. Medicina (Mex). 2024;60(1):148. doi:10.3390/medicina60010148

38. Liu J, Liu F, Wang C, Liu S. Prompt Engineering in Clinical Practice: Tutorial for Clinicians. J Med Internet Res. 2025;27:e72644. doi:10.2196/72644

39. Li Y, Ramprasad R, Zhang C. A Simple but Effective Approach to Improve Structured Language Model Output for Information Extraction. arXiv. Preprint posted online February 20, 2024:2402.13364. doi:10.48550/arXiv.2402.13364

40. Gu B, Shao V, Liao Z, et al. Scalable information extraction from free text electronic health records using large language models. BMC Med Res Methodol. 2025;25(1):23. doi:10.1186/s12874-025-02470-z

41. Li Y, Wang H, Yerebakan HZ, Shinagawa Y, Luo Y. FHIR-GPT Enhances Health Interoperability with Large Language Models. NEJM AI. 2024;1(8):AIcs2300301. doi:10.1056/AIcs2300301

42. Hatem R, Simmons B, Thornton JE. A Call to Address AI “Hallucinations” and How Healthcare Professionals Can Mitigate Their Risks. Cureus. 2023;15(9). doi:10.7759/cureus.44720

43. Stuhlmiller TJ, Rabe AJ, Rapp J, et al. A Scalable Method for Validated Data Extraction from Electronic Health Records with Large Language Models. medRxiv. Preprint posted online February 26, 2025:2025.02.25.25322898. doi:10.1101/2025.02.25.25322898

44. Far AT, Ayati A, Guillot J, Azzam S, Rudrapatna VA, Ge J. Large Language Models can Identify the Presence of MASH and Extract VCTE Measurements from Unstructured Documentation. Dig Dis Sci. Published online November 8, 2025. doi:10.1007/s10620-025-09539-1

45. Masoumi S, Amirkhani H, Sadeghian N, Shahraz S. Natural language processing (NLP) to facilitate abstract review in medical research: the application of BioBERT to exploring the 20-year use of NLP in medical research. Syst Rev. 2024;13(1):107. doi:10.1186/s13643-024-02470-y

46. Valsecchi MG, Steliarova-Foucher E. Cancer registration in developing countries: luxury or necessity? Lancet Oncol. 2008;9(2):159–167. doi:10.1016/S1470-2045(08)70028-7

47. Tangka FKL, Subramanian S, Edwards P, et al. Resource requirements for cancer registration in areas with limited resources: Analysis of cost data from four low- and middle-income countries. Cancer Epidemiol. 2016;45:S50–S58. doi:10.1016/j.canep.2016.10.009

48. Curado MP, Voti L, Sortino-Rachou AM. Cancer registration data and quality indicators in low and middle income countries: their interpretation and potential use for the improvement of cancer care. Cancer Causes Control. 2009;20(5):751–756. doi:10.1007/s10552-008-9288-5

49. Bommakanti K, Feldhaus I, Motwani G, Dicker RA, Juillard C. Trauma registry implementation in low- and middle-income countries: challenges and opportunities. J Surg Res. 2018;223:72–86. doi:10.1016/j.jss.2017.09.039

50. Safavi K, Linnander EL, Allam AA, Bradley EH, Krumholz HM. Implementation of a Registry for Acute Coronary Syndrome in Resource-Limited Settings: Barriers and Opportunities. Asia Pac J Public Health. 2010;22(3_suppl):90S–95S. doi:10.1177/1010539510373017

51. Kifle F, Kifleyohanes T, Moore J, Teshome A, Biccard BM. Indications, Challenges, and Characteristics of Successful Implementation of Perioperative Registries in Low Resource Settings: A Systematic Review. World J Surg. 2023;47(6):1. doi:10.1007/s00268-023-06909-6

52. Levine AC, Barry MA, Agrawal P, et al. Global Health and Emergency Care: Overcoming Clinical Research Barriers. Acad Emerg Med. 2017;24(4):484–493. doi:10.1111/acem.13142

53. Choi HS, Song JY, Shin KH, Chang JH, Jang BS. Developing prompts from large language model for extracting clinical information from pathology and ultrasound reports in breast cancer. Radiat Oncol J. 2023;41(3):209–216. doi:10.3857/roj.2023.00633

54. Ni Y, Wright J, Perentesis J, et al. Increasing the efficiency of trial-patient matching: automated clinical trial eligibility Pre-screening for pediatric oncology patients. BMC Med Inform Decis Mak. 2015;15(1):28. doi:10.1186/s12911-015-0149-3

55. Gauthier MP, Law JH, L. Lw, et al. Automating Access to Real-World Evidence. JTO Clin Res Rep. 2022;3(6):100340. doi:10.1016/j.jtocrr.2022.100340

56. Tavabi N, Pruneski J, Golchin S, et al. Building Large-Scale Registries from Unstructured Clinical Notes using a Low-Resource Natural Language Processing Pipeline. medRxiv. Preprint posted online December 26. doi:10.1101/2022.12.23.22283914

57. Wong C, Zhang S, Gu Y, et al. Scaling Clinical Trial Matching Using Large Language Models: A Case Study in Oncology. arXiv. Preprint posted online August 18, 2023:2308.02180. doi:10.48550/arXiv.2308.02180

58. Wyles CC, Fu S, Odum SL, et al. External Validation of Natural Language Processing Algorithms to Extract Common Data Elements in THA Operative Notes. J Arthroplasty. 2023;38(10):2081–2084. doi:10.1016/j.arth.2022.10.031

59. Nori H, Lee YT, Zhang S, et al. Can Generalist Foundation Models Outcompete Special-Purpose Tuning? Case Study in Medicine. arXiv. Preprint posted online November 28, 2023:2311.16452. doi:10.48550/arXiv.2311.16452

60. Zhang X, Talukdar N, Vemulapalli S, et al. Comparison of Prompt Engineering and Fine-Tuning Strategies in Large Language Models in the Classification of Clinical Notes. AMIA Jt Summits Transl Sci Proc. 2024;2024:478–487.

61. Yuan K, Yoon CH, Gu Q, et al. Transformers and large language models are efficient feature extractors for electronic health record studies. Commun Med. 2025;5(1):83. doi:10.1038/s43856-025-00790-1

62. Wiest IC, Leßmann ME, Wolf F, et al. Deidentifying Medical Documents with Local, Privacy-Preserving Large Language Models: The LLM-Anonymizer. NEJM AI. 2025;2(4):AIdbp2400537. doi:10.1056/AIdbp2400537

63. Chianumba EC, Ikhalea N, Mustapha AY, Forkuo AY. NLP Models for Extracting Healthcare Insights from Unstructured Medical Text. Int J Adv Multidiscip Res Stud. 2025;4(6):1533–1553.

64. Turpin M, Michael J, Perez E, Bowman SR. Language Models Don’t Always Say What They Think: Unfaithful Explanations in Chain-of-Thought Prompting. arXiv. Preprint posted online December 9, 2023:2305.04388. doi:10.48550/arXiv.2305.04388

