## Supplementary Material S1 for "General-purpose large language models can achieve physician-level accuracy in complex medical data extraction"

### SUPPLEMENTARY MATERIAL S1 – “The Dataset (N=50)”

### Summary 1

PHTN – Prior Bleeder, Extensive Esophageal Candididasis, Ulceration on right side of esophagus, Eradicated Varices

Cirrhosis - ? NASH ? AIH **(ICD 10 - K75.8)**

Decompensated - Ascites / AVB / HE / Jaundice

S/P PLEX – 3 sessions – May 2024

S/P PLEX – 3 sessions – November 2024

CTP - 12	CHILD-C	MELD Na - 27

**Comorbidities**: Hypertension / Severe Sarcopenia

**CURRENT ISSUES:**

- Sepsis 🡪 Resolving
  - Bilateral Lower Limb Cellulitis
  - Urosepsis – Fungal Urine Culture
  - Infected Penile Ulcer with Epididymo-orchitis
  - B/L Pneumonia
- Hepatic Encephalopathy Grade I / II 🡪 Resolved
- Ascites – Grade III – High SAAG, Low Protein, No SBP – s/p PCD - removed on 28/12/2024
- Advanced Liver Disease 🡪 LT Explained. No donor available at present.

**PRESENTING COMPLAINTS:**

- Worsening Abdominal Distension
- Altered Sensorium

**INDICATION FOR ADMISSION:**

Evaluation and the management of the symptoms

**HISTORY:**

Mr Patient A is a 43 year old gentleman, who had his index presentation in May 2023 at an outside centre with complaints of yellowish discolouration of eyes. Upon evaluation, he was diagnosed as chronic liver disease. He was on routine follow up since. In January 2024, he was admitted in view of worsening jaundice and underwent 3 sessions of PLEX. In April 2024, he had a decompensation in the form of HE and AVB and EVL was done at the outside centre. Over the past two months, he has been complaining of a worsening jaundice associated with a new onset abdominal distension. The yellowish discolouration of his eyes and urine was insidious in onset and progressive. It was not associated with itching or clay-coloured stools. He also complaints of a worsening lower limb swelling since the past one month. He was admitted at an outside centre for the same complaints 1 month ago. He underwent 3 sessions of PLEX and was referred to ILBS for further management. He has now come to ILBS with the above mentioned complaints and has been admitted for further evaluation and management. There is no h/o vomiting, cough, abdominal pain, altered bowel habits, hematemesis, and malena or burning micturition. There is no h/o any intoxications, indigenous medications, blood transfusions or IV drug abuse prior to onset of the disease.

**EXAMINATION**

Pt. was conscious, oriented to time place and person.

BP: - 112/78 mm Hg, Pulse: - 74/min, RR: - 14/min and afebrile.

Pallor-, Icterus+, Cyanosis-, Clubbing-, Pedal edema (pitting type) + , LNP-, JVP normal

On systemic examination

Respiratory system:--B/L vesicular breathing, B/L airway equal air entry, no wheeze,

Cardiovascular system: - S1 S2 normal, no murmurs

CNS: - conscious and oriented, GCS E3V5M6

Per Abdomen examination

ON INSPECTION: - distended abdomen, umbilicus central and inverted, skin over the abdomen is stretched with no visible venous prominences, no visible pulsations.

ON PALPATION: - soft, non-tender, No organomegaly. No guarding, no rigidity, no rebound tenderness.

ON PERCUSSION: - dull note, free fluid present.

ON AUSCULTATION normal bowel sounds present

**SYSTEMATIC REVIEW:**

Patient was admitted with the above mentioned complaints. His examination findings were as mentioned above. He was shifted to the ICU in view of sepsis. Immediate CCM review done and advice followed. He was started on IV fluids, IV antibiotics, antifungals and supportive medications. Blood Cultures revealed no growth, and urine culture revealed a fungal growth. S. PCT – 0.83. Plasma Ammonia – 233.8. Ascitic fluid tap was done which revealed a straw coloured fluid with WBC - 92, N – 32% and L – 67%, Protein: 1.05, Glu: 123, Albumin: 0.3, Gene Xpert: negative, Gram Stain negative. Cultures were negative. VDRL – NR. HSV 1 & 2 – IgG Reactive. He symptomatically improved and was shifted to the HDU on 15/12/2024 and sepsis measures continued. Urology consults were taken in view of epididymo-orchitis and penile ulcer with pus discharge. Advice followed. Blood products were transfused as required. Regular CCM reviews were continued. Plastic surgery, Urology and Dermatology referrals were done in view of penile ulcer and advice followed. Daily dressings were done. PCD was inserted on 18/12/2024 and daily 1-1.5 litres drained. His sensorium and sepsis markers improved and he was symptomatically better. He was shifted to the ward on 23/12/2024. The nature of the disease and long term prognosis was explained in great detail to the patient and the patient relatives and were also explained regarding the need of liver transplant in view of Decompensated Liver Disease. However, no donor is available at present. He is now planned for nutritional optimization and improvement in performance status. Proper diet and mobilisation was followed throughout the course of admission. All vitals, temperature, RBS and other necessary parameters were checked regularly and managed appropriately. He is currently being discharged in hemodynamically stable condition, with the advice to follow in OPD.

**PLAN / ADVICE AT DISCHARGE (Including duration of medication if any):**

2100 KCAL/DAY + 90 GRAMS PROTEIN/DAY, LOW SALT (<2GRAMS/DAY)

**WEIGHT REDUCTION WITH DAILY 30 MIN EXERCISE**

TAB FAROPENAM 200 MG PO BD FOR 5 DAYS

TAB FLUCOS 200 MG PO OD FOR 5 DAYS

T. RIFAGUT 550 MG PO BD	1-0-1 (START ON 6TH DAY)

TAB URSOCOL 450 MG PO BD

CAP HENZOVIT 1 TAB PO OD

TAB ME-12 1 TAB PO OD

CAP HEPAGRESS PO OD

HEPSURE SACHET PO BD

LORHEP SACHET PO BD

SYP LACTIHEP 30ML PO HS  0-0-1 (ENSURE 2-3 BOWEL MOTIONS/DAY)

LAXOPEG SACHET BD. 1-0-1 (ENSURE 2-3 BOWEL MOTIONS/DAY)

FUCIDIN OINTMENT L/A BD

T-BACT OINTMENT L/A BD

TAB MIRAGO-S 25 MG PO OD HS

SYP ZINCONIA 10 ML PO BD

NUTRIX ULTRA WHEY POWDER 2 SCOOPS PO QID WITH MILK *1- 1-1-12PC

INJ. ALBUMIN 20% 100 ML IV TWICE A WEEK UNDER MEDICAL SUPERVISION OVER 4-6 HOURS

REVIEW IN HEPATOLOGY WITH CBC / LFT / KFT / INR REPORTS AFTER 2 WEEKS

PLAN: TITRATE DOSE OF DIURETICS AND BETA BLOCKERS ON OPD BASIS

### Summary 2

Locally Advanced Carcinoma Gall Bladder with EHBO

- s/p SEMS – 20/09/2024
- s/p ERCP + Right Posterior Duct Stenting – 30/09/2024

PET-CT (27/09/2024)

- Locally Advanced Carcinoma Gallbladder
- Cystic Duct, Primary Confluence and CHD involved 🡪 EHBDO
- Infiltration of adjacent liver segments
- No e/o lesions elsewhere

**Comorbidities** – None

**Current Issues**:

EHBDO

**PRESENTING COMPLAINTS:**

Fever and Chills

Jaundice

**INDICATION FOR ADMISSION:**

Evaluation and management of current complaints

**HISTORY:**

Mr Patient B is a 50 year old gentleman with no known comorbidities. He had his index presentation at an outside centre 1 month ago with a fever and was diagnosed as typhoid. She received Ceftriaxone and Azithromycin for the same. However, she has noticed a yellowish discolouration of his eyes and urine since 5 days, which was insidious in onset and progressive. It was associated with itching and clay-coloured stools. She was evaluated outside and MRCP done on 17/09/2024 revealed a soft tissue mass at the hepatic duct confluence with dilated hepatic ducts and IHBRD, with suspicion of Klatskins Tumour or GB fossa mass. She presented to ILBS for further evaluation of the same. SEMS insertion of left biliary system was done on 20/09/2024. Cytology revealed atypical cells. There is no h/o abdominal pain, hematemesis, abdominal distension and malena, burning micturition, altered sensorium or decreased urine output. There is no h/o any intoxications, indigenous medications, major surgeries, blood transfusions or IV drug abuse prior to onset of the disease.

**EXAMINATION:**

Pt. was conscious, oriented to TPP, afebrile.

BP - 114/50 mm Hg , Pulse -80/min	RR -20 /min

Pallor-, Icterus-, Cyanosis-, Clubbing-, Pedal edema – Mild, LNP-, JVP normal

CVS -S1 S2 normal, no murmurs

Chest- B/L normal air entry & vesicular breathing, no adventitious sounds

CNS - conscious, Oriented, no focal neurological deficit.

P/A ON INSPECTION – No distension, umbilicus central and inverted, no visible venous prominences, no visible pulsations.

ON PALPATION - soft, non-tender, liver and spleen- non palpable. No guarding, no rigidity.

ON PERCUSSION - Tympanic note, no free fluid

ON AUSCULTATION- normal bowel sounds present, no bruits.

**SYSTEMATIC REVIEW:**

Patient was admitted with the above mentioned complaints. Her examination findings were as mentioned above. Her lab data is attached at the end of the summary. Urine RME was unremarkable. Serum PCT – 0.12. CEA – 1.341, CA 19-9 was 14.5. AFP – 1.1. Urine and blood C/S revealed no growth.

PET-CT done on 27/09/2024 revealed

- Cholelithiasis with metabolically active ill-defined heterogeneously enhancing circumferential mural thickening involving the gallbladder with infiltration of adjacent liver segments and primary biliary conference and common hepatic duct leading to proximal dilatation and bilobar IHBRD -features are consistent with locally advanced carcinoma gallbladder.
- No evidence of abnormal FDG avid lesions noted elsewhere.

USG screening on 28/09/2024 revealed Bilobar pneumobilia that was not feasible for PTBD. No IHBRD was seen. She underwent an ERCP and stenting on 30/09/2024 with stenting of right ductal system. Medical Oncology and HPB Surgery opinion taken and advice followed. Patients attendants were explained in detail about nature of disease and treatment options. Proper diet and mobilisation was followed throughout the course of admission. All vitals, temperature, RBS and other necessary parameters were checked regularly and managed appropriately. She is discharged in hemodynamically stable condition with the advice to follow up in OPD.

**PLAN / ADVICE AT DISCHARGE (Including duration of medication if any):**

2000 Kcal/day, normal diet

TAB TAXIM-O 200MG BD	1-0-1 X 7 DAYS

TAB ME-12 1 TAB PO OD

CAP HENZOVIT 1 TAB PO OD

TAB URSOCOL SR 450 MG PO BD

TAB VIADEK 1 TAB PO OD

Review in Hepatology, Medical Oncology and HPB Surgery OPD with CBC / LFT / RFT / PT-INR reports after 2 weeks.

### Summary 3

Acute viral hepatitis – HAV IgM positive B 159

**ISSUES –**

Impending ALF

**PRESENTING COMPLAINTS:**

- Fever X 7 days
- SOB X 3 days
- Nausea X 1 day

**INDICATION FOR ADMISSION:**

Evaluation & management of presenting complaints

**HISTORY:**

Miss. Patient C is 18 year old female, who was apparently asymptomatic when he developed fever for 7 days. It was associated with shortness of breath for last 3 days. He presented to an outside centre, IgM HAV was positive. He came to ILBS for further management. There is no h/o past major surgeries, or IV drug abuse prior to onset of the disease. There is no h/o HTN/CAD/COPD/T2DM/Thyroid disorder.

**EXAMINATION:**

Pt. was conscious, oriented to time, place and person

BP: - 110/76 mm Hg, Pulse: - 112/min, RR: - 20/min, Spo2 98% on RA.

Pallor -, Icterus +, Cyanosis-, Clubbing-, Pedal edema -, LNP-, JVP Normal

On Systemic Examination

Respiratory System: - B/L AE equal, no wheeze, no crepts

Cardiovascular system: - S1 S2 normal, no murmurs

CNS: - conscious, oriented to time, place and person

Per Abdomen examination

ON INSPECTION: - Not distended abdomen, umbilicus central and inverted, with no visible venous prominences, no visible pulsations.

ON PALPATION: - Soft, non-tender, liver and Spleen non palpable. No guarding, no rigidity, no rebound tenderness.

ON PERCUSSION: - Tympanic note, No free fluid is present

ON AUSCULTATION: - Diminished bowel sounds present

**SYSTEMATIC REVIEW:**

Patient was admitted with above mentioned complaints. Her examination findings were as mentioned above. Her initial lab data Hb/TLC/Plt: 10.3/4.25/1.35, Bil(T/D): 6.88/4.5, AST/ALT: 4455/3778, ALP: 133, TP/INJ. ALBUMIN 20% 100 ML IV ONCE A WEEK UNDER MEDICAL SUPERVISION OVER 4-6 HOURS: 6.44/3.67, Ammonia: 178, INR: 1.75. Anti HAV – IgM reactive (outside). Anti HB core and Anti HCV – NR. Her Ig M Anti HAV is positive. She was admitted to ICU. In view of increased INR she underwent one session of high volume PLEX in ICU. Post PLEX her sensorium and INR was near normal and she had no fever. She was later on shifted to ward from ICU and care was continued. Proper diet and mobilization was followed throughout the course of admission. All vitals, temperature, RBS and other necessary parameters were checked regularly and managed appropriately. He recovered symptomatically following the management and is discharged with the advice to follow up in OPD.

At the time of discharge her INR: 1.34, Bil (T/D): 7.5/4.67, AST/ALT: 757/1344, TP/Alb: 6.04/3.27, Ceruloplasmin: 0.19

**PLAN / ADVICE AT DISCHARGE (Including duration of medication if any):**

2100 KCAL/DAY + 90 GRAMS PROTEIN/DAY

AVOID ANY ALTERNATIVE MEDICATIONS

TAB URSOCOL 450 MG PO BD

CAP HENZOVIT 1 TAB PO OD

Avoid all sort of CAM

PLAN: CBC/ LFT/RFT AND INR AFTER ONE WEEK AND INFORM VIA VIRTUAL OPD

OPD after 2 weeks

IN CASE OF DECREASE IN URINE OUTPUT, ALTERED SENSORIUM, BLEEDING, FEVER, NEW ONSET COUGH AND REVIEW IN ILBS EMERGENCY ON SOS BASIS.

### Summary 4

Portal Hypertension - (Non Bleeder, Grade II Esophageal varices)

Chronic Liver Disease - HBV related (HBV DNA 5 log 3-->ND )

Compensated

HCC

- 4.8 x 4.2 x 5.1 cm in Segment V - S/P TACE on 15/09/2022 🡪 Complete Response
- 18 mm in Segment VII - S/P MWA on 19/07/2023 🡪 Complete Response
- Last PET-CT on 02/05/2024 🡪 Complete Response
- **New LR 3/4 Lesion Segment V - S/P MWA on 25/11/2024**

USG guided biopsy from normal liver parenchyma - Chronic hepatitis with mild activity and thin bridging fibrosis, Modified Histological activity index- 6/18, Fibrosis – 2-3 on 25/11/2024

USG guided biopsy from Liver SOL – 25/11/2024 - Awaited

AFP - 2 log4 (Aug 2022) 🡪 27 🡪 94 🡪 109 🡪 268 🡪733.5 on 23/11/2024

PIVKA II – 28 (Sept 2023) 🡪 30 🡪 167.41 🡪 507.70 on 23/11/2024

PS:-0	BCLC: B	CTP - 5A, MELD Na - 7

**Co morbidities** - T2DM/HTN

**ICD-K74.60/I10/E14.9/C22.0**

**Current issues:** New LR 3/4 Lesion Segment V - S/P MWA on 25/11/2024

**CHIEF COMPLAINTS**

Admitted with plan for MWA

**INDICATION FOR ADMISSION:**

Admitted for management of CLD with HCC

**HISTORY**

Mr Patient D, 61 year old non-smoker non-ethanolic and co morbidities of T2DM and hypertension on regular medication for the same. Index issue is in the form of abdominal pain in 2023 Feb. Patient was admitted for the same and was diagnosed with CLD HBV related with HCC in segment V for which he underwent TACE on 15/09/2022. Follow up imaging showed New Lesion in Seg VII measuring 18mm for which he underwent MWA on 19/07/2023. Now, patient has been admitted in ILBS as planned for immunotherapy in view of rising tumour markers and new detected lesion in Segment V – feasible for MWA. There is no h/o fever, jaundice, vomiting, cough, abdominal pain, altered bowel habits, hematemesis, and malena, burning micturition, altered sensorium or decreased urine output. There is no h/o any intoxications, indigenous medications, major surgeries, blood transfusions or IV drug abuse prior to onset of the disease. There are no h/o CAD/TB/COPD/Thyroid disorders.

**EXAMINATION**

Pt. was conscious, oriented to time place and person.

BP: - 134/72mm Hg, Pulse: - 86/min, RR: - 20/min and afebrile.

Pallor-, Icterus-, Cyanosis-, Clubbing-, Pedal edema (pitting type) - , LNP-, JVP normal

On systemic examination

Respiratory system: B/L vesicular breathing, B/L airway equal air entry, no wheeze, no crepts

Cardiovascular system: - S1 S2 normal, no murmurs

CNS: - conscious and oriented with no sensorimotor deficit,

Per Abdomen examination

ON INSPECTION: - non distended abdomen, umbilicus central and inverted, skin over the abdomen is stretched with no visible venous prominences, no visible pulsations.

ON PALPATION: - soft, non-tender, liver and Spleen non- palpable. No guarding, no rigidity, no rebound tenderness.

ON PERCUSSION: - Tympanic note with no fluid present

ON AUSCULTATION normal bowel sounds present,

**SYSTEMATIC REVIEW:**

Patient was admitted with above mentioned complaints. His examination findings were as mentioned above. His initial lab data and latest lab data is included at last of summary. Patient was treated with IV antibiotics, nutritional support and other supportive medication and treatment. The nature of the disease and long term prognosis was explained in great detail to the patient and the patient relatives. The risks and benefits of therapy options were explained, and the decision was taken to undergo Microwave Ablation. Patient underwent MWA of segment VII lesion on 25/11/2024. The procedures were tolerated well with no post procedural complications. Post – MWA screening revealed no peri-hepatic free fluid. Percutaneous biopsy was also taken from non-tumerous liver and the liver SOL. USG guided biopsy from normal liver parenchyma - Chronic hepatitis with mild activity and thin bridging fibrosis. USG guided biopsy from Liver SOL – report awaited. He is now planned for nutritional optimization and improvement in performance status. Proper diet and mobilisation was followed throughout the course of admission. All vitals, temperature, RBS and other necessary parameters were checked regularly and managed appropriately. He recovered symptomatically following the management and is discharged in hemodynamically stable condition with following advice to follow in OPD.

**PLAN / ADVICE AT DISCHARGE (Including duration of medication if any):**

1800kcal + 80 grams protein, low salt diet

TAB TAXIM-O 200MG BD	1-0-1 X 5 DAYS

TAB CARDIVAS 6.25 MG PO BD (DO NOT GIVE IF HR < 55 OR BP < 90/60 MMHG)

TAB CILACAR 5 MG PO SOS (DO NOT GIVE IF BP < 90/60 AND HR < 55)

TAB LENVATINIB 4 MG PO OD

TAB TAF 25 MG PO OD

HEPSURE SACHET PO BD

TAB URSOCOL SR 450 MG PO BD

TAB THYRONORM 75 MCG PO BBF

DIABETIC MEDICATIONS TO CONTINUE AS BEFORE

Review in Hepatology OPD with CBC / LFT / KFT / INR reports after 4 weeks

IN CASE OF DECREASE IN URINE OUTPUT, ALTERED SENSORIUM, BLEEDING VISIT ER SOS

### Summary 5

Portal Hypertension (Non-Bleeder, Grade I Esophageal Varices, Mild PHG - 22/10/2024)

CLD - NASH (ICD-K75.6)

HVPG-10 🡪 6 mm HG

Compensated

CAP 236	LSM 22.5	SSM 35.1

s/p Partial Splenic Artery Embolization on 26/03/2025 for Pancytopenia with Massive Splenomegaly

**CURRENT ISSUES**

- Pancytopenia: s/p PRBC and Platelet transfusions

**Comorbidity:** T2 DM (controlled, on OHAs)

Hypertension

Hypothyroidism

                         Beta thalassemia trait

                         HbD (Punjab) heterozygosity

Lower CBD stricture with microsludge S/P- ERCP + EPT done - 2021

Atrial Flutter--On Ditiazem

**PRESENTING COMPLAINTS:** Generalized Weakness and Fatigue

**INDICATION FOR ADMISSION:**

Evaluation and management of symptoms

**HISTORY:**

Mr. Patient E is a 59 year old gentleman, who is a known case hypertensive and diabetic. He had his index presentation was 5 years ago when he presented with a fatigue and was found to have low Hb. On evaluation he was found to have a cirrhotic liver. He has been requiring 2-3 PRBC transfusions yearly. 4 years ago he developed a jaundice and MRCP done revealed lower CBD stone. He underwent ERCP + EPT for the same. Currently he has presented with complaints of pedal edema since 2 weeks, and his Hb was 4.9 on routine investigations, he was previously evaluated and BM- ?primary MF. Outside hematology opinion obtainedand has given INJ DARBOPIETIN 5000 3doses. Partial splenic artery embolization was done by IR team on 26/03/2025. Post PSAE USG screening revealed no ascites / puncture site hematoma or bleeding. There is no h/o vomiting, cough, abdominal pain, altered bowel habits, hematemesis, and malena, burning micturition, altered sensorium or decreased urine output. There is no h/o any intoxications, major surgeries, blood transfusions or IV drug abuse prior to onset of the disease. There is no h/o CAD/TB/COPD/Thyroid disorder.

**EXAMINATION:**

Pt. was conscious, oriented, afebrile.

BP-110/72 mm Hg , Pulse 76/min	RR -18/min

Pallor+, Icterus +, Cyanosis-, Clubbing-, Pedal edema-, LNP-, JVP normal

CVS S1 S2 normal, no murmurs

Chest B/L normal air entry & vesicular breathing, no adventitious sounds

CNS Patient conscious, oriented, no sensorimotor deficit.

P/A ON INSPECTION -non-distended, umbilicus central and inverted, no visible venous prominences, no visible pulsations.

ON PALPATION: soft, liver not palpable. Spleen palpable 3 cm below SCM. No guarding, no rigidity, no rebound tenderness.

ON PERCUSSION tympanic note, no free fluid

ON AUSCULTATION normal bowel sounds present, no bruit

**SYSTEMATIC REVIEW:**

Patient was admitted with the above mentioned complaints. His examination findings were as mentioned above. Blood and Urine Cultures revealed no growth, and Urine Routine was unremarkable. S. PCT 0.19. HE had two episodes of fever and antibiotics upgraded. Blood products were transfused as required. IR screening was done - Spleen size ~ 22.4 cm. 12.5 x 8 cm infarct in lower pole. Tapping was done - 200cc fluid removed which revealed normal study. Haematology consults were taken and advise followed. Case was discussed with IR team and haematologist – advised conservative management for now in view of window period of therapeutic response of PSAE. Haematology advice – supportive care and Inj GCSF if ANC < 1000 and Romiplastim if Plt < 20k. Patients attendants were explained in detail about nature of disease and treatment options. Proper diet and mobilisation was followed throughout the course of admission. All vitals, temperature, RBS and other necessary parameters were checked regularly and managed appropriately. He is discharged in hemodynamically stable condition with the advice to follow up in OPD.

**PLAN / ADVICE AT DISCHARGE (Including duration of medication if any):**

1800 KCAL/DAY + 90 GRAMS PROTEIN/DAY, NORMAL DIET

DAILY 30 MIN MUSCLE STRENGTHENING EXERCISE

INJ DARBOPOETIN 40 MCG S/C EVERY 2 WEEKS

INJ ROMIPLASTIM 250 MCG S/C ONCE A WEEK

INJ ELORES 1.5 GM IV BD X 5 DAYS

INJ MOXIFLOXACIN 400 MG IV OD X 5 DAYS

FOSFOMYCIN SACHET PO OD X 5 DAYS

TAB FLUCOS 200 MG PO OD FOR 5 DAYS

TAB DILTIAZEM 30MG PO BD 1-0-1

TAB URSOCOL 450 MG PO BD

CAP HENZOVIT 1 TAB PO OD

SYRUP DEXORANGE 10 ML PO OD

TAB FOLVITE 5 MG PO OD

LAXOPEG SACHET BD. 1-0-1 (ENSURE 2-3 BOWEL MOTIONS/DAY)

MOVICOL SACHET 1 SACHET PO SOS

HEPSURE SACHET PO BD

DIABETIC MEDICATIONS TO CONTINUE AS BEFORE

REVIEW IN HEPATOLOGY OPD WITH CBC/LFT/KFT/INR REPORTS AFTER 4 WEEKS

CARDIOLOGY OPD FOLLOW UP FOR ATRIAL FLUTTER

IN CASE OF DECREASE IN URINE OUTPUT, ALTERED SENSORIUM, BLEEDING, FEVER, NEW ONSET COUGH AND REVIEW IN ILBS EMERGENCY ON SOS BASIS.

### Summary 6

Post DDLT for Cirrhosis NASH on 30/07/2024 **(ICD10 - Z94.4)**

Early Post OP Event: POD 8 ACR (Clinical) s/p PMP (500-500-250)

**ACR (November 2024)**: Graft Liver TJLB: Acute cellular rejection with RAI 7/9 (portal inflammation 3 duct damage 1 endothelitis 3)

S/P ATG – 50 mg (November 2024)

**CURRENT ISSUE**

- Worsening Hyperbilirubinemia with Skin Rash – Likely Viral Reactivations (CMV, EBV – NR) 🡪 Improving
- Sepsis – Fungal Pneumonia Left Lobe Consolidation – RSV +, Sputum positive for Aspergillus
- Uncompensated Metabolic Acidosis with AKI-AKIN I 🡪 Improved with hydration.

**PRESENTING COMPLAINTS:**

·       Worsening jaundice x 4-5 days

**INDICATION FOR ADMISSION:**

Evaluation and the management of presenting complaints

**HISTORY:**

Mr. Patient F is a 45 years old male, non-diabetic, normotensive with no other prior co morbidity. He underwent a DDLT at an outside centre on 30/07/2024. On POD 8, he had an ACR for which he received Pulse methylprednisolone. Further post-operative period was uneventful. He was admitted at ILBS in September 2024 with complaints of loose stools and a generalised weakness and for Uropsepsis with culture showing Pseudomonas and for AKI KDIGO III Non-oliguric - Creatinine 3.62-->0.9 and was subsequently discharged in hemodynamically stable condition.Now he has presented with complaints of worsening jaundice since one week which was painless, with yellowish discoloration of sclera and urine, insidious onset, gradually progressive, not associated with pruritus and associated with clay-colored stools.It was also associated with easy fatiquibility and generalized weakness.There is history of non compliance to medication for 10 days prior to onset of jaundice. Recently came to ILBS with worsening of jaundice on evaluation Biopsy done s/o of Acute cellular rejection with RAI 7/9 (portal inflammation 3 duct damage 1 endothelitis 3) S/P Glucocorticoid Pulse Therapy (Inj Methylprednisolone 500/500/250/250/100/100mg and planned for ATG if LFT worsening . Now again came with Worsening of jaundice from 3-4 days . Admitted for further evaluation and management There is no h/o fever, , vomiting, cough, abdominal pain, altered bowel habits, hematemesis, and malena, burning micturition, altered sensorium or decreased urine output. There is no h/o any intoxications, indigenous medications, major surgeries, blood transfusions or IV drug abuse prior to onset of the disease. There is no h/o DM/HTN/CAD/TB/COPD/Thyroid disorders.

**EXAMINATION**

Pt. was conscious, oriented to time place and person.

BP: - 120/74 mm Hg, Pulse: - 82/min, RR: - 18/min and afebrile.

Pallor-, Icterus+, Cyanosis-, Clubbing-, Pedal edema (pitting type) - , LNP-, JVP normal

On systemic examination

Respiratory system: B/L vesicular breathing, B/L airway equal air entry, no wheeze

Cardiovascular system: - S1 S2 normal, no murmurs

CNS: - conscious and oriented with no sensorimotor deficit

Per Abdomen examination

ON INSPECTION: - LT Scar noted - healed, no distension, umbilicus central and inverted, no visible venous prominences, no visible pulsations.

ON PALPATION: - soft, non-tender, liver and spleen is non- palpable. No guarding, no rigidity, no rebound tenderness.

ON PERCUSSION: - Tympanic note, no free fluid present

ON AUSCULTATION normal bowel sounds present,

**SYSTEMATIC REVIEW:**

Patient was admitted with above mentioned complaints. His examination findings were as mentioned above. Blood and Urine Cultures revealed no growth, and Urine Routine was unremarkable. S. PCT – 0.63. CMV DNA Not Detected. He was admitted in the HDU in view of uncompensated metabolic acidosis and AKI. He was treated with IV antibiotics, nutritional support and other supportive medication and treatment. He was initiated on Hydrocortisone. CXR revealed left lobe consolidation and Pulmonology consults done. Sputum revealed septate fungal hyphae and culture grew Aspergillus. RV Panel – Positive. In view of fungal pneumonia, his antibiotics and antifungals were upgraded. Transaminitis and AKI gradually improved, and he was shifted to the ward on 13/12/2024. HRCT Lung was done on 14/12/2024 – final report awaited. The nature of the disease and long term prognosis was explained in great detail to the patient and the patient relatives and were also explained regarding the need of repeat liver transplant. LT referral was done. However, no donor is available at present and they wish to continue with medical management. Proper diet and mobilisation was followed throughout the course of admission. All vitals, temperature, RBS and other necessary parameters were checked regularly and managed appropriately. He recovered symptomatically following the management and now is hemodynamically stable and is being discharged with the advice to follow up in OPD.

**PLAN / ADVICE AT DISCHARGE (Including duration of medication if any):**

2000 KCAL/DAY + 90 GRAMS PROTEIN + 30GM FAT /DAY

DAILY 30 MIN EXERCISE AND MUSCLE STRENGTHENING EXERCISE

INJ CASPOFUNGIN 35 MG IV OD FOR 7 DAYS

TAB CILACAR 10 MG PO SOS (DO NOT GIVE IF BP < 90/60 AND HR < 55)

ARKAMINE 0.1 MG PO TDS

TAB URSOCOL 450 MG PO BD

TAB NODOSIS 500 MG PO TDS

TAB LEVOFLOX 500 MG PO OD X 5 DAYS

T. RIFAGUT 550 MG PO BD	1-0-1 (START ON 6TH DAY)

TAB MAXILIV 500 MG PO BD

REVIEW IN HEPATOLOGY OPD WITH CBC/LFT/KFT/INR REPORTS AFTER 4 WEEK

REPEAT KFT AFTER SEVEN DAYS AND INFORM VIA VC

IN CASE OF FEVER, JAUNDICE, PAIN , BLEEDING / BLACKISH MOTION - REVIEW IN ILBS EMERGENCY SOS.

### Summary 7

**LEAVE AGAINST MEDICAL ADVICE**

PHTN- (Non-Bleeder – Small Esophageal Varices, Mild PHG – October 2023

Cirrhosis - NAFLD + Alcohol related (Last Intake – 1.5 years, Max Weight – 102 Kg)

Decompensated - Ascites / HE / Jaundice

CTP – 10 C	MELD – 21

**(ICD 10 - K74.6 + K70.3)**

**CO-MORBIDITIES**:

- Type II Diabetes Mellitus
- Hypothyroidism
- Umbilical Hernia

**CURRENT ISSUE:**

- Sepsis with SBP (outside culture – E.coli) 🡪 Resolving
- Acute on Chronic Kidney Disease – HRS - KDIGO II – Non-oliguric, s/p SLED – 2 sessions, ( S. Creatinine 1.52 🡪 1.37), Urine PCR – 0.58, Urinary Na – 8
- Hypovolemic Hyponatremia - Urinary Na – 8
- Refractory Ascites – Grade III – (High SAAG, Low Protein, SBP – outside culture – E.coli) s/p PCD insertion – 30/01/2025, removed on 13/02/2025
- Bilateral Leg and Scrotal Swelling – Cellulitis
- Advanced Liver Disease - **LT Explained.**

**PRESENTING COMPLAINTS:**

- Worsening Abdominal Distension

**INDICATION FOR ADMISSION:**

For further management & evaluation of current symptoms

**HISTORY:**

Mr. Patient G is a 49 old gentleman, with prior co morbidity of Diabetes Mellitus on regular medications for the same. Patient had his index presentation at an outside centre in October 2023 in the form of jaundice. On further evaluation he was found to have CLD-NAFLD + Alcohol related. Since then he has been on regular follow up at outside hospital. He had ascites from May 2024 which improved on diuretics. He has been admitted previously in view of HE at the outside centre in August 2024 and was conservatively managed. He currently presented to ILBS with the complaints of worsening abdominal distension and pedal edema. He has been admitted for further evaluation and management. There is no h/o indigenous medications, major surgeries, blood transfusions prior to onset of the disease. There are no h/o HTN/COPD/BA/ TB/ Renal disorders.

**EXAMINATION:**

Pt. was conscious, well oriented, a febrile.

BP-118/80 mm Hg	Pulse 82/min	RR 20/min SP 02 : 93%

Pallor+, Icterus-, Cyanosis-, Clubbing-, Pedal edema-, LNP-, JVP normal

CVS -S1 S2 normal, no murmurs

Chest- Air entry decreased right side.

CNS Conscious, oriented

P/A examination

ON INSPECTION- moderately distended, umbilicus central and inverted, visible venous prominences, no visible pulsations.

ON PALPATION- soft, non tender, hepatomegaly 2 cm BCM. Spleen is just palpable. No guarding, no rigidity, no rebound tenderness.

ON PERCUSSION- dull note present, free fluid present

ON AUSCULTATION- normal bowel sounds present, no bruits.

**SYSTEMATIC REVIEW:**

Patient was admitted with the above mentioned complaints. His examination findings were as mentioned above. His initial and final lab data is attached at the end of the summary. Plasma ammonia – 179.3. Blood Cultures revealed no growth, and Urine Routine revealed 6-7 leukocytes with full field RBCs and Yeast 4+. S. PCT – 0.21. Patient was treated with IV antibiotics, nutritional support and other supportive medication and treatment. Ascitic fluid PCD insertion was done which revealed a reddish haemorrhagic fluid with WBC - 112, N – 35% and L – 64%, Protein: 0.97, Glu: 132, Albumin: 0.53, SAAG: 2.41, ADA: 2.5 and Gene Xpert: negative, Gram Stain negative. Cultures were negative. Pulmonology and Nephrology consults taken and advice followed. Ophthalmology referral was done which revealed subconjunctival haemorrhage and mild ecchymoses with normal fundus; advice followed. He was closely monitored and electrolyte correction and AKI management done. Urology referrals done in view of scrotal swelling and urinary obstruction – Foleys catheterization done. In view of non-oliguric AKI an d fluid overload, he underwent 2 sessions of SLED. Blood products transfused as requied. The nature of the disease and long term prognosis was explained in great detail to the patient and the patient relatives and were also explained regarding the need of liver transplant in view of Decompensated Liver Disease. However, no donor is available. Strict alcohol abstinence advised. LT referral done. Proper diet, purging and mobilisation was followed throughout the course of admission. All vitals, temperature, RBS and other necessary parameters were checked regularly and managed appropriately. Presently he is conscious, oriented and hemodynamically stable. He is being **discharged on request** in stable hemodynamic state with advice of follow up in OPD.

**ADVICE ON DISCHARGE (Duration of medication if any):**

INJ IMIPENAM 500 MG IV TDS

INJ CLINDAMYCIN 600 MG IV TDS

TAB FLUCOS 200 MG PO OD FOR 5 DAYS

HEPSURE SACHET PO BD

NUTRIX ULTRA WHEY POWDER 2 SCOOPS PO QID WITH MILK 1- 1-1-1—2PC

TAB MELATONIN 5 MG PO SOS / HS

SYP LACTIHEP 30ML PO HS  0-0-1 (ENSURE 2-3 BOWEL MOTIONS/DAY)

LAXOPEG SACHET BD. 1-0-1 (ENSURE 2-3 BOWEL MOTIONS/DAY)

LORHEP SACHET PO BD

CAP HENZOVIT 1 TAB PO OD

TAB ME-12 1 TAB PO OD

TAB THYROXINE 75 MCG PO OD BBF

PLAN: LT WORKUP

### Summary 8

PHTN-Bleeder, Non-Bandable Esophageal Varices with Extensive Background Scarring and Neovascularization, Mild PHG - 12/12/2024

Cirrhosis: NASH related **{K 74.6}**

Decompensated: AVB | Ascites | Jaundice

Infiltrative HCC with PVTT (VP 2 TT) **{C 22.0}**

- Seg VII: 5.4 x 3.8 x 4.1
- S/P SBRT (5 sessions 18/11/22)
- ON ICI Regimen
- S/P 7 Doses of Inj. Nivolumab 240mg (Last cycle on 17/03/2024)
- S/P Inj. Atezolizumab 1200mg two Doses
- S/P Inj. Atezolizumab 1200mg + Inj. Bevacizumab 1000mg 4 doses
- S/P Restarted on InjNivolumab with Ninth dose given on 16/10/24
- S/P VMAT For new aortocaval and para-aortic lymph nodes 26/09/24 to 10/10/24
- First dose Tremelimumab 300mg + Durvalumab 1500mg given on 19/11/24

AFP- 14 🡪57🡪498🡪96 🡪13->388->1774.0-->2008🡪2114-->1938-->59

PIVKA II -910🡪124 🡪8944🡪303🡪1491->428->1067-->975🡪80-->237-->124   CA19.9🡪21

MELD Na -14         CTP- 8 B	BCLC-D

**Co morbidities**- T2DM/ HTN

S/P appendectomy 30 years ago

**CURRENT ISSUES**-

- Acute Variceal Bleed – S/P UGIE - Non-Bandable Esophageal Varices with Extensive Background Scarring and Neovascularization, Mild PHG - 12/12/2024
- Sepsis – SBP 🡪 Resolved on Response Tap

**PRESENTING COMPLAINTS:**

Malena x 2 days

**INDICATION FOR ADMISSION:**

For further evaluation and management CLD with HCC

**HISTORY:**

Mr Patient H is a 59 years old male with known co morbidity of T2DM and HTN. He had his index presentation in October 2022 in the form of pain in abdomen and on evaluation he was diagnosed as a case of HCC. He has had 5 SBRT sessions in November 2022. He was last admitted at ILBS in march 2023 with complaints of pain abdomen and melena for which he underwent UGIE on 21/3/23 which was s/o small high risk esophageal varices for which he underwent EVL; antral GAVE with active ooze . APC was done and patient was subsequently discharged in haemodynamically stable condition. He was again admitted 04 to 10 april 2023 with c/o of malena with no history of postural symptoms and UGI endoscopy was done to investigate the bleed, which showed small low risk esophageal varices. Antral GAVE for which he underwent APC. Post Bleed he underwent HVPG which was 16mmHg. In his recent admission UGI endoscopy was done which showed grade II esophageal varices with RCS for which he underwent EVL. PET scan was done which showed disease recurrence with multiple new liver lesions. He was planned for immunotherapy and 7 doses have been administered (last cycle on 17/03/2024). His last admission was with complaints of melena which was managed conservatively. Interval imaging show active lesion with arterializations of veins. With plan for new ICI therapy regimen he has been admitted. Immunotherapy with at atezolizumab and bevacizumab was initiated. 4 sessions of therapy with both drugs were given and 2 cycle of atezolizumab was given. For history of melena he underwent UGIE S/P EVL done.He was restarted on Injnivolumab and subsequently was given two dose of nivolumab 240mg with last on 16/10/24 and also underwent VMAT For new aortocaval and para-aortic lymph nodes 26/09/24 to 10/10/24. He was admitted for STRIDE regimen (tremelimumab + Durvalumab) andFirst dose Tremelimumab300mg +Durvalumab 1500mg was given on 19/11/24. He has currently presented with complaints of blackish altered stools since 2 days. He was admitted for management of bleed and further management of CLD with HCC.There is no h/o vomiting, altered sensorium, burning micturition or decreased urine output. There is no h/o any intoxications, indigenous medications, IV drug abuse prior to onset of the disease. There is no h/o thyroid disorders/CAD/COPD/genetic or hereditary disorders.

**GENERAL EXAMINATION:**

Pt. was conscious, oriented to TPP, febrile.

BP -116/78 mm Hg, Pulse -76/min;  RR -16/min

Pallor-, Icterus-, Cyanosis-, Clubbing-, Pedal enema - , JVP normal

CVS -S1 S2 normal, no murmurs

Chest- B/L normal air entry & vesicular breathing, no adventitious sounds

CNS - conscious, oriented, no focal neurological deficit

P/A ON INSPECTION- mildly distended, umbilicus central and inverted, no visible venous prominences, No visible pulsations.

ON PALPATION- soft, non tender, liver just palpable BCM and spleen non palpable, No guarding, no rigidity, no rebound tenderness

ON PERCUSSION-dull note, free fluid present.

ON AUSCULTATION- normal bowel sounds present, no bruits.

**SYSTEMATIC REVIEW:**

Patient was admitted with above mentioned complaints. His examination findings were as mentioned above. His initial lab data and latest labs are at end of summary in chart form. Blood Cultures revealed no growth. He was admitted in view of hematemesis and started on aggressive management with RT lavage, fluid resuscitation with IV fluids, IV antibiotics, and other supportive measures. He was started on IV Terlipressin and blood products in the form of PRBC were transfused to correct acute blood loss. UGI endoscopy was done to investigate the bleed, which showed Non-Bandable Esophageal Varices with Extensive Background Scarring and Neovascularization, Mild PHG - 12/12/2024. Ascitic fluid tap was done which revealed a straw coloured fluid with WBC - 515, N – 52% and L – 47%, Protein: 0.54, Glu: 164, Albumin: 0.3, SAAG: 2.7, ADA: 1.3. Antibiotics were upgraded in view of SBP. Response tap was done on 14/12/2024 revealed TLC – 442 with N – 78 % and L – 21 %.Proper diet and mobilization was followed throughout the course of admission. Nutritional optimization has been done and patient is tolerating orally well. The advanced nature of the disease and its long term prognosis and limited treatment options was explained in great detail to the patient and the patient attendant. Now he is being discharged in hemodynamically stable state with the advice to follow up in OPD.

**PLAN / ADVICE AT DISCHARGE (Including duration of medication if any):**

DIET AS ADVISED BY DIETICIAN (TARGET 1800KCAL/DAY, 70GM PROTEIN/DAY, SALT RESTRICTED)

TAB FAROPENAM 200 MG PO BD FOR 5 DAYS

T. RIFAGUT 550 MG	BD	1-0-1 DAY 6 ONWARDS

TAB THYRONORM 50MCG PO OD BBF 1-0-0

TAB LENVATINIB 4MG PO OD 1-0-0

TAB VOZAN 20MG PO OD 1-0-0

CAP HENZOVIT PO OD 1-0-0

TAB ME 12 PO 1-0-0

TAB MESACOL 400MG PO BD 1-0-1

TAB JANUMET (50/500) PO BD 1-0-1 (MONITOR RBS)

TAB GABAPIN 100MG PO OD/HS 0-0-1

HEPSURE SACHET PO BD 1-0-1

CAP HEPAGRESS PO OD 1-0-0

SOFTAVAC GRANULES 2 TSF 1-0-1

MOVICOL SACHET PO TDS 1-1-1

TAB PRUEASE 1MG PO BD 1-0-1

TAB ULTRACET PO BD 1-0-1

FENTANYL PATCH 25MCG OVER 72 HOURS SOS

CAP MYORIL 4MG PO SOS

VOLINI GEL FOR PAIN SOS

TAB NAPROXEN 250MG PO SOS

INJ. ALBUMIN 20% 100 ML IV ONCE A WEEK UNDER MEDICAL SUPERVISION OVER 4-6 HOURS

REVIEW IN HEPATOLOGY OPD WITH REPORTS OF CBC/LFT/KFT/INR AFTER 2 WEEKS.

PLAN: RESTART BETA BLOCKERS AND DIURETICS ON OPD BASIS / 2 ND SESSION OF STRIDE AFTER 2 WEEKS

IN CASE OF FEVER, JAUNDICE, PAIN , BLEEDING / BLACKISH MOTION - REVIEW IN ILBS EMERGENCY SOS.

### Summary 9

Portal HTN (Bleeder, Small High Risk Esophageal Varices – EVL done – 10/03/2025)

Cirrhosis - NASH related (ICD10 - K75.8)

Decompensated:-Ascites / AVB

CAP – 181	LSM- 63.6	SSM – 73.1

MELD – 17	CTP – 10 / C

Multifocal HCC with MHV involvement;

- segments II / IVA / IVB / VIII – LR5 lesions with no PVT (ICD10 - C22.0)
- Started on TKI – Sorafenib on 11/03/2025

AFP – 4 log3	CHILD – C ; BCLC - D

**CO-MORBIDITIES**:T2DM / HTN / Obesity

**ICD-K75.6/C22.0/E14.9/I10**

**CURRENT ISSUES:-**

- Malena – on and off – IGIE Small High Risk Esophageal Varices – EVL done – 10/03/2025
- Severe Anemia (Hb 4.9 🡪 7.3) s/p PRBC transfusions
- AKI KDIGO I (S.Creat - 1.23) 🡪 Improving
- Ascites Grade III – High SAAG, Low Protein, No SBP

**INDICATION FOR ADMISSION:**

Evaluation and the management of presenting symptoms

**PRESENTING COMPLAINTS:**

- Distension of abdomen x 2 months
- Bilateral pedal edema x 2 months

**HISTORY:**

Mr Patient I is an 80 years old gentleman who had index presentation in October 2023 in the form of ascites with B/L pedal edema. He was evaluated in ILBS and started on diuretics for the same. He has currently presented to ILBS in view of generalized weakness and worsening abdominal distension. Patient now has come to ILBS with above mentioned complains and for further evaluation and management. There is no h/o fever, dysuria, sweating, altered sensorium, burning micturition or decreased urine output. There is no h/o any intoxications, major surgeries, IV drug abuse prior to onset of the disease. There are no h/o B.Asthma/CAD/Thyroid disorders.

**GENERAL EXAMINATION:**

Pt. was conscious, oriented, afebrile

BP- 128/78 mm Hg     Pulse 70/min RR-18/min

Pallor +, Icterus-, Cyanosis-, Clubbing-, Pedal edema+, LNP-, JVP-normal

CVS -S1 S2 normal, no murmurs.

Chest- B/L equal air entry present, no added sounds.

CNS -conscious, oriented to time,place & person.

P/A examination

ON INSPECTION-distended, umbilicus central and inverted, no visible venous prominences,

no visible pulsations.

ON PALPATION-Soft,non tender, liver and spleen palpable. No guarding, no rigidity, no rebound tenderness.

ON PERCUSSION-dull note, free fluid present.

ON AUSCULTATION-Normal bowel sounds present, no bruits.

**SYSTEMATIC REVIEW:**

Patient was admitted with above mentioned complaints. His examination findings were as mentioned above. His initial lab data and latest labs are at end of summary in chart form. Blood Cultures revealed no growth. S. PCT – 0.12. He was admitted in view of melena and started on aggressive management with fluid resuscitation with IV fluids, IV antibiotics, and other supportive measures. Blood products in the form of PRBC were transfused to correct acute blood loss. UGI endoscopy was done on 10/03/2025 - Small High Risk Esophageal Varices – EVL done. Ascitic fluid tap was done which revealed a straw coloured fluid with WBC - 39, N 1% and L 98%, Protein: 1.45, Glu: 136, Albumin: 0.65, SAAG: 2.02, ADA: 6.3. CECT W/A was done– revealed multifocal HCC with MPV involvement - provisional report. Final report awaited. Sorafenib was started on 11/03/2025. Proper diet and mobilization was followed throughout the course of admission. Nutritional optimization has been done and patient is tolerating orally well. The advanced nature of the disease and its long term prognosis and limited treatment options was explained in great detail to the patient and the patient attendant.  Now he is being discharged in hemodynamically stable state with the advice to follow up in OPD.

**PLAN / ADVICE AT DISCHARGE (Including duration of medication if any):**

DIET AS ADVISED BY DIETICIAN

HOME BP MONITORING

TAB TAXIM-O 200MG BD	1-0-1 X 5 DAYS

TAB SORAFENIB 200 MG PO OD

TAB DYTOR 5 MG PO **EVERY ALETRNATE DAY** (MONITOR KFT WEEKLY)

TAB ARKAMINE 0.1 MG PO TDS (DO NOT GIVE IF BP < 90/60 AND HR < 55)

TAB CILACAR 10 MG PO BD (DO NOT GIVE IF BP < 90/60 AND HR < 55)

LAXOPEG SACHET BD. 1-0-1 (ENSURE 2-3 BOWEL MOTIONS/DAY)

TAB ME-12 1 TAB PO OD

TAB PANTOCID 40 MG PO OD BBF X 14 DAYS

SYP SUCRAL 10 ML PO QID X 14 DAYS

CAP HENZOVIT 1 TAB PO OD

INJ LANTUS 8 U S/C HS OD

HYPOGLYCEMIA EDUCATION DONE. REGULAR RBS MONITORING ADVISED.

INJ. ALBUMIN 20% 100 ML IV ONCE A WEEK UNDER MEDICAL SUPERVISION OVER 4-6 HOURS

**REPEAT KFT AFTER 3 DAYS AND INFORM**

VOPD AFTER ONE WEEK WITH CBC/LFT/KFT REPORTS

PLAN: TITRATE DIURETICS; BETA BOCKERS

UGIE + ENDOTHERAPY AFTER 3 WEEKS

REVIEW IN HEPATOLOGY OPD WITH REPORTS OF CBC/LFT/KFT/INR AFTER 4 WEEKS.

### Summary 10

**S/P DDLT** (Split Right Lobe Graft) for Cirrhosis - HCV Related (30/05/2024)

- Right lobe without MHV
- V8 + V5 reconstructed with cadaveric iliac vein, anastomosed to RHV to form single venous outflow
- Single RPV, Single RHA, Single RHD
- PV eversion thrombectomy done
- GRWR - 1.95 (1136gm)

**Post Op Sequelae:**

- Seizure episode POD#10 🡪 Left parietal ICH
- GNB sepsis, pleural effusion and AKI (blood c/s + urine c/s- K. Pneumoniae)
- Hematochezia from rectal ulcer🡪 Sigmoidoscopy guided ADR Endotherapy done
- ACR with Persistent Transaminitis –
  - Liver graft biopsy, 17/10/2024 - Mild ACR with co-existent biliary pathology. RAI- 4/9
  - S/P Steroid pulse therapy- Inj. Methylprednisolone 250mg OD (day 1 to day 3) 🡪 tapered to Wysolone 15mg OD

**Immunosuppression History:**

- Inj. Methylprednisolone 16mg OD (POD#1 to POD#5)
- Tab Wysolone 15mg OD
- Tacrolimus 0.5mg BD (31/05/2024 till 20/07/2024)
  - TAC level lowest 2.2ng.ml (POD#22) | highest 13.2ng/ml (POD#13)--> Discontinued I/v/o TAC toxicity (ICH-left parietal)
- MMF 500mg (POD#6 onwards) 🡪 Discontinued
- Cyclosporine (20/07/2024 onwards) 🡪 Currently on Cyclosporine 100mg BD -S.Cyclosporine Levels: 201 on 09/11/2024
- Everolimus 0.5 mg BD 🡪 1 mg BD

**Current issues:**

- Focal Seziures
  - EEG normal, Anti-epileptic dose optimized 🡪 Resolved
- Transaminitis 🡪 Immunosuppresants optimized

**Co morbidity -** Osteoporosis | Cervical spondilytis | NODAT

**PRESENTING COMPLAINTS:**

- Right Facial Twitching

**INDICATION FOR ADMISSION:**

For further evaluation and management of presenting complaints

**HISTORY:**

Mrs. Patient J is 61 years old female with prior co morbidities of osteoporosis/Cervical spondylitis. Patient had her index presentation in 2016 when she presented with abdominal distension. On further evaluation she was found to be HCV reactive. Further assessment showed presence of cirrhosis. She has been on OPD follow up thereafter. In view of hepatic hydrothorax, refilling ascites, Coagulopathy she was advised for DDLT. Patient got registered and underwent the same on 30/05/2024. Post procedure sequelae has been mentioned above. Follow up investigations have shown gradual elevation in AST/ALT levels, and she was admitted on October 2024 with ACR. USG guided percutaneous biopsy done – 17/10/2024 reported Mild ACR with co-existent biliary pathology. RAI- 4/9. She has currently presented with an intermittent right facial twitching. No h/o fever, pain abdomen, hematemesis, melena, jaundice, alteration of bowel habits, burning micturition. There is no h/o of hereditary/ familial diseases/ T2DM/ HTN/ CAD/ thyroid/ renal/ no major surgery done in the past.

**EXAMINATION:**

Pt. was conscious, oriented to time place and person.

BP: - 110/60mm Hg, Pulse: - 92/min, RR: - 24/min and afebrile.

Pallor-, Icterus-, Cyanosis-, clubbing -, Pedal edema -JVP normal ,Tremor-

On systemic examination

Respiratory system:--Normal breath sounds present over bilateral air fields. No added sounds

Cardiovascular system: - S1 S2 normal, no murmurs

CNS: - conscious, oriented, no sensori-motor deficit, right sided facial twitching present

? Tics

Per Abdomen examination

ON INSPECTION: - Not distended, umbilicus central and inverted, skin over the abdomen is not stretched with no visible venous prominences, no visible pulsations. Surgical scar of previous intervention observed

ON PALPATION: - soft, non-tender, liver and spleen non palpable. No guarding, no rigidity, no rebound tenderness.

ON PERCUSSION: - Tympanic note, no free fluid

ON AUSCULTATION normal bowel sounds present.

**SYSTEMATIC REVIEW:**

Patient was admitted with above mentioned complaints. Her examination findings were as mentioned above. Her initial lab data has been mentioned below. Sepsis screening was done. Urine routine showed normal study. In view of facial tics. neurology consults were taken and EEG was done – Normal study. Anti-epileptics modified accordingly and facial twitching resolved. In view of transaminitis, immunosuppressants were modified and cyclosporine dose titrated to 100 MG BD, and Everolimus increased from 0.5 mg BD to 1 mg BD. Following that, she improved symptomatically. She is now planned for OPD follow up with response assessment. The nature of disease and its prognosis has been explained to patient’s attendants in full details. She has been managed conservatively and is now being discharged in stable state to be followed up on OPD basis.

**ADVICE AT DISCHARGE (Duration of medications if any):**

1800 KCAL/DAY, 70 GRAMS PROTEIN/DAY, NORMAL DIABETIC DIET AS ADVISED

TO PRACTICE STRICT SOCIAL DISTANCING AND GOOD PERSONAL HYGIENE WHILE ON STEROIDS/ IMMUNOSUPPRESSANT.

ANY FEVER, BURNING MICTURITION, COUGH, SOB, JOINT PAIN IS TO BE NOTIFIED IMMEDIATELY.

REGULAR PHYSICAL ACTIVITY OF MINIMUM 30 MINUTES TO BE INCLUDED INTO ROUTINE

TAB WYSOLONE 15 MG PO OD

TAB EVEROLIMUS 1 MG PO BD

TAB CYCLOSPORINE 100 MG PO BD

TAB LEVEPIL 500 MG MORNIING AND 750 MG EVENING

TAB NICARDIA 10 MG PO BD

CAP HENZOVIT 1 TAB PO OD

TAB URSOCOL 450 MG PO BD

TAB GABAPIN 100 MG PO OD HS

FOLLOW UP IN HEPATOLOGY OPD AFTER 1 WEEK WITH CBC/KFT/INR/LFT REPORTS

PLAN: OPTIMIZATION OF IMMUNOSUPRESSANTS ON OPD BASIS

IN CASE OF DECREASE IN URINE OUTPUT, ALTERED SENSORIUM, BLEEDING

### Summary 11

**LAMA SUMMARY**

PHTN – Non- Bleeder, Grade II Esophageal Varices

Cirrhosis – Metabolic (K74.6) + Ethanol (K70.9) (LI –1.5 years ago)

Decompensated – Ascites / Jaundice / AKI

CTP – 12 C	MELD Na – 36

**Explained about need of LT.**

**CURRENT ISSUES:-**

- Ascites – Grade III
- AKI – KDIGO III Non- oliguric (? Diuretic Induced + Sepsis - Creatinine 3.60 🡪 2.90)

**CHIEF COMPLAINTS:-**

Worsening Abdominal Distension

Generalised Swelling

**INDICATION FOR ADMISSION:**

For evaluation and management of presenting complains

**HISTORY:**

Mr Patient K is a 48 year old gentleman with no prior comorbidities. He had his index presentation in 2021 when he developed abdominal distension. This was associated with a loss of appetite and swelling of lower limbs. He was diagnosed at an outside centre as cirrhosis and was initiated on diuretics. In April 2024, he developed a yellowish discolouration of his eyes and urine, which was insidious in onset and progressive. It was not associated with itching or clay-coloured stools. Over the past 1 month, his abdominal distension has increased. He has also developed a progressive swelling over his whole body. There is no h/o burning micturition or decreased urine output. There is no h/o major surgeries or IV drug abuse prior to onset of the disease. There is history of CAM intake. There are no h/o CAD/HTN/COPD/ TB/Thyroid or renal disorders.

**GENERAL EXAMINATION:**

Pt. was conscious, oriented to time place and person, and afebrile.

BP- 100/64 mm Hg, Pulse- 88/min, RR- 20/min and afebrile.

Pallor+, Icterus+, Cyanosis-, Clubbing-, Pedal edema +, LNP-, JVP raised

On systemic examination

Respiratory system: - B/L airway entry present, basal crepts present bilaterally

Cardiovascular system: - S1 S2 normal, no murmurs

CNS: - conscious, oriented with no sensorimotor deficit,

Per Abdomen examination

ON INSPECTION: - distended abdomen, umbilicus central and inverted, skin over the abdomen is stretched with no visible venous prominences, no visible pulsations.

ON PALPATION: - soft, non-tender, no palpable organomegaly. No localized rise of temperature observed over the swelling region.

ON PERCUSSION: dull note present in all quadrants, free fluid present.

ON AUSCULTATION-Normal bowel sounds present.

**SYSTEMATIC REVIEW:**

Patient was admitted with above mentioned complaints. His examination findings were as mentioned above. S. PCT – 0.16. Ascitic fluid tap was done which revealed a straw coloured fluid with WBC - 69, N – 27% and L – 72%, Protein: 0.97, Glu: 107, Albumin: 0.41, SAAG: 2.19, ADA: 4.1 and Gene Xpert: negative, Gram Stain negative. Cultures were negative. He was started on IV antibiotics, IV diuretics. Blood products were transfused as required. The nature of the disease and long term prognosis was explained in great detail to the patient and the patient relatives. The need for urgent Liver Transplant in view of a Decompensated Liver Disease was explained in great detail, along with the risks and benefits of the same, however they do not have a donor at present. He is now planned for nutritional optimization and improvement in performance status. Proper diet and mobilisation was followed throughout the course of admission. All vitals, temperature, RBS and other necessary parameters were checked regularly and managed appropriately. He is currently being **discharged against medical advice**, with the advice to follow up at a tertiary centre with Hepatology and Liver Transplant services.

**ONGOING MEDICATIONS:**

1800 KCAL/DAY + 90 GRAMS PROTEIN/DAY, LOW SALT DIET

INJ PIPTAZ 4.5 GM IV TDS

T. RIFAGUT 550 MG PO BD	1-0-1

SYP LACTIHEP 30ML PO HS  0-0-1 (ENSURE 2-3 BOWEL MOTIONS/DAY)

HEPSURE SACHET PO BD

TAB ME-12 1 TAB PO OD

TAB ZINC PO OD

TAB TRENTAL 400 MG PO OD

CAP HENZOVIT 1 TAB PO OD

TO FOLLOW UP AT A TERTIARY CENTRE WITH HEPATOLOGY AND LIVER TRANSPLANT SERVICES

IN CASE OF DECREASE IN URINE OUTPUT, ALTERED SENSORIUM, BLEEDING

### Summary 12

**1.)** Portal hypertension - (Non bleeder, Grade II esophageal varices without RCS, mild PHG, Early GAVE, duodenal ulcer, S/P-EVL(28/05/2021), HVPG - 14mm of Hg - 2021)

Chronic liver disease (DoI – 8 years) **(ICD10 – K75.8)**

Prior ACLF – 2017 – s/p steroids

Currently Recompensated

Prior Decompensated with UGI bleed, Prior Ascites and Jaundice

MELD Na -17        CTP- 9	CHILD – B

**2.)** Carcinoma of Unknown Primary (T0 N3b N1) – Biopsy – Squamous Cell Carcinoma

- DoI – 1.5 years
- With Bilateral Cervical LN, Sternal Metastasis
- Left Cervical Lymph node – Biopsy Proven
- s/p NACT – 3 cycles TP (Paclitaxel + Carboplatin) regimen (LD – 06/05/2024)
- s/p Concurrent CT-RT – 4 cycles CDOP regimen + RT (03/04/2024 – 28/05/2024)
- s/p 7 cycles of TP (Paclitaxel + Carboplatin) + 4 cycles of Immunotherapy - Nivolumab last received on 08/02/2025
- PET-CT – 28/02/2025 – Progressive Disease

**COMORBIDITIES:**

- T2DM
- Hypothyroidism (TSH-10)
- Psoriasis (h/o steroid intake)
- h/o COVID 19 pneumonia: Severe Category (Recovered on 31/05/2021)
- h/o GTCS – 1 episode on 11/02/2025 – on Levera
- Cholelithiasis with Choledocholithiasis

**CURRENT ISSUE**:

- Sepsis with Septic Shock –
  - Bilateral Pneumonia - Sputum C/S – Klebsiella pneumoniae
- Sepsis Related Worsening of Jaundice (TB – 6.93 🡪 2.93)
- Carcinoma of Unknown Primary – Squamous Cell Carcinoma 🡪 PET-CT on 28/02/2025 - Progressive Disease

**PRESENTING COMPLAINTS:**

Abdominal Pain x 7 days

Fever x 1 day

**INDICATION FOR ADMISSION:**

For further evaluation and management

**HISTORY:**

Mr Patient L is a 52 years old gentleman, with prior known co-morbidity of T2DM and hypothyroidism. His index presentation was in October 2017 when he developed jaundice and ascites with pedal edema, for which he was evaluated outside and diagnosed as a case of chronic liver disease. Later he was admitted in ILBS on 13/12/2017 and was managed conservatively as ACLF with steroid course, after which he was discharged on 16/12/2017. He was admitted at ILBS from 27/05/21 to 02/06/21 with complaints of fever, generalised weakness and was detected with COVID 19 and managed conservatively and subsequently discharged in hemodynamically stable condition. He consumed steroids in tapered doses for 1 month i/v/o severe disease – COVID related . He then presented at an outside centre where he was revealed to have a left lymph node. Biopsy was done outside revealed – SCC. Imaging revealed Bilateral Cervical LN with Sternal Metastasis. He received NACT – 3 cycles TP (Paclitaxel + Carboplatin) regimen (LD – 06/05/2024), followed by Concurrent CT-RT – 4 cycles CDOP regimen + RT (03/04/2024 – 28/05/2024). PET-CT showed resolution of left sided nodes, but new metabolically active right sided lymph nodes. He then underwent 7 cycles of TP (Paclitaxel + Carboplatin) + 4 cycles of Immunotherapy - Nivolumab last received on 08/02/2025. He was recently admitted at an outside centre with abdominal pain, fever and vomiting. He was managed conservatively as acute cholangitis secondary to passed off CBD stone and was discharged in satisfactory condition. However, his fever recurred and he presented to ILBS for further management of the same. There is no h/o vomiting, hematemesis, altered bowel habits or decreased urine output. There is no h/o CAM intake, indigenous medication in the past. There is no h/o HTN/COPD/CAD/CVA/TB

**EXAMINATION:**

BP- 80/60 mm of Hg, Pulse- 94/min, RR- 20/min, febrile

Pallor-, Icterus-, Clubbing-, Cyanosis-, Pedal edema-, LNP-, JVP normal

CVS SI, S2 normally heard, no murmurs

CHEST - B/L airway equal air entry, no wheeze, B/L occasional crepts

CNS - Patient conscious and oriented, No sensorimotor deficit.

P/A - ON INSPECTION scaphoid, umbilicus central and inverted, no visible venous prominences, no visible pulsations

ON PALPATION - soft, non tender, liver and spleen not palpable, no guarding, no rebound tenderness

ON PERCUSSION- Tympanic note, free fluid absent

ON AUSCULTATION - Normal bowel sounds present

**SYSTEMATIC REVIEW:**

Patient was admitted with the above mentioned complaints. His examination findings were as mentioned above. Patient was admitted with above mentioned complaints and baseline investigations were sent. His examination findings were as mentioned above. His initial lab data and latest lab data is included at the end of the summary. He was shifted to the HDU in view of sepsis with septic shock. Sepsis protocol was initiated and he was started on inotropes and fluid resuscitation. Blood and Urine Cultures revealed no growth, and Urine Routine was unremarkable. S.PCT – 6.59. He was started on empirical IV antibiotics and IV antifungals. CXR revealed bilateral pneumonia. Sputum c/s revealed Klebsiella pneumonia and antibiotics upgraded accordingly. He improved clinically and was shifted to the ward. Blood products were transfused as required and he gradually improved. Incentive spirometry, chest physiotherapy, nebulisations and supportive management continued. Oxygen support was gradually tapered. Medical Oncology opinion was sought in view of SCC with unknown primary. PET-CT was done on 27/02/2025 revealed –

- Metabolically active large conglomerated heterogeneously enhancing mass in the left cervical region as detailed above with other discrete cervical lymph nodes-residual disease.
- Features of CLD with portal hypertension (splenomegaly, dilated s/p axis and abdominal collaterals).
- No evidence of abnormal metabolically active or arterial enhancing lesions noted in the liver parenchyma.
- Multiple patchy consolidative changes with increased metabolic activity involving both lungs (left >>right) -likely infective.
- Mild metabolic activity in few mediastinal lymph nodes - part of same disease.
- Non-FDG avid lytic lesion involving the manubrium sternum -resolved disease.

As compared to previous FDG PET CT scan dated 12-09-2024,

- There is significant interval increase in size of the conglomerated mass involving the left cervical region with appearance of new discrete cervical lymph nodes -suggestive of progressive disease.
- Interval appearance of infective changes in both lungs.
- Interval appearance of minimal free fluid in the pelvic cavity.

Proper diet and mobilisation was followed throughout the course of admission. All vitals, temperature, RBS and other necessary parameters were checked regularly and managed appropriately. He recovered symptomatically following the management and is discharged in hemodynamically stable condition with following advice to follow in OPD.

**PLAN / ADVICE AT DISCHARGE (Including duration of medication if any):**

2000 KCAL/DAY + 90 GRAMS PROTEIN/DAY, LOW SALT (<2GRAMS/DAY), LOW FAT DIET

INJ ELORES 1.5 GM IV TWICE DAILY FOR 8 DAYS

TAB FLUCOS 200 MG PO OD FOR 5 DAYS

TAB MIDODRINE 5 MG PO TDS

TAB UDCA 450 MG PO BD

TAB LEVEPIL 500 MG PO BD

CAP HENZOVIT 1 TAB PO OD

TAB SHELCAL-M 1 TAB PO OD

SYP LACTIHEP 30ML PO HS  0-0-1 (ENSURE 2-3 BOWEL MOTIONS/DAY)

LAXOPEG SACHET SOS (ENSURE 2-3 BOWEL MOTIONS/DAY)

TAB ULTRACET ½ TAB PO SOS FOR PAIN

TAB THYRONORM 125 MCG PO BBF

REVIEW IN HEPATOLOGY OPD WITH CBC/LFT/KFT/INR REPORTS AFTER 4 WEEKS.

REVIEW IN MEDICAL ONCOLOGY OPD WITH CBC/LFT/KFT/INR REPORTS AFTER 1 WEEKS.

REPEAT CBC, LFT, KFT, INR AFTER 1 WEEK AND INFORM IN VIRTUAL OPD

IN CASE OF DECREASE IN URINE OUTPUT, ALTERED SENSORIUM, BLEEDING, FEVER, NEW ONSET COUGH AND REVIEW IN ILBS EMERGENCY ON SOS BASIS.

### Summary 13

PHTN - Bleeder, Eradicated Esophageal Varices, Antral GAVE - 26/07/2023

CLD - HCV

Decompensated with Ascites / Right hydrothorax

HCC Segment IVA with Peritoneal Metastasis

PVT - left branch portal vein with extension into the main portal vein (non-tumoral)

S/P SBRT 5 Cycles (Feb 2023)

Lenvatinib intolerant

PET-CT – 06/11/24 - suggestive of progressive disease with omental, ovarian and pleural deposits.

Omental Biopsy – 05/11/2024 – suggestive of metastatic HCC

Comorbidities - None

**(ICD 10 - C22.0)**

**Current issues:**

- Diuretic Intractable Ascites (High SAAG, Low Protein, No SBP) s/p PCD inserted on 05/11/2024 🡪 discharged with PCD in situ
- Dyselectrolemia: Hyponatremia

**CHIEF COMPLAINTS**

- Abdominal Distension x 1 month
- Generalised Weakness

**INDICATION FOR ADMISSION:**

For further evaluation and management

**HISTORY:**

Mrs Patient M is a 57 years old lady with no known comorbidities. She had her index presentation in 2013 when she was evaluated for loose stools and was diagnosed with CLD- HCV related. She was started on dual antivirals and was on regular follow up. She was admitted at ILBS in January 2023 with complaints of pain in abdomen for which she was evaluated and underwent PET Scan, which showed left branch portal vein with extension into the portal vein. Metabolically active ill-defined hypodense space-occupying mass lesion noted in segment IVA of liver with significant enhancement in the late arterial phase and washout in the delayed phase LIRADS V-features are in favour of HCC and she underwent FNA from liver SOL on 7/1/23 which showed atypical cells present. She subsequently underwent SBRT 5 cycles in February 2023 and was started on TKI Lenvatinib which was stopped as she was intolerant. She was recently admitted with a fever on and off since 2 months with an evening rise of temperature. She was admitted outside and blood culture revealed a growth of Rhizobium radiobacter, and then transferred to ILBS. Sepsis controlled and discharged in hemodynamically stable condition. Currently, she has presented with worsening abdominal distension. There is no h/o hemoptysis, hematemesis, altered sensorium, burning micturition or decreased urine output. With these complaints she has come to ILBS and got admitted for further evaluation and management. There is no h/o any intoxications, indigenous medications, blood transfusions or IV drug abuse prior to onset of the disease. There is no h/o DM/HTN/CAD/TB/COPD/Thyroid disorders.

**EXAMINATION:**

Pt. was conscious, oriented to TPP, afebrile.

BP -124/70 mm Hg , Pulse -110/min	RR -20 /min

Pallor+, Icterus-, Cyanosis-, Clubbing-, Pedal edema- LNP-, JVP normal

CVS -S1 S2 normal, no murmurs

Chest- B/L normal air entry & vesicular breathing, no adventitious sounds

CNS - conscious, Oriented, no focal neurological deficit.

P/A ON INSPECTION- distended Abdomen, umbilicus central and inverted, no visible venous prominences, no visible pulsations.

ON PALPATION- soft, non-tender, liver - palpable 2 cm BCM and Spleen - non palpable , No guarding, no rigidity, no rebound tenderness.

ON PERCUSSION- shifting dullness present

ON AUSCULTATION- normal bowel sounds present, no bruits.

**SYSTEMATIC REVIEW:**

Patient was admitted with the above mentioned complaints. Her examination findings were as mentioned above. Ascitic fluid tap was done which revealed a straw coloured fluid with WBC - 99, N 11% and L 88%, Protein: 1.02, Glu: 106.2, Albumin: 0.69, SAAG: 2.52, ADA: 5.2 and Gene Xpert: Negative, Gram stain showed pus cells. Cultures were negative. Cytology was negative for malignancy. PCD was inserted on 05/11/2024 and daily 1-1.5 L drained. Omental biopsy was done on 05/11/2024 – suggestive of metastatic hepatolocellular carcinoma. Gynecology opinion was sought in view of bilateral bulky ovaries (primary malignancy) and PET-CT advised.

PET-CT was done on 06/11/2024 –

- Features of CLD with portal hypertension (splenomegaly, dilated s/p Axis, abdominal collaterals and ascites) as detailed above.
- Faint metabolic activity in an ill-defined hypodense lesion noted in segment IVA of liver with no significant arterial enhancement –? Residual disease cannot be ruled out.
- Non-FDG avid thrombus involving the left branch portal vein with extension into the main portal vein -likely bland thrombus.
- Faint metabolic activity in multiple omental and peritoneal nodules along with enlarged para-aortic, aortocaval and bilateral external iliac lymph nodes-likely metastases.
- Bilateral mild pleural effusion with multiple irregular pleural-based deposits -likely metastases.
- Bilateral bulky ovaries with heterogeneous enhancement and mildly increased metabolic activity -likely secondary deposits
- As compared to previous scan dated 13-08-2024;
   -	There is interval appearance of multiple pleural-based deposits in both lungs, multiple omental and peritoneal nodules in the abdominal and pelvic cavity, enlarged lymph nodes and para-aortic, aortocaval and bilateral external iliac regions and bilateral ovarian deposits -suggestive of progressive disease.
  -	There is interval increase in volume of bilateral mild pleural effusion.
  -	Interval appearance of fracture and left second rib anteriorly.

Patient was treated with IV antibiotics, IV albumin, nutritional support and other supportive medication and treatment. Medical Oncology opinion sought and advice followed. Proper diet and mobilisation was followed throughout the course of admission.patient was planned for immunotherapy for HCC but they requested for time to decide for the same. They were initiated on TKI therapy. All vitals, temperature, RBS and other necessary parameters were checked regularly and managed appropriately. The nature of the disease and its long term prognosis was explained in great detail to the patient and the patient attendant also explained in detail about the limited treatment options available. Now she is being discharged in hemodynamically stable state on patients request with the advice to follow up in Hepatology and medical oncology OPD.

**PLAN / ADVICE AT DISCHARGE (Including duration of medication if any):**

2100 KCAL/DAY + 90 GRAMS PROTEIN/DAY, NORMAL DIET

PCD CARE AS ADVISED

TAB TAXIM-O 200MG BD	1-0-1 X 5 DAYS

T. RIFAGUT 550 MG PO BD	1-0-1 (START ON 6TH DAY)

TAB CABOZANTINIB 40 MG PO OD (PLAN DOSE OPTIMISATION ON OPD BASIS)

CAP HENZOVIT 1 TAB PO OD

TAB ULTRACET 1 TAB SOS (FOR PAIN)

PROHANCE HP POWDER 2 SCOOP PO TDS

HEPSURE SACHET PO BD

BENZ PEARLS PO BD

INJ. ALBUMIN 20% 100 ML IV ALTERNATE DAY UNDER MEDICAL SUPERVISION OVER 4-6 HOURS

Review in Hepatology OPD and medical Oncology OPD after 1 week with CBC, LFT, KFT, INR

Plan : Half dose Immunotherapy for Metastatic HCC

IN CASE OF DECREASE IN URINE OUTPUT, ALTERED SENSORIUM, BLEEDING, FEVER, NEW ONSET COUGH AND REVIEW IN ILBS EMERGENCY ON SOS BASIS.

### Summary 14

PHTN - Non Bleeder, Small High Risk Esophageal Varices S/P EVL - September 2024, outside

Cirrhosis- HBV (HBV DNA – 1.15 log2 | HBeAg-R | Anti HBe-NR | HBsAg - 5260) + Superadded HDV (HDV RNA - 2.68 log 4)

S/P PLEX – 4 sessions – Normal Volume on 28/01/2025, 29/01/2025, 30/01/2025 and High Volume on 01/02/2025

Decompensated- Jaundice | Ascites | Prior HE

Hepatocellular Carcinoma (ICD – C22.0)

- Segment VIII – 13 mm LR4 lesion

CTP-10	Child-B	MELD Na-25

LT Counselled as best option.

**ICD-B19.2/K75.8 / C22.0**

**CO MORBIDITY -** External haemorrhoids

**CHIEF COMPLAINTS**

- Worsening jaundice
- Generalized weakness and loss of appetite

**CURRENT ISSUES:-**

- HBV + HDV Superinfection
  - S/P PLEX – 4 sessions – Normal Volume on 28/01/2025, 29/01/2025, 30/01/2025 and High Volume on 01/02/2025
- HCC - Segment VIII – 13 mm LR4 lesion

**INDICATION FOR ADMISSION:**

Evaluation and Management

**HISTORY:**

Mr. Patient N is a 41 years old male patient with no prior co morbidity. He had his index presentation in 2010 in the form of jaundice painless, with yellowish discoloration of sclera and urine, insidious onset, gradually progressive, not associated with pruritus or clay-colored stools. This was also presented with prodrome of nausea and malaise. However he was only managed conservatively and no follow up was done. In 2020 during blood donation he was incidentally screened to be HBV +. In 2022 patient developed swelling over both legs with abdominal distension which was insidious in onset, gradually progressive, painless, non-tender, not accompanied with pedal edema, and not associated with nausea, vomiting, colicky pain, constipation, obstipation, fever, diarrhea, passage of mucous or blood per rectum. He was initiated on oral diuretics for the same and responded well. In 2023 patient had an episode of bleeding per rectum from haemorrhoid for which he was on homeopathic medications (CAM+). In June 2023 he had an episode of altered sensorium precipitated by constipation which was managed conservatively. Currently, he has been admitted with a worsening jaundice. There is no h/o fever, vomiting, cough, altered bowel habits, hematemesis, and malena, burning micturition, altered sensorium or decreased urine output. There is no h/o any intoxications, indigenous medications, major surgeries, blood transfusions or IV drug abuse prior to onset of the disease. There are no h/o DM/HTN/CAD/TB/COPD/Thyroid disorders.

**EXAMINATION**

Pt. was conscious, oriented to time place and person.

BP: 116/70 mmHg, HR- 86/min, RR- 14/min and afebrile.

Pallor-, Icterus+, Cyanosis-, Clubbing-, Pedal edema (pitting type)+ , LNP-, JVP normal

On systemic examination

Respiratory system:--B/L vesicular breathing, B/L airway equal air entry, no wheeze, no crepts

Cardiovascular system: - S1 S2 normal, no murmurs

CNS: - conscious and oriented with no sensorimotor deficit,

Per Abdomen examination

ON INSPECTION: Distended abdomen, umbilicus central and inverted, skin over the abdomen is stretched with no visible venous prominences, no visible pulsations.

ON PALPATION: soft, non-tender, liver and spleen non- palpable below coastal margins. No guarding, no rigidity, no rebound tenderness.

ON PERCUSSION: Dull note over all quadrants with free fluid present

ON AUSCULTATION: normal bowel sounds present,

**SYSTEMIC REVIEW:**

Patient was admitted with above mentioned complaints. His examination findings were as mentioned above. His initial lab data has been mentioned at the end of this case summary. Sepsis screening was done. Blood and urine cultures showed no growth. Urine routine showed normal study. 4 sessions of PLEX was done. Nutritional optimization has been done and patient is tolerating orally well. Now he is planned for discharge and follow up on OPD basis. Patient was treated with IV antibiotics, IV albumin, nutritional support and other supportive medication and treatment. Proper diet, purging and mobilization was followed throughout the course of admission. All vitals, temperature, RBS and other necessary parameters were checked regularly and managed appropriately. The nature of the disease and long term prognosis was explained in great detail to the patient and the patient’s relatives. Family was also explained regarding the need of liver transplant as a definite option of management. He has recovered symptomatically following the management and is being discharged with advice to follow up on OPD basis.

**PLAN / ADVICE AT DISCHARGE (Including duration of medication if any):**

·         1800 KCAL/DAY + 80 GRAMS PROTEIN/DAY, LOW SALT (<2GRAMS/DAY), NORMAL DIET AS ADVISED

·         WEIGHT REDUCTION WITH DAILY 30 MIN EXERCISE TO BE INCLUDED INTO ROUTINE

·         ENSURE 2-3 BOWEL MOVEMENTS/DAY AND AVOID CONSTIPATION WITH GOOD HYDRATION

TAB TENOFOVIR ALFENAMIDE 25MG PO OD 1-0-0

TAB ENTACAVIR 0.5MG PO OD 1-0-0

TAB UDCA 450MG PO BD 1-0-1

TAB TRENTAL 400MG PO OD 1-0-0 (WITH MEALS)

TAB HEPKART PO BD 1-0-1

TAB CARDIVAS 3.125MG PO BD 1-0-1 (STOP IF HR<55/MIN OR BP<90/50MMHG)

TAB DOM DT 10MG PO SOS (IN CASE OF NAUSEA)

MOVICOL SACHET PO SOS (IN CASE OF CONSTIPATION)

CAP. LUMIA D3 60K IU PO OD 1-0-0 FOR 7 DAYS THEN ONCE A WEEK FOR 4 WEEKS THEN STOP

NURAYELD HP POWDER 2 SCOOPS PO BD 1-0-1

SYP LACTIFIBER 15ML PO HS 0-0-1 (ENSURE 2-3 BOWEL MOTIONS/DAY)

REVIEW IN HEPATOLOGY OPD WITH CBC/LFT/KFT/INR REPORTS AFTER 2 WEEKS

PLAN: PEG-INTERFERON VS BULEVIRTIDE

ER SOS

### Summary 15

Portal Hypertension - (Bleeder, Residual Non-Bandable Varices, Mild PHG, Obturated GOV, GAVE – APC done – 14/02/2025)

CLD - Cryptogenic

Decompensated with Ascites  / Recurrent AVB

TIPS done on 23/11/2023 for Recurrent Variceal Bleeding.

S/P TIPS Venoplasty done on 23/01/2023

**CURRENT ISSUES**:

- TIPSS Stent Block - S/P TIPS Venoplasty done on 02/05/2025
- Post procedure – Right SCM Haematoma 🡪 Improved
- SOL Liver LR4 - 2x2.2 cm Segment VIII

AFP – 3.3

PIVKA – 339.76

**CO-MORBIDITIES:**

- T2 –DM
- Sarcopenia
- IDA
- s/p modified ATT from June 2022 for abdominal kochs
- h/o left inguinal hernia operation

**PRESENTING COMPLAINT**

- Worsening Abdominal Distension x 3 month

**INDICATION FOR ADMISSION:**

Evaluation and the management of the symptoms

**HISTORY:**

Mr. Patient O is a 61 years old male, with known case of T2DM, Sarcopenia and no history of any addiction. He had his index presentation on 2021 march in the form of haematuria and melena on evaluation it’s found to have CLD. Admitted in outside hospital treatment taken. TIPS procedure was done on 23/11/22 after cardiac clearance, under strict aseptic precautions right internal jugular vein was accessed using standard angiographic technique and a 10 F vascular access sheath is placed, no post procedure complication. Three months back, he was operated for a left inguinal hernia, following which he noticed a worsening abdominal distension. Currently, he is admitted for the same. There is no recent h/o vomiting, altered bowel habits, abdominal distension abdominal pain, cough, pedal edema, burning micturition or decreased urine output. With these complaints he got admitted for further evaluation and management. There is no h/o any intoxications, indigenous medications, blood transfusions or IV drug abuse prior to onset of the disease.

**EXAMINATION:**

Pt. was conscious, oriented to time place and person.

BP: - 128/81 mm Hg, Pulse: - 54min, RR: - 18/min and afebrile.

Pallor-, Icterus-, Cyanosis-, Clubbing-, Pedal edema - , LNP-, JVP normal

On systemic examination .

Respiratory system:B/L vesicular breathing,B/L airway equal air entry, no wheeze, no crepts

Cardiovascular system: - S1 S2 normal, no murmurs

CNS: - conscious, oriented with no sensorimotor deficit,

Per Abdomen examination .

ON INSPECTION: - distended abdomen, umbilicus central and inverted, skin over the abdomen is normal with no visible venous prominences, no visible pulsations.

ON PALPATION: - soft, liver not palpable	Spleen	not palpable, No guarding, no rigidity

ON PERCUSSION: - dull note , free fluid present – Grade III Ascites

ON AUSCULTATION-normal bowel sounds present.

**SYSTEMATIC REVIEW:**

Patient was admitted with above mentioned complaints. His examination findings were as mentioned above. Blood and Urine Cultures revealed no growth, and Urine Routine was unremarkable. S. PCT – 0.13. PIVKA II – 339. USG screening was done to check for TIPS patency TIPS stent in situ; No flow seen with in the stent; Hypoechoic lesion in segment - 8 of liver; Moderate ascites and advised:- TIPS venogram +/- venoplasty. Cardiology clearance for the same obtained. Case discussed with IR team and TIPS Venogram + Angioplasty for TIPS Block was done on 02/05/2025. IR screening post procedure was done – TIPS stent in situ show air artefact within flow at PV and HV side appears normal. PSV ~88 cm/s
PV Show normal flow. PSV ~22 cm/s. He developed a swelling over the right neck post-procedure. USG screening revealed a focal hypoechoic hematoma measuring 1x1.5cm in right SCM, which was managed conservatively, following which he improved. Proper diet, purging and mobilization was followed throughout the course of admission. All vitals, temperature, RBS and other necessary parameters were checked regularly and managed appropriately. The nature of the disease and long term prognosis was explained in great detail to the patient and the patients relatives. He is being discharged in haemodynamically stable condition with advice to follow up on OPD basis.

**PLAN / ADVICE AT DISCHARGE (Including duration of medication if any):**

**1800 KCAL /DAY, 90 GM PROTEINS / DAY, SALT IN TAKE < 2 GM PER DAY, EXERCISE**

**ENSURE 2-3 BOWEL MOTIONS/DAY**

TAB TAXIM-O 200MG BD	1-0-1 X 5 DAYS

T. RIFAGUT 550 MG PO BD	1-0-1 (START ON 6TH DAY)

SYP DUPHALAC 10ML PO HS  0-0-1 (ENSURE 2-3 BOWEL MOTIONS/DAY)

LAXOPEG SACHET BD. 1-0-1 (**ENSURE 2-3 BOWEL MOTIONS/DAY**)

LORNET SACHET PO BD

CAP HENZOVIT 1 TAB PO OD

NUTRIX ULTRA WHEY POWDER 2 SCOOPS PO TDS WITH MILK 1- 1-1-1—2PC

EFEZAC SACHETS PO BD

INJ. ALBUMIN 20% 100 ML IV ONCE A WEEK UNDER MEDICAL SUPERVISION OVER 4-6 HOURS

REVIEW IN HEPATOLOGY OPD WITH CBC/LFT/KFT/INR/USG SCREENING FOR ASCITES REPORT AFTER 2 WEEKS

### Summary 16

Portal HTN – Bleeder, Eradicated Esophageal Varices – 14/02/2025

Cirrhosis – Met ALD (LI- 1 year ago)

Decompensated - Jaundice | Ascites | Prior HE | AVB

CTP – 7	Child – B	MELD Na - 9

Right sided hydrothorax

- S/P STA (High SPAG, Low protein, No SBE), Diuretic intolerant
- STA done 4 times till date

**CURRENT ISSUES:**

- Right-sided Hydrothorax 🡪 S/P STA (High SPAG, Low protein, No SBE)
- Ascites – Grade III – (High SAAG, Low Protein, No SBP) - s/p STA – 3 L, Albumin Infusions and Nutritional therapy 🡪 Diuretics Optimized

**CO-MORBIDITY-**

- CAD S/P PTCA - 2010

**ICD-K75.6/70.9, I25**

**CHIEF COMPLAINTS**

Shortness of breath

**INDICATION FOR ADMISSION:**

Evaluation and the management of presenting symptoms

**HISTORY:**

Mr Patient P is a 56 years old gentleman with prior co morbidities of CAD s/p PTCA. He is a known ethanol user with LI- 1 year back. He had his index presentation in 2021 in the form of loss of appetite and shortness of breath. He was evaluated for the same and was incidentally diagnosed with CLD and pleural effusion. He had episodes of hematemtsis in January 2025 and EVL was done. Further evaluation showed hydrothorax for which he underwent STA. Till date 3 times tapping has been done, with a recent admission in February 2025 for the same. This time patient has presented with similar complaints of shortness of breath, insidious onset, gradually progressive, associated with dry cough and loss of appetite. With this clinical picture he has been admitted for further management. There is no h/o fever, vomiting, abdominal pain, altered bowel habits, hematemesis, and malena, burning micturition, altered sensorium or decreased urine output. There is no h/o any intoxications, indigenous medications, major surgeries, blood transfusions or IV drug abuse prior to onset of the disease. There is no h/o DM/ HTN/ TB/ COPD/ Thyroid disorders.

**EXAMINATION**

Pt. was conscious, oriented to time place and person.

BP: 106/70 mmHg, HR- 86/min, RR- 14/min and afebrile.

Pallor-, Icterus-, Cyanosis-, Clubbing-, Pedal edema (pitting type) - , LNP-, JVP normal

On systemic examination

Respiratory system:--Right sided decreased entry, no wheeze, occasional crepts

Cardiovascular system: - S1 S2 normal, no murmurs

CNS: - conscious and oriented with no sensorimotor deficit,

Per Abdomen examination

ON INSPECTION: distended abdomen, umbilicus central and inverted, skin over the abdomen is stretched with no visible venous prominences, no visible pulsations.

ON PALPATION: soft, non-tender, liver palpable 1 cm below right coastal margin. Spleen non- palpable below left coastal margin. No guarding, no rigidity, no rebound tenderness.

ON PERCUSSION: Dull note over all quadrants with free fluid present

ON AUSCULTATION: normal bowel sounds present,

**SYSTEMIC REVIEW:**

Patient was admitted with above mentioned complaints. His examination findings were as mentioned above. His initial lab data has been mentioned at the end of this case summary. Sepsis screening was done. Blood and Urine cultures showed no growth. Urine routine showed normal study. He was treated with IV antibiotics, IV albumin, nutritional support and other supportive medication and treatment. CXR showed right sided massive hydrothorax. STA of Ascitic Fluid – 3 L and Pleural Fluid – 1 L was done by Vascular Lab under USG guidance. Pleural fluid analysis was done which revealed a straw coloured fluid with WBC - 102, N – 14% and L – 85%, Protein: 1.51, Glu: 116, Albumin: 0.74, SPAG: 1.93, ADA: 6.9 and Gene Xpert: negative, Gram Stain negative. Cultures were negative. Proper diet, purging and mobilization was followed throughout the course of admission. All vitals, temperature, RBS and other necessary parameters were checked regularly and managed appropriately. Ascitic fluid analysis was done which revealed a straw coloured fluid with WBC - 172, N – 28% and L – 71%, Protein: 0.7, Glu: 100, Albumin: 0.3, SAAG: 2.26, ADA: 3.4 and Gene Xpert: negative, Gram Stain negative. Cultures were negative. The details of TIPSS as a bridge to transplant were explained. The nature of the disease and long term prognosis was explained in great detail to the patient and the patients relatives. Family was also explained regarding the need of liver transplant as a definite option of management. He has recovered symptomatically following the management and is being discharged with advice to follow up on OPD basis.

**PLAN / ADVICE AT DISCHARGE (Including duration of medication if any):**

1800 KCAL/DAY + 80 GRAMS PROTEIN/DAY, NORMAL DIET AS ADVISED

DAILY 30 MIN EXERCISE TO BE INCLUDED INTO ROUTINE

ENSURE 2-3 BOWEL MOVEMENTS/DAY

T. TAXIM 200MG PO BD 1-0-1 FOR 5 DAYS THEN STOP

TAB DYTOR 10 MG PO OD (MONITOR KFT WEEKLY)

TAB ALDACTONE 50 MG PO OD (MONITOR KFT WEEKLY)

SYP ZINCONIA 10 ML PO BD

TAB ECOSPRIN 75 MG PO OD

LORHEP SACHET PO BD

HEPSURE SACHET PO BD

SYP LACTIHEP 30ML PO HS  0-0-1 (ENSURE 2-3 BOWEL MOTIONS/DAY)

CAP HENZOVIT 1 TAB PO OD

REVIEW IN HEPATOLOGY OPD WITH CBC/LFT/KFT/INR REPORTS AFTER 4 WEEKS

KFT AFTER 1 WEEK AND INFORM

PLAN: - TITRATE THE DIURETIC DOSE AND FOLLOW UP

### Summary 17

Recurrent Acute Pancreatitis

Etiology: Biliary – Cholelithiasis with Choledocholithiasis

s/p ERCP + Stenting - 10/09/2024

s/p ERCP + Stent Exchange - 21/12/2024

s/p ERCP + Stent Exchange - 26/03/2025

s/p EUS Guided Liver Biopsy – Report Awaited

**CURRENT ISSUES:**

- Mild Acute Cholangitis (ICD 10 – K83.1)
  - S/P- ERCP + Stent Exchange - 26/03/2025

**CO MORBIDITIES -** None

**PRESENTING COMPLAINTS-**

Fever x 1 day after EUS guided live biopsy

**INDICATION FOR ADMISSION:**

Evaluation and management of presenting symptoms

**HISTORY:**

Mr. Patient Q is a 38 year old male, with no known comorbidities, presented with complaints of abdominal pain and jaundice in 2024. He was evaluated elsewhere and was found to have pancreatitis and he was referred to ILBS for the same. MRCP done on 04/09/2024 revealed a Cholelithiasis, choledocholithiasis with ? impacted periampullary calculus. IHBR irregularity with intraductal sludge / calculi and features of cholangitis. He had no history of trauma, drugs, alcohol prior to the presentation. ERCP and stenting was done on 10/09/2024 in ILBS, and stent exchange on 21/12/2024. The procedures were tolerated well and he was discharged in haemodynamically stable condition. Currently, he had undergone a EUS guided liver biopsy, following which he presented with a fever with chills. He has now been admitted for evaluation and management. There is no h/o jaundice, decreased urine output, cough, loose stools, altered sensorium, hematemesis, altered bowel movements. There were no indigenous medications intake or IV drug abuse history, alcohol intake, recent blood transfusion, IV drug therapy. There was no h/o DM/CAD/TB/asthma/Thyroid or renal disorder.

**EXAMINATION:**

Pt. was conscious, oriented and afebrile.

BP- 138/60mm Hg	Pulse- 76/min	RR- 22/min

Pallor -, Icterus-, Cyanosis-, Clubbing-, Pedal edema-, LNP-, JVP normal

CVS - S1 S2 normal, no murmurs

Chest - B/L normal air entry & vesicular breathing.

CNS- Patient is conscious and oriented with no focal neurological deficit.

P/A ON INSPECTION- non distended, umbilicus central and inverted, no surgical scar present, no visible venous prominences and no visible pulsations

ON PALPATION - soft, non- distended, epigastrium tenderness present - mild, liver not palpable and spleen not palpable. No guarding, no rebound tenderness.

ON PERCUSSION- Tympanic note over all quadrants, no free fluid present.

ON AUSCULTATION- normal bowel sounds present, no bruits.

**SYSTEMATIC REVIEW:**

Patient was admitted with the above mentioned complaints. His examination findings were as mentioned above. His initial lab data and latest lab data is at end of summary in chart form. Blood Cultures awaited. S. PCT – 3.64. USG Screening - GB is grossly distended with normal wall thickness. No IHBRD seen. Increased echogenicity seen around liver ducts ? cholangitis. He underwent ERCP with stent exchange on 26/03/2025. Patient tolerated the procedure well and post procedure period was uneventful. There were no further febrile episodes in ward. Nature of disease and its prognosis has been clearly explained to patient and his attendants in full details. He has been managed conservatively and is now being discharged in stable state to be followed up on OPD basis.

**PLAN / ADVICE AT DISCHARGE (Including duration of medication if any):**

1600 KCAL/ DAY, 60 GM PROTEIN/DAY, LOW FAT DIET

TAB TAXIM-O 200MG BD	1-0-1 X 5 DAYS

TAB PANTOCID 40 MG PO OD BBF

TAB EMESET 4 MG PO SOS FOR VOMITING

TAB UDCA 450 MG PO BD

TAB CROCIN 1 TAB PO SOS

T. ULTRACET PO SOS

TAB VIADEK PO OD

F/U IN VIRTUAL OPD AFTER ONE WEEK WITH CBC / LFT REPORTS

REVIEW IN HEPATOLOGY OPD WITH CBC/LFT/KFT/INR AFTER 6 WEEKS

IN CASE OF DECREASE IN URINE OUTPUT, ALTERED SENSORIUM, BLEEDING, FEVER, NEW ONSET COUGH AND REVIEW IN ILBS EMERGENCY ON SOS BASIS.

### Summary 18

PHTN - Non bleeder, Grade I Esophageal Varices, Mild PHG, Obturated IGV 1, Diffuse GAVE 04/01/2025

Cirrhosis with HCC   (**ICD: B 181)**

Decompensated: Ascites

HCC: Multifocal largest 4.1 x 3.1 cm in segment 8/4a of liver.

AFP: 1544	PIVKA 2: 1791

S/P STRIDE Regimen

-          TREMELIMUMAB - 300 mg (Single Dose Given on 22/01/2025)

-          DURVALUMAB - 1500 mg (Plan: 4 weekly cycles)

- s/p 2 sessions given on 22/01/2025 and 20/02/2025

MELD Na -22        CTP- 7 B	BCLC-C	PS- 0

**CO MORBIDITIES**- CKD Urine PCR: 4.91, HTN

**CURRENT ISSUES**-

-         Evaluation of liver SOL

**PRESENTING COMPLAINTS:**

For further evaluation and management CLD with HCC

**INDICATION FOR ADMISSION:**

For further evaluation and management CLD with HCC

**HISTORY:**

Mr. Patient R is a 58 years old male with known co morbidity of HTN and CKD diagnosed since 2024 January. He was being followed up for the same. He was recently diagnosed to have CLD with HCC while being evaluated for fever. PET CT was done elsewhere which showed FDG avid lesions predominantly in left lobe. He was admitted for further evaluation and management of CLD with HCC. He received his 1^st^ session of STRIDE Regimen on 22/01/2025. TREMELIMUMAB - 300 mg 22/01/2025 (Single Dose) was given, followed by DURVALUMAB - 1500 mg - 22/01/2025. He is currently admitted for second session of STRIDE. There is no h/o vomiting, altered sensorium, burning micturition or decreased urine output. There is no h/o any intoxications, indigenous medications, IV drug abuse prior to onset of the disease. There is no h/o thyroid disorders/CAD/COPD/genetic or hereditary disorders.

**GENERAL EXAMINATION:**

Pt. was conscious, oriented to TPP, febrile.

BP -116/78 mm Hg, Pulse -76/min;  RR -16/min

Pallor-, Icterus-, Cyanosis-, Clubbing-, Pedal enema - , JVP normal

CVS -S1 S2 normal, no murmurs

Chest- B/L normal air entry & vesicular breathing, no adventitious sounds

CNS - conscious, oriented, no focal neurological deficit

P/A ON INSPECTION- mildly distended, umbilicus central and inverted, no visible venous prominences, No visible pulsations.

ON PALPATION- soft, non-tender, liver just palpable BCM and spleen non palpable, No guarding, no rigidity, no rebound tenderness

ON PERCUSSION-dull note, free fluid present.

ON AUSCULTATION- normal bowel sounds present, no bruits.

**SYSTEMATIC REVIEW:**

Patient was admitted with above mentioned complaints. His examination findings were as mentioned above. His initial lab data and latest labs are at end of summary in chart form. HBsAg – Positive. He was admitted and initiated on nutrition and other supportive measures. Nephrology consult was taken, it was decided to proceed with immunotherapy with close watch on renal parametres. Sepsis ruled out. He received his 2^nd^ session of - DURVALUMAB - 1500 mg on 20/02/2025. No uneventful incident/adverse effect was noted. Nephro consult done in view of CKD. Patient was treated with nutritional support and other supportive medication and treatment. The nature of the disease and long term prognosis was explained in great detail to the patient and the patient relatives. He is now planned for nutritional optimization and improvement in performance status. Proper diet, purging and mobilization was followed throughout the course of admission. All vitals, temperature, RBS and other necessary parameters were checked regularly and managed appropriately. He recovered symptomatically following the management and is discharged in haemodynamically stable condition with following advice to follow in OPD.

**PLAN / ADVICE AT DISCHARGE (Including duration of medication if any):**

DIET AS ADVISED BY DIETICIAN (TARGET 1800KCAL/DAY, 70GM PROTEIN/DAY, SALT RESTRICTED)

HOME BP MONITORING

TAB TAXIM-O 200MG BD	1-0-1 X 5 DAYS

TAB TAF 25 MG OG

TAB UDCA 450 MG PO BD

TAB CILACAR 10 MG PO BD (DO NOT GIVE IF BP < 90/60 AND HR < 55)

TAB MOXINIDINE 0.3 MG BD (DO NOT GIVE IF HR < 55 OR BP < 90/60 MMHG)

TAB FEBUTAZ 40 MG HS

TAB PHOSCUT 400 MG OD

INJ. ALBUMIN 20% 100 ML IV ONCE IN 2 WEEKS UNDER MEDICAL SUPERVISION OVER 4-6 HOURS

REVIEW IN HEPATOLOGY OPD WITH REPORTS OF CBC/LFT/KFT/INR AFTER 4 WEEKS.

PLAN: CONTINUE STRIDE REGIMEN - 4-5 SESSION AFTER ASSESSING RESPONSE - - INJ DURVALUMAB - 1500 MG - 4 WEEKLY.

CONTINUE NEPHROLOGY FOLLOW UP

### Summary 19

**TRANSFER SUMMARY**

Inflammatory Bowel Disease – Crohn’s Disease (Biopsy Proven) **(ICD10 - K51.90)**

- s/p Pulse steroid f/b tapering over 1 month
- s/p Adalimumab – 18/02/2025
- on Methotrexate and Pentasa
- Perianal Abscess – s/p I & D on 15/02/2025 (outside)
- Hollow Viscous Perforation – sealed off – Conservatively Managed

Colonoscopy – 05/03/2025 – Ileal Ulcers and erosions; Large external and internal hemorrhoids

Biopsy – 05/03/2025 – Ileal Biopsy – Active ileitis; Colonic Biopsy – Active colitis with ulceration

MRI Pelvis: s/o Small Perianal Abscess

**COMORBIDITIES**: BPH

**CURRENT ISSUES:**

- Hollow Viscous Perforation – ?sealed off – Conservatively Managed
- Diarrhea (10 episodes per day)
- Dyselectrolemia

**PRESENTING COMPLAINTS:**

- Abdominal Pain x 4 days
- Fever x 1 day

**INDICATION FOR ADMISSION:**

Evaluation and the management of the symptoms

**HISTORY:**

Mr Patient S is a 63 year old gentleman with no known comorbidities. His index issue was in September 2024 2023 when he developed loose stools and fever. He presented to an outside centre for the same. He also had oral ulceration – painful palatal ulcers associated with dysphagia. ANA was positive and colonoscopy done outside confirmed diagnosis of Crohn’s disease. In February 2025, he received a tapering course of prednisolone for one month. However, active disease continued. He received Adalimumab in February 2025. He also received Pentasa and Methotrexate. He had a perianal abscess in February 2025, for which I & D was done. He was recently admitted in ILBS for further evaluation, from 03/03/2025 to 12/03/2025. He underwent Colonoscopy which revealed Ileal Ulcers and erosions; Large external and internal hemorrhoids. Biopsy reported Ileal Biopsy – Active ileitis and Colonic Biopsy – Active colitis with ulceration. Sigmoidoscopy was done on 10/03/2025 in view of bleeding P/R and revealed large external haemorrhoids with ulcer. His diarrhea frequency gradually improved, and bleeding P/R subsided. He was switched to Wysolone 20 mg, and Pentasa and Methotrexate continued. He recovered symptomatically following the management and is discharged in hemodynamically stable condition. However, he has currently presented with a pain abdomen and fever since 4 days. The pain was insidious in onset, localised to upper abdomen, dull aching, non-colicky, with no radiation or migration. He also had an episode of fever 1 day before presentation, which continued through the first day of admission. There is no h/o jaundice, vomiting, cough, abdominal pain, hematemesis, burning micturition, altered sensorium or decreased urine output. There is no h/o any intoxications, indigenous medications or IV drug abuse prior to onset of the disease.

**EXAMINATION**

Pt. was conscious, oriented to time place and person.

BP: - 126/70 mm Hg, Pulse: - 86/min, RR: - 14/min and afebrile.

Pallor -, Icterus - , Cyanosis - , Clubbing - , Pedal edema- , LNP-, JVP normal

On systemic examination

Respiratory system:--B/L vesicular breathing, B/L airway equal air entry, no wheeze,

Cardiovascular system: - S1 S2 normal, no murmurs

CNS: - conscious and oriented with no sensorimotor deficit,

Per Abdomen examination

ON INSPECTION: - abdomen not distended, umbilicus central and inverted, no visible venous prominences, no visible pulsations.

ON PALPATION: - soft, mild tenderness epigastrium, no organomegaly. No guarding, no rigidity, no rebound tenderness.

ON PERCUSSION: - Tympanic note with no fluid present

ON AUSCULTATION: - Normal bowel sounds present

**SYSTEMATIC REVIEW:**

Patient was admitted with above mentioned complaints. His examination findings were as mentioned above. His initial lab data and latest lab data is included at last of summary. Blood and Urine Cultures revealed no growth, and Urine Routine was unremarkable. S. PCT – 0.20. CXR revealed air under the diaphragm and urgent surgery referral was done. NPO was initiated, IV fluids and IV antibiotics were started, and an urgent surgery referral was undertaken. NCCT abdomen with oral gastrograffin was done – showed pneumoperitoneum with no obvious contrast leak - ?sealed off perforation, final report awaited. A conservative approach was opted for, and daily abdominal x-rays and screening was undertaken. Family and the patient were explained regarding the nature of disease and course undertaken. He symptomatically improved, with no further fever episodes and pain subsided. He was started gradually on oral liquids and subsequently, a soft diet as tolerated. His loose stools continued, and Pentasa was re-initiated. All vitals, temperature, RBS and other necessary parameters were checked regularly and managed appropriately. He is now discharged in hemodynamically stable condition with the advice to transfer to another hospital – Gastroenterology Department for further management.

**PLAN / ADVICE AT DISCHARGE (Including duration of medication if any):**

1800 KCAL/DAY + 80 GRAMS PROTEIN/DAY, LOW SALT (<2GRAMS/DAY)

INJ DORIPENEM 500 MG IV TDS (STARTED ON 24/03/2025)

ING GENTAMICIN 80 MG IV BD (STARTED ON 24/03/2025)

TAB NITAZOXANIDE 500 MG PO BD (STARTED ON 23/03/2025)

TAB RIFAGUT 550 MG PO BD

TAB PENTASA 1 GM PO QID

TAB TOFACETINIB 5 MG PO OD

TAB REDOTIL 100 MG PO BD

CAP VSL #3 PO BD 1-0-1

TAB PANTOCID 40 MG PO OD BBF

SYP SUCRAL 10 ML PO QID

SYP MUCAINE GEL 10 ML PO SOS

TAB FOLVITE 5 MG PO OD

TAB ZINC 50 MG PO TDS

TAB SILOSIN 8 MG PO OD HS

TAB SITCOM FORTE 1 TAB PO OD

SITZ BATH TO CONTINUE

ANOVATE GEL L/A TDS

ORS SOS IF LOOSE STOOLS

IN CASE OF DECREASE IN URINE OUTPUT, ALTERED SENSORIUM, BLEEDING, FEVER, NEW ONSET COUGH AND REVIEW IN ILBS EMERGENCY ON SOS BASIS.

### Summary 20

**1.)** Acute Necrotizing Pancreatitis (K85.9)

- Etiology: Under Evaluation
- With Sepsis(Bilateral Pneumonia – PCT (4🡪0.4) s/p Intubation, Extubated on 28/09/2024
- With Portal Vein Thrombosis (Cavernoma formation)
- With MPD leak s/p ERCP – PD stenting (12/09/2024)

**2.)** PHTN – (Non-bleeder, Small High risk Esophageal varices with Mild PHG, Pancreatic stent in situ - 08/10/2024)

?Secondary to PVT

Ascites (High SAAG, low protein)?PHTN related vs pancreatic – s/p LAR octreotide, on diuretics (Controlled)

CAP – 237 LSM – 18.2 SSM – 55.2

**3.)** Urethral Injury 🡪 SPC insertion (outside) 🡪 Foley’s in-situ at present

**PRESENTING COMPLAINTS:**

Pain abdomen x 2 days

Vomiting x 2 days

**INDICATION FOR ADMISSION:**

Evaluation and the management of the symptoms

**HISTORY:**

Mr. Patient T is a 38 year old gentleman, with known comorbidities of Type 3 C Diabetes Mellitus x 4 months. He had his index presentation at an outside centre in 2021 with abdominal pain that was insidious in onset, dull aching, non-colicky, with radiation to the back. He was diagnosed as Acute Pancreatitis. The pain was severe in intensity and required hospital admission for the same. He was managed conservatively and discharged within 15 days. Since then, he has had multiple recurrent attacks of similar pain, that resolve with painkillers. In July 2024, he had an attack of AP which required hospitalization, and was discharged in 20 days after conservative management. Episdoes of pain recurred and he had an MRCP done outside, and ERCP + PD stenting was done on 12/09/2024.After that, he developed a progressive abdominal distension and 1.5 Litres of fluid was tapped. He was admitted at the outside centre. He developed jaundice over the course of this admission. SPC was inserted for Urinary Tract obstruction on 19/09/2024. The distension was associated with a high grade fever and chills. The fever progressed with worsening sepsis and respiratory distress. He was intubated in view of worsening respiratory distress and altered behaviour secondary to septic shock. He was referred to ILBS for further management. He was admitted in the LCICU for further evaluation and management. There is no h/o any intoxications, indigenous medications, major surgeries, or IV drug abuse prior to onset of the disease. There is no h/o HTN/CAD/TB/COPD/Thyroid disorders.

**EXAMINATION:**

Pt. sedated, paralyzed.

BP: - 134/74 mm Hg, Pulse: - 70/min, RR: - 16/min and afebrile.

Pallor+, Icterus-, Cyanosis-, Clubbing-, Pedal edema (pitting type) - , LNP-, JVP normal

On systemic examination

Respiratory system: B/L vesicular breathing, B/L airway equal air entry, no crepts

Cardiovascular system: - S1 S2 normal, no murmurs

CNS: - conscious and oriented with no sensorimotor deficit,

Per Abdomen examination

ON INSPECTION: - Non-distended abdomen, umbilicus central and inverted, skin over the abdomen is stretched with no visible venous prominences, no visible pulsations.

ON PALPATION: - soft, tenderness over epigastrium, liver non- palpable. No guarding, no rigidity, no rebound tenderness.

ON PERCUSSION: - Tympanic note, No free fluid present.

ON AUSCULTATION: - normal bowel sounds present,

**SYSTEMATIC REVIEW:**

Patient was admitted with above mentioned complaints. His examination findings were as mentioned above. Initial lab data and latest lab data is included at last of summary. S. Amylase – 13. PEth positive. Patient denies intake of alcohol. Blood Cultures revealed no growth, and Urine Routine revealed 10-15 leukocytes. Urine C/S revealed fungal growth. S. PCT – 2.39. Ascitic fluid tap was done which revealed a straw coloured fluid with WBC - 332, N – 38% and L – 61%, Protein: 1.59, Glu: 174, Albumin: 0.3, SAAG: 1.94, ADA: 4.4 and Gene Xpert: negative, Gram Stain negative. Cultures were negative. Amylase level of ascitic fluid – 4. CECT Abdomen was done on 25/09/2024 reported

- the entire main portal vein and its right and left branches show hypodense thrombus extending to SPC, SMV its tributaries.
- Multiple collateral channels along peripancreatic, periportal, mesenteric regions seen. Associated, multiple tiny collaterals are also noted along paracholedochal, pericholecystic, juxtahilar and peribiliary regions.
- Stent is seen in MPD.
- Mild prominence of bilobar IHBR noted.
- Associated thin para-esophageal, perigastric, peripancreatic, and retroperitoneal collaterals are also seen.
- Multiple subcentimeter as well as prominent homogeneously hypoenhancing lymph nodes are seen in the upper abdomen mesentery and retroperitoneum.
- Pancreas displays normal parenchymal enhancement. No definite calcification is seen. MPD is not dilated.

MRCP was done on 07/10/2024 – report awaited. He was started on LMWH, high grade antibiotics and inotropes. Urology opinion was sought, and Foley’s inserted on 27/09/2024, and SPC removed. His clinical and laboratory parameters gradually improved, inotropic support was tapered and he was extubated on 28/09/2024. He was mobilized and daily physiotherapy done. Sensorium was still affecgted and psychiatry opinion sought for ? ICU psychosis, and advice followed. His condition gradually improved, and he was shifted to the ward on 05/10/2024. Antibiotics were downgraded. Urology consult taken and advised to leave Foley’s catheter in-situ. Other supportive medication and treatments continued. Further urine cultures were negative. He underwent a UGIE on 08/10/2024 revealed Small High risk Esophageal varices with Mild PHG, Pancreatic stent in situ. The nature of the disease and long term prognosis was explained in great detail to the patient and the patient relatives. He recovered symptomatically following the management and is discharged with following advice, and to follow up in OPD.

**ADVICE**

2100 Kcal/day + 60 grams protein/day, low fat diet

TAB TAXIM-O 200MG BD	1-0-1 X 5 DAYS

TAB FLUCOS 200 MG PO OD FOR 5 DAYS

TAB PANLIPASE 25K TDS WITH MEALS

TAB PANTOCID 40MG PO OD BEFORE BREAKFAST

TAB URSOCOL 450 MG PO BD

TAB CARDIVAS 3.125 MG PO BD (DO NOT GIVE IF HR < 55 OR BP < 90/60 MMHG)

CAP HENZOVIT 1 TAB PO OD

CAP ALCOMAX 300 MG PO BD

PEPTAMINE POWDER 2 SCOOPS PO BD

TAB LASILACTONE (20/50) ½ TABLET PO BD (MONITOR KFT WEEKLY)

**PLAN: - INJ OCTREOTIDE LAR 20 MG IM AFTER 4 WEEKS, ERCP +/- MPD STENTING AFTER 4 WEEKS**

### Summary 21

PHTN – Prior Bleeder, Eradicated Esophageal Varices, Mild PHG, Antral GAVE – 27/09/2024

Cirrhosis - NASH related **(ICD10 - K75.8)**

Decompensated – AVB – 2016, Ascites

CTP- 8, Child B, MELD Na-27

**Co morbidities:**

·         T2DM x 20 y, with diabetic retinopathy

·         HTN x 20 y

·         CKD - Stage VD (Diabetic nephropathy) on MHD (thrice a week)

·         Anemia - GAVE + Anemia of Chronic Disease

**CURRENT ISSUES-**

- Refilling ascites - No SBP, LVP done
- Anemia of chronic disease - PRBC transfused

**PRESENTING COMPLAINTS:**

- Loss of appetite and generalized pruritus from last 3-4 days
- Progressive abdominal distension from last 1 week

**INDICATION FOR ADMISSION:**

Evaluation & management of presenting complaints

**HISTORY:**

Mr Patient U is a 57 years old male f/u case of Cirrhosis-NASH with co morbidities T2DM/ Diabetic nephropathy (CKD Stage 5, Baseline 4.41)/ HTN on management and regular MHD sessions. His index presentation was in 2016 in the form of hemetemesis without any postural symptoms. He also had complaint of ascites which was insidious in onset, gradually progressive and not associated with abdominal pain. On evaluation, he was diagnosed as a case of CLD (NASH related) with PHTN. He has had prior admissions in view of ascites, pain abdomen; loose stools for which he was managed conservatively and discharged with nephrology follow up advised on OPD basis. Currently, he has presented with complaints of progressive abdominal distension and loss of appetite and generalized weakness. He has been admitted for further evaluation and management. There is no h/o vomiting, altered bowel habits, hematemesis, malena, burning micturition or decreased urine output. There is no h/o any intoxications, indigenous medications, blood transfusions or IV drug abuse prior to onset of the disease. There are no h/o CAD/TB/COPD/Thyroid disorders.

**EXAMINATION:**

Pt. was conscious, oriented, a febrile

BP-140/80 mmHg, Pulse 84/min, RR -18/min

Pallor+, Icterus-, Cyanosis-, Clubbing-, Pedal edema+, LNP-, JVP normal

CVS -S1 S2 normal, no murmurs

Chest- B/L normal air entry &amp; vesicular breathing, no adventitious sounds

CNS -Patient conscious, well oriented, no sensorimotor deficit.

P/A ON INSPECTION grossly distended, umbilicus centrally located and inverted. No visible

superficial veins +, no visible pulsations.

ON PALPATION -soft, non tender, liver - non palpable spleen - non palpable. No guarding,

no rigidity, no rebound tenderness.

ON PERCUSSION - dull note, free fluid present.

ON AUSCULTATION- normal bowel sounds present, no bruits.

**SYSTEMATIC REVIEW:**

Patient was admitted with the above mentioned plan. His examination findings were as mentioned above. Blood cultures revealed no growth. Ascitic fluid tap was done which revealed a straw coloured fluid with WBC - 137, N – 39% and L – 61%, Protein: 2.08, Glu: 74.5, Albumin: 0.72, SAAG: 1.62, ADA: 23.8 and Gene Xpert: negative, Gram Stain negative. Nephrology review was done and advice was followed, and he underwent two MHD sessions on 04/11/2024 and 06/11/2024. PET-CT was done on 06/11/2024 –

- Features of CLD with portal hypertension (splenomegaly, abdominal collaterals, dilated s/p axis and moderate ascites).
- Metabolically active multiple areas of patchy pleural thickening involving the right lung with loculated right pleural effusion -likely infective (? TB).
- Metabolically active enlarged lymph nodes in the right paratracheal, prevascular and subcarinal regions-likely part of same disease.
- Bilateral small kidneys with reduced cortical thickness –CKD.
- No evidence of abnormal metabolically active lesions noted elsewhere.

He was managed with IV antibiotics, blood products, nutritional therapy & other supportive measures. ATT was initiated on 07/11/2024. Now he is being discharged in haemodynamically stable state. Nephrology consult taken and there advised followed. The advanced nature of the disease and its prognosis was explained in great detail to the patient and his attendants.

**PLAN / ADVICE AT DISCHARGE (Including duration of medication if any):**

2100 KCAL/DAY + 50 GRAMS PROTEIN/DAY, LOW SALT (LESS THAN 2GRAMS/DAY), DIABETIC RENAL DIET

T. INH 100 MG PO OD

T. BENADON 40 MG PO OD

TAB LEVOFLOX 500 MG PO OD

TAB COMBUTOL 800 MG PO OD

TAB THALIDOMIDE 50 MG PO OD

T. CARCA 12.5MG PO BD 1-0-1

T. CILACAR 10MG PO BD 1-0-1

T DYTOR 40MG PO BD 1-0-1 – ON NON-HD DAYS

T. NODOSIS 500MG PO TDS 1-1-1

T. NEFITA PO OD 1-0-0 (ALTERNATE DAY)

T. SHELCAL 500MG PO BD 1-0-1

T. TRAJENTA 5MG PO OD 1-0-0

NEPHRO HP POWDER 2SCOOPS PO TDS 1-1-1

LAXOPEG SACHET PO SOS

INJ. DARBA 60IU SC ONCE A WEEK

ADV: BLOOD SUGAR MONITORING (WARNING SIGNS OF HYPOGLYCEMIA EXPLAINED)

REVIEW IN HEPATOLOGY OPD WITH CBC/LFT/KFT/INR REPORTS AFTER 3 WEEKS.

PLAN- NCV FOLLOWED BY THALIDOMIDE INITIATION

REVIEW IN NEPHROLOGY OPD AND FOLLOW THEIR ADVICE; ALBUMIN SUPPLEMENTATION TO BE DONE DURING MHD SESSIONS.

IN CASE OF DECREASE IN URINE OUTPUT, ALTERED SENSORIUM, BLEEDING, FEVER, NEW ONSET COUGH AND REVIEW IN ILBS EMERGENCY ON SOS BASIS.

### Summary 22

PHTN - (Non Bleeder, Small LR Esophageal Varices, Mild PHG - 12/08/2023

Cirrhosis Ethanol related (LI - Aug 2023) **[ICD K 70.9]**

Decompensated with Ascites / HE

Prior ACLF with Acute: SAH s/p steroids (Responder) and Chronic: ALD

CTP – 8	Child – B	MELD - 15

**CURRENT ISSUES:**

- Hepatic Encephalopathy Grade III 🡪 Resolved

**PRESENTING COMPLAINTS:**

Altered Sensorium x 2 days

**INDICATION FOR ADMISSION:**

For further evaluation and management of symptoms

**HISTORY:**

Mr. Patient V is a 49 years old male with no known comorbidities with chronic alcohol intake(LI-Aug 23) He was apparently alright until July 2023 when he started developing jaundice with yellowish discolouration of urine and sclera, painless, gradually progressive, non pruritis, not associated with clay coloured stool, followed by abdominal distension which was insidious in onset, gradually progressive, associated with pedal edema, not associated with burning micturition  and also associated with decreased urine output. He was then admitted in ILBS and diagnosed as ACLF. Liver biopsy (TJLB) was done showed -Alcoholic Steatohepatitis with Cirrhosis and started on steroid therapy and was steroid responder and was discharged. He was again admitted in October 2023 and managed as case of pneumonia and was subsequently discharged. Now he has presented to ILBS with complaints of altered sensorium and got admitted for further evaluation and management. There is no h/o hemoptysis, hematemesis, altered bowel habit. There is no h/o any recidivism, indigenous medications, blood transfusions or IV drug abuse prior to onset of the disease. There is no h/o T2DM/HTN/TB/COPD/Thyroid disorders.

**EXAMINATION:**

Pt. was conscious, oriented to TPP, afebrile.

BP -107/76 mm Hg , Pulse -78/min    RR -20 /min

Pallor+, Icterus+ , Cyanosis-, Clubbing+, Pedal edema-,LNP-, JVP normal

CVS -S1 S2 normal, no murmurs

Chest- B/L normal air entry & vesicular breathing, crepts +

CNS - conscious, Oriented, no focal neurological deficit.

P/A ON INSPECTION- mild distended, umbilicus central and inverted, no visible venous prominences, no visible pulsations.

ON PALPATION- soft, non tender, liver -palpable 2 cm BCM and Spleen - just palpable below left costal margin. No guarding, no rigidity, no rebound tenderness.

ON PERCUSSION- dull note, with free fluid present

ON AUSCULTATION- normal bowel sounds present, no bruits.

**SYSTEMATIC REVIEW:**

Patient was admitted with the above mentioned complaints. His examination findings were as mentioned above. Blood and Urine Cultures revealed no growth, and Urine Routine was unremarkable. S. PCT – 0.22. Plasma Ammonia – 264.1. CT Head – Normal Study. He was shifted to the HDU. Anti-coma measures initiated. He was started on IV antibiotics, and supportive medications until she symptomatically improved. Sensorium improved and she was symptomatically better. He was shifted to the ward on 30/11/2024. Blood products transfused as required. The nature of the disease and long term prognosis was explained in great detail to the patient and the patient relatives and were also explained regarding the need of liver transplant in view of Decompensated Liver Disease. Strict alcohol abstinence advised. She is now planned for nutritional optimization and improvement in performance status. Proper diet and mobilisation was followed throughout the course of admission. All vitals, temperature, RBS and other necessary parameters were checked regularly and managed appropriately. She is currently being discharged in hemodynamically stable condition, with the advice to follow in OPD.

**PLAN / ADVICE AT DISCHARGE (Including duration of medication if any):**

30 MINS DAILY PHYSICAL ACTIVITY, STRICT ALCOHOL AND TOBACCO ABSTINENCE

TAB FAROPENAM 200 MG PO BD FOR 5 DAYS

TAB FLUCOS 200 MG PO OD FOR 5 DAYS

TAB CIPLAR LA 20 MG PO HS (DO NOT GIVE IF BP < 90/60 AND HR < 55)*

SYP ZINCONIA 10 ML PO BD

SYP LACTIFIBER 20 ML PO BD (ENSURE 2-3 BOWEL MOTIONS/DAY)

LAXOPEG SACHET BD. 1-0-1 (ENSURE 2-3 BOWEL MOTIONS/DAY)

MOVICOL SACHET 1 SACHET PO SOS

LORHEP SACHET PO BD

CAP HENZOVIT 1 TAB PO OD

TAB ME-12 1 TAB PO OD

REVIEW IN HEPATOLOGY OPD WITH REPORTS OF CBC/LFT/KFT/INR AFTER 2 WEEKS

### Summary 23

PHTN - Bleeder, Small High Risk Esophageal Varices, EVL done, Mild PHG – 02/03/2025

CLD- NAFLD	  **(ICD10 - K75.8)**

Compensated

Disseminated Kochs vs Sarcoidosis (Calcium raised, ACE-115 outside)

- S/P- LN FNAC outside- Granulomatous Disease- Outside
- S/P ATT outside x 3weeks 🡪 No relief
- S/P - Diagnostic Bronchoscopy BAL negative for malignancy / TB
- Endobronchial Biopsy from right secondary carina Non-necrotizing granulomatous inflammation. No evidence of malignancy.
- S/P - Modified ATT started on 01/07/2024

CTP – 10 C	MELD Na – 15

**CURRENT ISSUES:-**

- UGI Bleed with Hematemesis
  - UGIE - Small High Risk Esophageal Varices, EVL done, Mild PHG – 02/03/2025
- AKI KDIGO I - Diuretic Related (S. Creat 1.23 🡪 1.01)
- Generalized Lymphadenopathy ?Disseminated Kochs + Sarcoidosis
  - On Modified ATT started on 01/07/2024
  - Prednisolone started on 04/03/2025 – Plan to taper

**CO MORBIDITIES-**

-          T2DM

-          Hypothyroidism

**PRESENTING ISSUES-**

-          Generalized Weakness x 7 days

-          Cough and shortness of breath x 7 days

**INDICATION FOR ADMISSION:**

Evaluation and management of symptoms

**HISTORY-**

Mr W is a 61 year old gentleman with k/c/o co morbidities of T2DM and Hypothyroidism. He had his index presentation in form of cough 1 year ago which was associated with weakness and significant weight loss. He was evaluated at an outside centre and was diagnosed as TB for which he was given ATT for a period of three weeks . However, he did not showe response and stopped ATT. He was then admitted in ILBS and reviewed. Sarcoidosis and TB were kept as the possibilities and he was evaluated further. Endobronchial Biopsy from right secondary carina Non-necrotizing granulomatous inflammation. No evidence of malignancy. He was initiated on modified ATT on 01/07/2024 and was discharged in satisfactory condition. He was recently admitted in ILBS with a shortness of breath. HRCT lung was done on 29/01/2025 reported - bilateral hilar and mediastinal lymphadenopathy with nodular-and-irregular thickening of the peribronchovascular interstitium with bilateral perihilar mild architectural distortion and traction bronchiolectasis as described. Findings likely represent pulmonary sarcoidosis. Mild interval reduction of lymph nodes likely post treatment. ATT was continued and he was discharged in satisfactory condition. He was admitted at an outside centre 15 days ago for evaluation of generalized lymphadenopathy an d PET-CT was done s/o increase in size of lymph nodes. In view of high ACE levels and serum calcium levels, he was started on trial of methylprednisolone on 24/02/2025. He has currently presented with complaints of blood in vomitus – 1 episode. There is no h/o jaundice, abdominal pain, altered bowel habits, burning micturition, altered sensorium or decreased urine output. There is no h/o any intoxications, indigenous medications, major surgeries or IV drug abuse prior to onset of the disease. There is no h/o CVA/ CAD/HTN/COPD.

**EXAMINATION:**

Pt. was conscious, oriented, afebrile

BP 124/76 mm Hg	Pulse 74/min	RR 20/min

Pallor+, Icterus-, Cyanosis-, Clubbing-, Pedal edema-, LNP-, JVP normal

CVS S1 S2 normal, no murmurs

Chest B/L decrease right sided air entry & vesicular breathing, wheeze present

CNS Patient conscious, oriented, no sensorimotor deficit.

P/A ON INSPECTION non-distended, umbilicus central and inverted, no visible venous prominences, no visible pulsations.

ON PALPATION- soft, non-tender, liver not palpable, Spleen not palpable, No guarding, no rigidity, no rebound tenderness.

ON PERCUSSION -tympanic note, no free fluid

ON AUSCULTATION normal bowel sounds present, no bruits.

**SYSTEMATIC REVIEW:**

Mr. Patient W was admitted with above mention complaints. His examination findings were as mentioned above. His initial and latest lab data is to be found at the end of summary in form of lab charts. Blood and Urine Cultures revealed no growth, and Urine Routine revealed 6-8 leukocytes. He was shifted to the GI Bleed Unit in view of hematemesis and Bleeder Protocol initiated. He was started on Terlipressin and fluid resuscitation. UGIE revealed Small High Risk Esophageal Varices, EVL done, Mild PHG on 02/03/2025.

Bloods revealed an AKI I. He was shifted to the GI Bleed ICU and started on IV antibiotics, vasopressors, nutritional support and other supportive medication and treatment until he symptomatically improved. Pulmonology opinion was sought throughout admission and advice followed. He gradually improved symptomatically, and oxygen support was tapered and patient improved. He was shifted to the ward. A review of outside PET-CT was done on 04/03/2025 –

- Features of CLD with portal hypertension (splenomegaly, mildly dilated s/p excision abdominal collaterals).
- Metabolically active nodular and irregular thickening involving the peribronchovascular interstitium with multiple perilymphatic parenchymal nodules involving both lungs along with enlarged lymph nodes on either side of diaphragm and multiple ill-defined discrete and confluent liver lesions -residual disease involvement.
- Bilateral pleural effusion (right>left).
- No evidence of abnormal metabolically active lesions noted elsewhere.

He was started on Prednisolone 40 mg on 04/03/2025, with plan to taper down on OPD basis. The nature of the disease and long term prognosis was explained in great detail to the patient and the patient relatives. He is now planned for nutritional optimization and improvement in performance status. Proper diet and mobilisation was followed throughout the course of admission. All vitals, temperature, RBS and other necessary parameters were checked regularly and managed appropriately. He is currently being discharged in hemodynamically stable condition, with the advice to follow in OPD.

**PLAN / ADVICE AT DISCHARGE (INCLUDING DURATION OF MEDICATION IF ANY):**

1600KCAL, 80 GM PROTEIN, NORMAL DIET LOW CALCIUM DIET

TAB TAXIM-O 200MG BD	1-0-1 X 5 DAYS

TAB FLUCOS 200 MG PO OD FOR 5 DAYS

TAB PANTOCID 40 MG PO OD BBF

SYP SUCRAL 10 ML PO QID X 14 DAYS

TAB CARDIVAS 3.125 MG PO BD (DO NOT GIVE IF HR < 55 OR BP < 90/60 MMHG)

**TAB PREDNISOLONE 40 MG PO OD x 1 WEEK, FOLLOWED BY 30 MG PO OD X 1 WEEK, FOLLOWED BY 20 MG OD FOR 1 WEEK, FOLLOWED BY 10 MG OD FOR 1 WEEK, FOLLOWED BY 5 MG OD TO CONTINUE**

**TAB LEVOFLOX 500 MG PO OD 1-0-0**

**TAB ETHAMBUTOL 800 MG PO OD 1-0-0**

TAB FOLVITE 5 MG PO OD

TAB SHELCAL 1 TAB PO OD

TAB UDCA 450 MG PO BD

TAB THYRONORM 50 MCG PO BBF

TAB SILDOFAST 8MG PO OD 0-0-1

TAB FEBUXOSTAT 40MG PO OD 0-0-1

HEPSURE SACHET PO BD

CAP HENZOVIT 1 TAB PO OD

NUTRIX ULTRA 2SCOOP PO TDS 1-1-1

SYP LACTIHEP 30ML PO HS  0-0-1 (ENSURE 2-3 BOWEL MOTIONS/DAY)

LAXOPEG SACHET SOS (ENSURE 2-3 BOWEL MOTIONS/DAY)

REVIEW AFTER 2 WEEKS IN HEPATOLOGY AND PULMONOLOGY OPD WITH CBC/KFT/LFT/INR/ CXR-PA REPORTS

FOLLOW UP IN PULMONARY MEDICINE OPD

PLAN: TAPER STEROIDS / UGIE + EVL AFTER 3 WEEKS / ADD DIURETICS ON OPD BASIS

F/U IN VIRTUAL OPD AFTER 1 WEEK WITH CBC/LFT/KFT REPORTS

IN CASE OF ALTERED SENSORIUM, DECREASE IN URINE OUTPUT, BLEEDING, FEVER, NEW ONSET COUGH AND REVIEW IN ILBS EMERGENCY ON SOS BASIS.

### Summary 24

PHTN – (Non-Bleeder / No varices – 13/05/2025)

CLD - PSC **(ICD10 - K75.8 + K83.01)**

- Outside Liver Biopsy –Chronic Biliary Pathology with changes of sclerosing cholangitis; Ludwig stage 2/4; Negative for malignancy

CAP – 124	LSM – 5.6	SSM – 18.8

Decompensated - Ascites

MELD-9	CTP- 8B

Inflammatory Bowel Disease - Ulcerative Colitis

- Outside Colon Biopsy – Chronic colitis with moderate activity – Ulcerative colitis
- s/p Colonoscopy – 13/05/2025--- Inflammatory Bowel Disease ?Ulcerative Colitis; **Biopsy Awaited**

**COMORBIDITIES -** None

**PRESENTING COMPLAINTS**:

- Loss of Appetite x 2 months
- Weight Loss x 2 months

**INDICATION FOR ADMISSION:**

For further evaluation and management of the presenting symptoms

**HISTORY:**

Mr. Patient X is a 21-year old gentleman with no history of substance abuse or addiction. He had his index presentation 2 months ago in the form of altered bowel movements. He was conservatively managed for the same. He then developed a pain abdomen which worsened on defecation. He also noticed stool mixed with blood. He was evaluated for the same and colonoscopy revealed Pancolitis - Ulcerative Colitis. Biopsy was done revealed - Chronic colitis with moderate activity – Ulcerative colitis. A liver biopsy was done which revealed Chronic Biliary Pathology with changes of sclerosing cholangitis; Ludwig stage 2/4; Negative for malignancy. He was initiated on Wysolone and Azathioprine for the same. Currently, he has presented with complaints of loss of weight and generalized weakness.Patient has been admitted for further evaluation and management. There is no h/o jaundice, cough, abdominal pain, hematemesis, and malena, burning micturition, altered sensorium or decreased urine output. There is no h/o any intoxications, indigenous medications, major surgeries, blood transfusions or IV drug abuse prior to onset of the disease. There is no h/o HTN/CAD/TB/COPD/Thyroid disorders.

**EXAMINATION:**

Pt. was conscious, oriented and afebrile.

BP- 107/60 mm Hg	Pulse- 79/min	RR- 18/min

Pallor-, Icterus-, Cyanosis-, Clubbing-, Pedal edema-, LNP-, JVP normal

CVS - S1 S2 normal, no murmurs

Chest - B/L air entery +, no additional sounds

CNS- Patient conscious, oriented.

P/A ON INSPECTION- non-distended umbilicus central, no visible venous prominences and no visible pulsations.

ON PALPATION - soft; non tender, liver not palpable and spleen not palpable. No guarding, rebound tenderness absent.

ON PERCUSSION- tympanic note, free fluid present.

ON AUSCULTATION- normal bowel sounds present.

**SYSTEMATIC REVIEW:**

Patient was admitted with above mentioned complaints. His examination findings were as mentioned above. His initial lab data and latest labs are at the end of summary in chart form. He was managed conservatively with IV antibiotics, IV fluids, nutritional therapy & other supportive measures.Stool GI Panel – ND. Viral markers – Negative. CA 19-9 – 40.2.HLA B27 – Negative. UGIE was done - No varices – 13/05/2025. Colonoscopy was done on 13/05/2025 - Inflammatory Bowel Disease ?Ulcerative Colitis. He was restarted on Azathioprine and prednisolone, along with mesalamine for IBD. Proper diet and mobilization was followed throughout the course of admission. He symptomatically improved, and further hospital course was uneventful. All vitals, temperature, and other necessary parameters were checked regularly and managed appropriately. He recovered clinically and discharged in hemodynamically stable state.

**PLAN / ADVICE AT DISCHARGE (Including duration of medication if any):**

1800 KCAL/DAY + 90 GRAMS PROTEIN/DAY

**PERSONAL PROTECTION AND HYGIENE AS ADVISED.**

**AVOID SOCIAL GATHERINGS, WEAR MASK REGULARLY.**

TAB TAXIM-O 200MG BD	1-0-1 X 5 DAYS

TAB WYSOLONE 30 MG PO OD

TAB AZORAN 50 MG PO OD

T. MESACOL 1.2 GM PO BD

MESACOL SUPPOSITORY P/R HS OD

TAB BILYPSA 4 MG PO OD

TAB PANTOCID 40 MG PO OD BBF

TAB SHELCAL PO OD

CAP HENZOVIT 1 TAB PO OD

TA BETADEK PO OD

TAB FOLVITE 5 MG PO OD

TAB URSOCOL SR 450 MG PO BD

FOLLOW UP IN HEPATOLOGY OPD AFTER 4 WEEKS WITH CBC/KFT/INR/LFT/ BIOPSY REPORTS

### Summary 25

Hyperacute Liver Failure - HAV related (KCH criteria: 0/5)

Jaundice to HE 🡪3 days

- s/p 3 sessions of cytokine filter
- S/P CRRT

**COMORBIDITY :**

None

**CURRENT ISSUES –**

- Sepsis with Septic Shock 🡪 Resolved
  - Blood c/s Staphylococcus hemolyticus
  - HD Catheter Tip Culture – CONS
  - Urine C/S – Klebsiella pneumoniae
- Hepatic encephalopathy grade III s/p Mechanical ventilation🡪 Tracheostomy 🡪 Removed
- UGI Bleed : UGIE duodenopathy changes at D1 and D2 with oozing from D2
- Jaundice (T. Bil: 15.13 🡪 8)

`

**PRESENTING COMPLAINTS**

- Fever x 9 days
- Generalized weakness x 8 days
- Shortness of breath x 4 days
- Altered behaviour x 3 days

**INDICATION FOR ADMISSION:**

Evaluation & management

**HISTORY:**

Mrs Patient Y is a 32 year old lady had his index presentation on 1^st^ Feb in the form of fever for which he took some medication. Following which he developed generalized weakness. 3 days later, he developed breathlessness, altered behaviour in the form of decreased talking, yellowish discoloration of eye. He was admitted in outside hospital where he was intubated, and referred to ILBS hospital with above mentioned complaints. He was admitted for further evaluation and management. There is no h/o past major surgeries, or IV drug abuse prior to onset of the disease. There is no h/o T2DM/HTN/CAD/COPD/Thyroid disorder.

**EXAMINATION:**

At the time of admission

Pt. was in intubated state

BP: - 149/83 mm Hg, Pulse: 81/min, RR: - 24/min, Spo2 100% on RA.

Pallor -, Icterus +, Cyanosis-, Clubbing-, Pedal edema -, LNP-, JVP Normal

On Systemic Examination

Respiratory System: - B/L vesicular breathing, B/L airway equal air entry, no wheeze, no crepts

Cardiovascular system: - S1 S2 normal, no murmurs

CNS: - E1VtM1

Per Abdomen examination

ON INSPECTION: - Non distended abdomen, umbilicus central, no visible venous prominences, no visible pulsations.

ON PALPATION: - Soft, non tender, liver palpable 2 cm below coastal cartilage, Spleen non palpable. No guarding, no rigidity, no rebound tenderness.

ON PERCUSSION: - Tympanic note, No free fluid is present

ON AUSCULTATION: - Bowel sounds present

**SYSTEMATIC REVIEW:** Patient was admitted with above mentioned complaints. His examination findings were as mentioned above. Investigations are listed below in chart form. Patient was received in ER. Immediate CCM review was taken and advice followed. Patient was shifted to LC ICU. Patient was started on aggressive management with IV antibiotics, IV anti-fungals, IV fluids, IV albumin and other supportive measures. IV inotrope was started in view of shock.

NCCT WA:- Liver is enlarged (craniocaudal span 21-cm) in size show normal outline with diffuse reduced attenuation. No focal lesion / IHBR are seen. GB is partially distended show diffusely thickened edematous wall. CBD is not dilated. Few subcentimeteric and some prominent upper abdomen and periportal lymph nodes are seen. Pancreas has normal outlines and parenchymal attenuation. No definite calcification is seen. MPD is not dilated. Spleen span is approx. 17-cm. Bilateral kidneys show normal size appear globular. Right kidney shows upper pole subcentimeter calculus. Stomach, visualized small and large bowel loops are unremarkable. Minimal free fluid is seen in the abdomen pelvis.

NCCT HEAD:- Bilateral cerebral parenchyma shows no significant focal brain parenchymal lesion. Bilateral basal ganglia and thalami are unremarkable. Ventricular system appears normal in size and shape. Brain stem and bilateral cerebellar hemispheres are unremarkable. No overt CP angle mass lesion is seen. IHF is in mid line. Sella and parasellar region reveal no definite abnormality. No e/o epidural / sub-dural collection is present.

HRCT CHEST:- Bilateral mild pleural effusion with basal atelectasis. Few subcentimeter mediastinal lymph nodes noted. Trachea is in the mid line. Main and segmental bronchi show no abnormality. No e/o bronchiectasis is seen. The mediastinal structures including the non-opacified aorta and mediastinal vessels are unremarkable. No evidence of pericardial effusion is seen.

**Anti HAV- Reactive. HAV RNA detected.** Blood c/s- no growth, urine c/s- no growth. Mini BAL sample was sent, Biofire - no organism detected, AFB gene xpert - ND, AFB- NG, KOH - NG, GS - NG, C/S - normal commensal flora grown. IV antibiotics upgraded accordingly. Respiratory galactomanan - 0.1. IV antifungal upgraded accordingly. Patient was started on CRRT on 09/02/2025. HbA1c- 6.43, Dengue ns 1 Ag, Leptospira IgM- NR. Viral markers were sent HBsAg/Anti HCV/Anti HEV/Anti HB core/HIV - Non reactive. Urine R/M- alb/sugar- 2+/nil, RBC/WBC- 0-1/2-4. CMV IgG/EBV IgG/HSV 1 &2 IgG- Reactive, VZV IgG- NR. Transcranial Doppler was done which showed- RMCA- 0.87 and LMCA- 0.72. Nephrology review was taken and advice followed. Patient underwent a session of cytokine filter on 12/02/2025 i/v/o hypercytokinemia. HPB surgery review was taken and advice followed. Need for LT was explained to the attendants. EEG graph was suggestive of encephalopathy. However with conservative measures encephalopathy improved and serial ONSD and TCD showed improvement with improving INR and ammonia .Pulmonology review was taken and advice followed. Cardiology review was taken and advice followed. PLAIN ECHO:- Normal LV size and normal systolic function. LA normal size. No clot. RV &RA normal size. RV contractility normal. No LA/LV clot. No pericardial effusion. EEG graph was suggestive of encephalopathy with drug activity. Mini BAL c/s- no growth. blood c/s and urine c/s - no growth. Urine c/s and urine c/s showed no growth. Mini BAL c/s - normal commensal growth. Patient was being planned for weaning for which sedation break was given however the patient was biting tube persistently and thus it was becoming challenging to wean him off . Thus , he was tracheostomised on 17/2/25. Following which he was doing well , however he had persistent fever for which he underwent 2^nd^ session of cytokine filter on 17/02/25. Patient developed shock and was put on inotropic support. He underwent another session of cytokine filter on 21/02/25 i/v/o persistent hypercytokinemia. Therapeutic bronchoscopy was done on 21/02/25 i/v/o thick secretions. Patients shock improved and inotropic support was gradually tapered & discontinued. Repeat blood culture showed Enterococcus faecium growth following which all IV lines were changed and antibiotics were revised accordingly. Tracheostomy tube was changed on 27/02/25. Repeat blood culture showed Staphylococcus haemolyticus growth following which antibiotics were revised accordingly. Patient underwent underwent UGIE on 1/03/2025 i/v/o malena which was suggestive of duodenopathy changes at D1 and D2 with oozing from D2 .Routine CCM review is being done. Tracheostomy closure was done on date 03/03/2025. Adequate blood products and ROTEM based coagulation correction was done. He gradually improved and was shifted to the ward. The nature of the disease and course was explained in great detail to the patient and the patient relatives. Patient remained afebrile with no significant complains in the ward. Proper diet and mobilization was followed throughout the course of admission. All vitals, temperature, RBS and other necessary parameters were checked regularly and managed appropriately. He recovered symptomatically following the management and is discharged with the advice to follow up in OPD.

**PLAN / ADVICE AT DISCHARGE (Including duration of medication if any):**

2100 KCAL/DAY + 90 GRAMS PROTEIN/DAY NORMAL DIET

AVOID ANY ALTERNATIVE MEDICATIONS

INJ ELORES 1.5 GM IV BD x 3 DAYS

FOLLOWED BY TAB FAROPENAM 200 MG PO BD FOR 5 DAYS (START ON DAY 4)

INJ TEICOPLANIN 400 MG IV OD X 3 DAYS

TAB FLUCOS 200 MG PO OD x 3 DAYS

TAB PANTOCID-DSR 1 TAB PO OD BBF X 14 DAYS

SYP SUCRAL 10 ML PO QID X 14 DAYS

TAB HEPKART 400 MG PO OD

CAP HENZOVIT 1 TAB PO OD

REVIEW IN OPD WITHIN 4 WEEKS WITH CBC, LFT, KFT, INR REPORTS

CBC/LFT/KFT/INR AFTER 7 DAYS AND INFORM IN VOPD

BED REST FOR 3 WEEKS

LIGHT WORK ADVISED FOR 2 MONTHS

IN CASE OF PAIN ABDOMEN, CONTINUED FEVER OR ANY BLACK STOOLS INFORM SOS

IN CASE OF DECREASE IN URINE OUTPUT, ALTERED SENSORIUM, BLEEDING, FEVER, NEW ONSET COUGH AND REVIEW IN ILBS EMERGENCY ON SOS BASIS.

### Summary 26

Cholelithiasis with Choledocholithiasis

- S/P EUS + EPT+ ERCP guided Stenting (Outside: 13/09/2024)

          -  Post Stenting bleed🡪 S/P ERCP guided SEMS placement (Outside)

          -  Post SEMS cholangitis; S/P ERCP guided RHD stenting (03/10/2024); c/s on 03/10/2024 - E. coli + K. Pneumoniae) and iv antibiotics as per C/S

**Co morbidity** - GSD

**Current issue:** Acute Cholecystitis

ICD- K80.8

**PRESENTING COMPLAINTS:**

Fever x 3 days

**INDICATION FOR ADMISSION:**

Evaluation and the management of presenting symptoms

**HISTORY:**

Mrs. Patient Y is a 34 years old lady with prior co morbidity of GSD. In September she developed pain abdomen and persistent vomiting. She was evaluated outside and MRCP showed choledocholithiasis. She underwent ERCP + Stenting on 13/09/2024. Procedure went uneventful but after procedure patient developed malena episodes without any postural symptoms. She again underwent rescue ERCP and SEMS placement was done for Haemorrhage control. PRBC transfusion was also done, and she was discharged form the outside centre in hemodynamically stable condition. Following this she developed a fever with acute cholangitis and presented to ILBS for the same. She was admitted and ERCP guided RHD stenting done on 03/10/2024, and discharged. She now complaints of further episode of fever since the past two days. With this clinical picture she has been admitted for further evaluation and management. There is no h/o jaundice, cough, altered bowel habits, hematemesis, and malena, burning micturition, altered sensorium or decreased urine output. There is no h/o any intoxications, indigenous medications, major surgeries, blood transfusions or IV drug abuse prior to onset of the disease. There is no h/o DM/ CAD/TB/COPD/Thyroid or renal disorders.

**EXAMINATION:**
Pt. was conscious, oriented to time place and person.
BP: - 120/70 mm Hg, Pulse: - 86/min, RR: - 14/min and afebrile.
Pallor-, Icterus-, Cyanosis-, Clubbing-, Pedal edema (pitting type) -, LNP-, JVP normal
On systemic examination 
Respiratory system: B/L vesicular breathing, B/L airway equal air entry, no crepts
Cardiovascular system: - S1 S2 normal, no murmurs
CNS: - conscious and oriented with no sensorimotor deficit,
Per Abdomen examination
ON INSPECTION: - flat abdomen, umbilicus central and inverted, skin over the abdomen is stretched with no visible venous prominences, no visible pulsations with excessive scaling
ON PALPATION: - soft, non-tender, liver non-palpable. Spleen is also non-palpable. No guarding, no rigidity, no rebound tenderness.
ON PERCUSSION: -Tympanic note with no fluid present
ON AUSCULTATION - normal bowel sounds present,

**SYSTEMATIC REVIEW:**
Patient was admitted with above mentioned complaints. Her examination findings were as mentioned above. Her initial lab data has been mentioned below the summary. In view of suspected sepsis she was initiated on broad spectrum antibiotic cover. Round the clock vital monitoring and febrile episodes were monitored. USG screening revealed GB distended and shows multiple calculi and sludge withinits lumen of ~ size 10 mm with symmetrically thickened GB wall with s/o acute cholecystitis. CBD stent in situ. No evidence of choledocholethiasis. No IHBR dilatation seen. Bilobar pneumobilia seen. There were no febrile episodes during hospitalization. The nature of disease and its prognosis has been explained to patient and her attendant in full details. She has been managed conservatively and is now being discharged in stable state to be readmitted for ERCP + Stent Removal on 21/10/24.

**ADVICE ON DISCHARGE (Duration of medications if any):**

1800 Kcal/day + 70 grams protein/day, normal full fat diet as advised

TAB FAROPENAM 200 MG PO BD FOR 5 DAYS THEN STOP

TAB TAXIM-O 200MG BD	1-0-1 X 5 DAYS THEN STOP

TAB PANTOCID 40 MG PO BBF

TAB UDCA 450 MG PO BD

CAP HENZOVIT 1 TAB PO OD

TAB ULTRACET PO SOS IF PAIN

READMIT (DIRECT ADMISSION) ON 21/10/2024 FOR ERCP+STENT REMOVAL.

**PLAN: ERCP + STENT REMOVAL ON 22/10/2024; F/B INTERVAL CHOLECYSTECTOMY**

IN CASE OF DECREASE IN URINE OUTPUT, ALTERED SENSORIUM, BLEEDING, FEVER, NEW ONSET COUGH AND REVIEW IN ILBS EMERGENCY ON SOS BASIS.

### Summary 27

Acute on Chronic – Day 80 (K85.92)  -	?Etiology

- Local complication – Intrapancreatic collection, Lesser Sac collection (3 x 4 cm)
- Disconnected Pancreatic Duct Syndrome
- h/o Pancreatic ascites (s/p PCD – removed on 15/10/2024)

**Comorbidities:** Nil

**Current Issues**: Disconnected Pancreatic Duct Syndrome - s/p ERCP + Pancreatic Duct Stenting on 15/10/2024

**Presenting complaint:**

Abdominal pain x 3 months

**INDICATION FOR ADMISSION:**

Evaluation and Treatment of Presenting Complaints

**HISTORY**

Mr. Patient AA, 17 year old male, with no known comorbidities, presented with complaints of abdominal pain for last 3 months. He had his index presentation at an outside centre in May 2024 and was found to have acute pancreatitis. Episode resolved within 2 days and he was discharged in satisfactory condition. In July 2024, he had recurrent attack of pain following which he developed an abdominal distension. He was referred to ILBS for further management and was admitted. Imaging revealed a disconnected pancreatic duct syndrome with 2 collections – lesser sac and peripancreatic. PCD was inserted and he was discharged with PCD in situ and on LAR injections. Currently he has been admitted for follow up of the same. PCD output is nil. He had no history of trauma, drugs, alcohol prior to the presentation. He is now admitted for further evaluation and management.

**EXAMINATION:**

Pt. was conscious, oriented to time place and person.

BP: - 125/80 mm Hg, Pulse: - 80/min, RR: - 14/min and afebrile.

Pallor-, Icterus-, Cyanosis-, Clubbing-, Pedal edema (pitting type) - , LNP-, JVP normal

On systemic examination

Respiratory system:--B/L vesicular breathing, B/L airway equal air entry, no wheeze, no crepts

Cardiovascular system: - S1 S2 normal, no murmurs

CNS: - Conscious and oriented with no sensorimotor deficit,

Per Abdomen examination

ON INSPECTION: - Scaphoid abdomen, umbilicus central and inverted, skin over the abdomen with no visible venous prominences, no visible pulsations. Left sided PCD drain present but with nil output.

ON PALPATION: - Soft, tenderness - epigastrium, liver non palpable. Spleen non- palpable. No guarding, no rigidity, no rebound tenderness.

ON PERCUSSION: - Tympanic note with fluid present

ON AUSCULTATION: Normal bowel sounds present

**SYSTEMATIC REVIEW:**

Patient was admitted with above mentioned complaints. His initial lab data revealed is attached at the end of the summary. CECT abdomen was done on 14/10/2024 with report awaited, provisionally disrupted pancreatic duct with peri-pancreatic collection and lesser sac – 3x 4 cm collection. ERCP + PD stenting was done on 15/10/2024. PCD was removed on 15/10/2024. The nature of the disease and long term prognosis was explained in great detail to the patient and the patient relatives. He is discharged in hemodynamically stable condition and advised to follow up.

**TREATMENT:**

2100 Kcal/Day + 90 Grams of Protein/Day + fat restricted diet

TAB TAXIM-O 200 MG PO BD 1-0-1 FOR 5 DAYS

CAP HENZOVIT 1 TAB PO OD

CAP PANLIPASE 25K PO TDS WITH MEALS

PEPTAMEN POWDER 2 SCOOPS PO TDS

TAB PANTOP 40 MG PO OD

Review after 30 days in Hepatology OPD with CBC, LFT, RFT report

IN CASE OF DECREASE IN URINE OUTPUT, ALTERED SENSORIUM, BLEEDING, FEVER, NEW ONSET COUGH AND REVIEW IN ILBS EMERGENCY ON SOS BASIS.

### Summary 28

EHBO with Hilar Mass

- Etiology - Benign Biliary Stricture
  - ? IgG4 related (IgG4 – 2.74)
  - ?? Inflammatory stricture
- Primary Confluence and Secondary Right Confluence with involvement of RAD
- Periportal / Portocaval / Peripancreatic Lymph Node present
- Focal Acute Pancreatitis – Head and Uncinate Process
- FNAC from biliary stricture – 30/11/2024 – No e/o granulomatous or neoplastic pathology
- Brush Cytology from biliary stricture – 07/12/2024 - No e/o granulomatous or neoplastic pathology

S/P- Right and Left PTBD inserted on 30/11/2024, removed on 11/12/2024

S/P ERCP + Rendezvous Procedure on 11/12/2024 🡪 stent in RHD, LHD

CEA – 0.5, CA19.9 – 1.2

**Co Morbidities** – PSVT S/P RFA in July 2024

**CURRENT ISSUES:**

- Progessive Bilirubinemia (TB/DB/IB – 21.64/7.7/13.9 🡪 11.32/5.8/5.5) s/p Right and Left PTBD
- Acute Cholangitis 🡪 Resolved
- Bilioma s/p PCD inserted on 09/12/2024 🡪 s/p PCD removed on 12/12/2024

**PRESENTING COMPLAINTS:**

- Jaundice x 2 months
- Weight Loss x 2 months

**INDICATION FOR ADMISSION:**

Evaluation and management of symptoms

**HISTORY:**

Mr Patient AB is a 19 year old gentleman with known co morbidities of PSVT for which he underwent RFA in July 2024. He had his index presentation at an outside centre 2 months ago. He had first noticed a yellowish discolouration of his eyes and urine, which was insidious in onset and progressive. It was associated with itching and clay-coloured stools. He was evaluated outside and biopsy taken revealed no signs of malignancy. Jaundice worsened, and he was referred to ILBS for further management. There is no h/o, cough, altered bowel habits, hematemesis, and malena, burning micturition, altered sensorium or decreased urine output. There is no h/o any intoxications, indigenous medications, other major surgeries, blood transfusions or IV drug abuse prior to onset of the disease. There is no h/o DM/HTN/CAD/TB/COPD.

**EXAMINATION**

Pt. was conscious, oriented to time place and person.

BP: - 106/70 mm Hg, Pulse: - 86/min, RR: - 14/min and afebrile.

Pallor+, Icterus++, Cyanosis-, Clubbing-, Pedal edema (pitting type) - , LNP-, JVP normal

On systemic examination

Respiratory system:--B/L vesicular breathing, B/L airway equal air entry, no wheeze, no crepts

Cardiovascular system: - S1 S2 normal, no murmurs

CNS: - conscious and oriented with no sensorimotor deficit,

Per Abdomen examination

ON INSPECTION: - Non distended abdomen, umbilicus central and inverted, skin over the abdomen is stretched with no visible venous prominences, no visible pulsations.

ON PALPATION: - soft, non-tender, GB palpable. No guarding, no rigidity, no rebound tenderness.

ON PERCUSSION: - Tympanic note with no fluid present

ON AUSCULTATION -normal bowel sounds present,

**SYSTEMIC REVIEW**

Patient was admitted with above mentioned complaints. His examination findings were as mentioned above. Blood and Urine Cultures revealed no growth, and Urine Routine was unremarkable. S. PCT – 0.57. He was initiated on IV antibiotics, nutritional therapy and other supportive medications. A PET-CT was done on 30/11/2024-

- Mild focal metabolic activity involving soft tissue thickening involving the primary hilar confluence with proximal dilation bilobar IHBRD and bilateral PTBD in-situ.
- Focal increased metabolic activity involving the head and uncinate process of pancreas with adjacent fat stranding -suggestive of focal pancreatitis.
- Above features are more in favour of IgG4 related disease. Advise FNAC correlation.
- Mild metabolic activity in few lymph nodes in periportal, portocaval and peripancreatic regions - likely inflammatory.
- No evidence of abnormal metabolic activity disease noted elsewhere.
- Contracted gallbladder with radiopaque sludge.

He underwent Right and Left PTBD inserted on 30/11/2024. ERCP + Rendezvous Procedure on 11/12/2024 with stent placed in RHD and LHD. In view of bilioma PCD was inserted on 09/12/2024, and subsequently removed on 12/12/2024. -	FNAC from biliary stricture on 30/11/2024 reported no e/o granulomatous or neoplastic pathology. Brush Cytology from biliary stricture taken on 07/12/2024 revelaed no e/o granulomatous or neoplastic pathology Proper diet and mobilization was followed throughout the course of admission. All vitals, temperature, RBS and other necessary parameters were checked regularly and managed appropriately. The nature of the disease and long term prognosis was explained in great detail to the patient. Now he has improved symptomatically and is being discharged in hemodynamically stable condition with the advice to follow up in OPD.

**ADVICE** **ON DISCHARGE (Duration for medications if any):**

2000 KCAL/DAY + 90 GRAMS PROTEIN/DAY, NORMAL DIET

TAB TAXIM-O 200MG BD	1-0-1 X 5 DAYS

TAB FLUCOS 200 MG PO OD FOR 5 DAYS

TAB ATARAX 25 MG PO HS

TAB UDCA 450 MG PO BD

CAP BETADEK PO OD

TAB SHELCAL 1 TAB PO OD

LAXOPEG SACHET BD. 1-0-1 (ENSURE 2-3 BOWEL MOTIONS/DAY)

IN CASE OF DECREASE IN URINE OUTPUT, ALTERED SENSORIUM, BLEEDING, FEVER, NEW ONSET COUGH AND REVIEW IN ILBS EMERGENCY ON SOS BASIS.

### Summary 29

**1.)** PHTN - (Bleeder, Small High Risk Esophageal Varices, EVL done, Mild PHG, GAVE - APC done (19/07/2024)

CLD - NASH related (K75.8)

Decompensated - Ascites / AVB

**2.)** Obstructive Uropathy **-** Left Renal Stone causing Left Moderate to Gross Hydroureteronephrosis

- S/P Left DJ Stenting - 27/07/2024

- s/p Left RIRS with DJ stenting – 07/10/2024

**CURRENT ISSUE**

- Obstructive Uropathy s/p Left RIRS with DJ stenting – 07/10/2024

- Post-operative Sepsis with Septic Shock (Resolved), S. PCT – 94.7 🡪17.2

- AKI Stage II – Sepsis related, (Resolved – Creatinine 1.45 🡪 0.71)

- Sepsis-induced Liver Dysfunction

- Thrombocytopenia – Sepsis related (Platelet count 65000 🡪15000 🡪60000)

**CO-MORBIDITY** - HTN / T2 Diabetes Mellitus

**PRESENTING COMPLAINTS:**

Hematuria x 7 days

**INDICATION FOR ADMISSION:**

Evaluation and management

**HISTORY:**

Mrs Patient AC is a 65 years old lady, and known diabetic and hypertensive. She had her index presentation in Nov 2023 as fever evaluate found to have CLD. In June 2024 patient had abdominal distension, fatigue /weakness and malena - S/P APC done 08/06/2024. She was admitted in July 2024 in view of sepsis secondary to left renal stone causing obstructive uropathy. Left sided DJ stenting was done on 27/07/2024 and she was discharged in hemodynamically stable condition. She was planned for an elective Left RIRS. She has currently presented for Left RIRS. There is no h/o vomiting, cough, altered bowel habits, burning micturition, altered sensorium or decreased urine output. There is no h/o any intoxications, indigenous medications, or IV drug abuse prior to onset of the disease. There is no h/o - COPD/ Thyroid disease/CKD.

**EXAMINATION:**

Pt. was conscious, oriented, a febrile.

BP 110/58 mm Hg	Pulse 92/min	RR 18/min

Pallor-, Icterus-, Cyanosis-, Clubbing-, Pedal edema+, LNP-, JVP normal,

CVS S1 S2 normal, no murmurs

Chest B/L normal air entry & vesicular breathing, no adventitious sounds

CNS Patient conscious, oriented, no sensorimotor deficit.

P/A ON INSPECTION – Non-distended, umbilicus central and inverted, no visible venous prominences, no visible pulsations.

ON PALPATION -soft, non-tender, liver and spleen non-palpable. No guarding, no rigidity, no rebound tenderness.

ON PERCUSSION: Tympanic note, no	free fluid present.

ON AUSCULTATION: normal bowel sounds present, no bruits.

**SURGERY NOTES:**

**Patient Name: Mrs. Patient AC	Age/gender: 65/F**

**In time: 1:30PM	Starting Time: 2:15PM	close time: 3: 45PM	out time: 4:05 PM**

**Date of surgery: 7-10-2024	Duration of surgery: 1 HOURS; 30 mins**

**Pre-Operative Diagnosis**: Left Renal calculi

**Operative Diagnosis:** Left Renal calculi

**Procedure Planned:** Retrograde Intrarenal Surgery (RIRS) with dj stenting under SA

**Procedure Performed:** Retrograde Intrarenal Surgery (RIRS) with dj stenting under SA

**Anesthesia:** SA	**Epidural:** no

**Cystoscopic findings:-**

Meatus Normal, Urethra Normal,

Left Ureteric Orifice -distal end of DJ stent seen and orifice seen dilated.

Stone noted in left puj

No E/o any Growth and Calculus in Urinary Bladder

**Flexible Ureteroscopic findings:-**

There is normal mucosa of ureter. Stone of approx. 10mm seen in the renal pelvis (left)

**Patient Position:** Lithotomy.

**Procedure Details:**

Patient positioned and part prepared, painted and draped under GA.

Cystoscopy done and dj stent removed from left ureteric orifice and findings noted above.

Left ureter cannulated with guidewire and confirmed with C-Arm.

Ureter access sheath (9.5-11 Fr) passed over guide wire into the left ureter till mid Ureteric level and fixed.

Flexible ureteroscope passed and stone seen at pelvi-ureteric junction and dusted & fragmented using laser fiber & stone free status confirmed by C ARM.

6/16 DJ stenting done and position confirmed using c arm

PUC (16 Fr) placed and balloon inflated to 10cc. Clear urine colour noted.

WHO safety checklist was ensured.

**Estimated Blood Loss: 40ml**

**Extubated/ Shifted on ventilator:** Extubated

**SYSTEMATIC REVIEW:**

Patient AC was admitted in Urology with the above mentioned complaints. Left sided RIRS with DJ stenting was done on 07/10/2024. In the immediate post-operative period, she underwent episodes of hypotension. Fluid resuscitation was initiated and inotropic support was required. She was shifted to the ICU for closer monitoring. Blood and Urine Cultures revealed no growth, and Urine Routine revealed 8-10 leukocytes with full field RBCs. S. PCT – 94.7. She was started on inotropes, fluid resuscitation, and antibiotics were upgraded. She clinically improved, inotropes were tapered, and she was shifted to the ward on POD - 3. There were no further episodes of fever. Nephrology advice taken and advice followed. Antibiotics were downgraded, and she was managed conservatively with IV fluids, nutritional therapy & other supportive treatments. Further urine and blood cultures revealed no growth. Urology advices were taken and advice followed. She recovered symptomatically following the management and now she is being discharged in stable hemodynamic state with advice of follow up in OPD.

**PLAN / ADVICE AT DISCHARGE (Including duration of medication if any):**

2100 KCAL/DAY + 90 GRAMS PROTEIN/DAY, LOW SALT (<2GRAMS/DAY)

TAB FAROPENAM 200 MG PO BD FOR 5 DAYS

TAB LEVOFLOX 500 MG PO OD X 5 DAYS

TAB B COMPLEX PO BD

TAB FOLIC ACID PO BD

TAB MONTAIR LC PO HS X 5 DAYS

SYP LACTIHEP 30ML PO HS  0-0-1 (ENSURE 2-3 BOWEL MOTIONS/DAY)

TAB EPTUS 25 MG PO OD (MONITOR KFT WEEKLY)

TAB DYTOR 5 MG PO BD (8 AM AND 2 PM) (MONITOR KFT WEEKLY)

HEPSURE SACHET PO OD

SYRUP ALKASOL 2 TSP PO HS

TAB URIMAX 0.4 MG PO HS

REVIEW IN UROLOGY OPD ON SATURDAY – 19/10/2024 – 2PM

IN CASE OF DECREASE IN URINE OUTPUT, ALTERED SENSORIUM, BLEEDING, FEVER, NEW ONSET COUGH AND REVIEW IN ILBS EMERGENCY ON SOS BASIS.

### Summary 30

**DISCHARGE ON REQUEST**

PHTN (Non-Bleeder, Small Low Risk Esophageal Varices with mild PHG – 05/09/2024)

Acute Severe DILI (h/o Amoxicillin + Clavulanic Acid)

HVPG – 14 mmHg - 05/11/2024

Biopsy – Severe Acute Hepatitis with Panacinar Necrosis

s/p PLEX –

- 09/11/2024 – Normal Volume
- 11/11/2024 - Normal Volume
- 13/11/2024 - High Volume
- 16/11/2024 - Normal Volume

On Steroid Therapy started on 20/11/2024

CTP - 8 ,CHILD - B, MELD Na – 22

**Comorbidities –** Hypertension, Obesity

**Current Issues**:

- Severe DILI
  - s/p PLEX 3 sessions (TB 31.59 🡪13.04)
  - Rising Bilirubin after 4^th^ session of PLEX (TB 13.04 🡪 15.98) 🡪 Started on Steroids on 20/11/2024

**PRESENTING COMPLAINTS**

Yellowish discolouration of eyes since 1 month

**INDICATION FOR ADMISSION:**

Evaluation and management of symptoms

**HISTORY:**

Mr Patient AD is a 41 years old gentleman with comorbidities of Hypertension since 15 years, for which he is taking Telmisartan (irregular compliance). He is obese with a max weight of 95 kg, and has no addictions. His index presentation was 1 month ago with a history of high grade fever which lasted for 1 day. He self-medicated with Paracetamol and fever subsided. He then developed a progressive swelling of feet with reddish discolouration and presented at an outside centre. He was prescribed DEC and a course of Amoxicillin + Clavulanic Acid with suspicion of filariasis. One week after completion of course, he developed new onset jaundice, which was insidious in onset and progressive. It was not associated with itching or clay-coloured stools. He was re3ferred to ILBS for management of the same. There is no h/o vomiting, cough, abdominal pain, altered bowel habits, hematemesis, and malena, burning micturition, altered sensorium or decreased urine output. There is no history of loss of weight or appetite. There is no history of CAM intake. There is no h/o any intoxications, major surgeries, blood transfusions or IV drug abuse prior to onset of the disease. There is no h/o DM/CAD/TB/COPD/Thyroid disorder.

**EXAMINATION**

Pt. was conscious, oriented to time place and person.

BP: - 116/70 mm Hg, Pulse: - 86/min, RR: - 14/min and afebrile.

Pallor-, Icterus++, Cyanosis-, Clubbing-, Pedal edema (pitting type) + , LNP-, JVP normal

On systemic examination

Respiratory system:--B/L vesicular breathing, B/L airway equal air entry, no wheeze

Cardiovascular system: - S1 S2 normal, no murmurs

CNS: - conscious and oriented with no sensorimotor deficit,

Per Abdomen examination

ON INSPECTION: - umbilicus central and inverted, no visible swellings or pulsations.

ON PALPATION: - soft, non-tender, Liver palpable 3 cm below right costal margin. Spleen not palpable. No guarding, no rigidity, no rebound tenderness.

ON PERCUSSION: -No appreciable free fluid

ON AUSCULTATION normal bowel sounds present

**SYSTEMATIC REVIEW:**

Patient was admitted with above mentioned complaints and baseline investigations were sent. His examination findings were as mentioned above. His initial lab data and latest lab data is included at the end of the summary. Blood C/S and Urine C/S showed no bacterial growth. Serum PCT – 0.43. Autoimmune Profile IgG – 18.2. Anti HEV IgM and Anti HAV IgM negative. ELF Score 13.91. UGIE screening was done on 05/09/2024 - Small Low Risk Esophageal Varices with mild PHG. HVPG and TJLB was done on 05/11/2024 – 14 mmHg. Biopsy – Severe Acute Hepatitis with Panacinar Necrosis. HD Cath was inserted and he underwent 4 sessions of PLEX on 09/11, 11/11, 13/11 and 16/11. Bilirubin initially decreased post PLEX, however, rising trend ensued after 4^th^ session of PLEX. He was then started on steroid course on 20/11/2024. In view of blood-tinged feces and painful defecation, he underwent a sigmoidoscopy on 12/11/2024 which revealed acute anal fissure and managed accordingly. Proper diet and mobilisation was followed throughout the course of admission. All vitals, temperature, RBS and other necessary parameters were checked regularly and managed appropriately. The nature of disease and its prognosis has been clearly explained to patient and his attendants and also counselled for liver transplantation. However, no **prospective donor available at present.** He is currently **discharged on request**, in hemodynamically stable condition with advice to follow in OPD.

**PLAN / ADVICE AT DISCHARGE (Including duration of medication if any):**

2400 KCAL/DAY + 90 GRAMS PROTEIN/DAY, LOW SALT (<2GRAMS/DAY)

PERSONAL PROTECTION AND HYGIENE AS ADVISED.

AVOID SOCIAL GATHERINGS, WEAR MASK REGULARLY.

Tab WYSOLONE 40 mg PO OD, 1-0-0 at 10 am for 7 days

TAB FAROPENAM 200 MG PO BD FOR 7 DAYS

TAB LEVOFLOX 500 MG PO OD FOR 7 DAYS

TAB FLUCOS 200 MG PO OD FOR 7 DAYS

TAB CARDIVAS 3.125 MG PO BD (DO NOT GIVE IF HR < 55 OR BP < 90/60 MMHG)

CAP HENZOVIT 1 TAB PO OD

TAB UDILIV 450 MG PO BD

HEPSURE SACHET PO BD

ANOVATE OINTMENT L/A BD

SITZ BATH TDS AS ADVISED

TAB ATARAX 25 MG PO SOS

SYP LACTIHEP 30ML PO HS  0-0-1 (ENSURE 2-3 BOWEL MOTIONS/DAY)

MOVICOL SACHET 1 SACHET PO SOS

TAB SHELCAL 1 TAB PO OD X 7 DAYS

TAB PANTOP 40 MG PO OD BBF X 7 DAYS

**PLAN:** REPEAT KFT, LFT, INR, CBC AFTER 7 DAYS AND FOLLOW UP IN HEPATOLOGY OPD OR 4^TH^ FLOOR PRIVATE - PHASE II

IN CASE OF DECREASE IN URINE OUTPUT, ALTERED SENSORIUM, BLEEDING, FEVER, NEW ONSET COUGH AND REVIEW IN ILBS EMERGENCY ON SOS BASIS.

### Summary 31

Acute-on-Chronic Liver Failure (K72.1) ------ Acute: Hepatitis B Reactivation + DILI

Chronic: HBV Cirrhosis (B18.9)

PHTN (Non-Bleeder, Small Low Risk Esophageal Varices, mild PHG as of 02/09/2024)

k/c/o Ca Pancreas s/p NACT - 4 cycles of FOLFOX, s/p Distal Pancreaticosplenectomy, s/p NACT - 3 cycles of FOLFOX

CAP 223 LSM 62.1

CTP - 10, CHILD - C, MELD Na - 28

HVPG 12 mmHg, Biopsy – Chronic Hepatitis with mild activity and superimposed DILI

Ishak’s modified HAI – 6/18; Fibrosis – 5/6

**Comorbidities:** T2 Diabetes Mellitus

**Current Issues:** 1.) Reactivation of HBV (DNA – log 7, HBe reactive) + DILI

2.) ? Suspected TB

3.) Peripheral Neuropathy: ?Diabetic ?Chemotherapy-Induced

**PRESENTING COMPLAINTS**

Yellowish discolouration of eyes since 15 days

**INDICATION FOR ADMISSION:**

Evaluation and management of symptoms

**HISTORY:**

Mr Patient AE is a 61 year old gentleman, who is a known case of CA Pancreas. He had his index presentation in December 2023 when he was detected to have a lesion in bodyu of pancreas. He was also detected to have a Chronic Liver Disease with Portal Hypertension which was Compensated.

He underwent a distal pancreaticosplenectomy (Anterior RAMPS) in January 2024. A liver biopsy was done intra-op which revealed macrovascular steatosis, ballooning degeneration and portal tracts showing moderate chronic inflammation with interface hepatitis; Bridging fibrosis with fibrosis score 3/6. He underwent cycles of chemotherapy (Oxaliplatin based) and was started on oral chemotherapy (5-Fluorouracil based). A recent biopsy in September 2024 revealed Zone 3 and Zone 2 hepatocanalicular cholestasis, cholestatic rosettes, moderate ballooning regeneration. Mixed lobular infiltrates (Eo snd spotty necrosis) and apoptotic bodies. Fibrosis 5/6.

Currently he has presented with a history of new onset jaundice since 25 days and abdominal distension for 5 days. Jaundice was not associated with itching or clay-coloured stools. There was no h/o altered sensorium or acute bleeds. There is no h/o any intoxications, major surgeries, blood transfusions or IV drug abuse prior to onset of the disease. There is no h/o DM/HTN/CAD/TB/COPD/Thyroid disorder.

**EXAMINATION**

Pt. was conscious, oriented to time place and person.

BP: - 116/70 mm Hg, Pulse: - 86/min, RR: - 14/min and afebrile.

Pallor-, Icterus+ , Cyanosis-, Clubbing-, Pedal edema (pitting type) - , LNP-, JVP normal

On systemic examination

Respiratory system:--B/L vesicular breathing, B/L airway equal air entry, no wheeze

Cardiovascular system: - S1 S2 normal, no murmurs

CNS: - conscious and oriented with no sensorimotor deficit,

Per Abdomen examination

ON INSPECTION: - mild distended abdomen, no visible swellings or pulsations.

ON PALPATION: - soft, non-tender, Liver palpable 2 cm below right costal margin. Spleen not palpable. No guarding, no rigidity, no rebound tenderness.

ON PERCUSSION: - Dull note, free fluid present

ON AUSCULTATION normal bowel sounds present

**SYSTEMATIC REVIEW:**

Patient was admitted with above mentioned complaints and baseline investigations were sent. His examination findings were as mentioned above. His initial lab data and latest lab data is included at the end of the summary.

PIVKA II was 1649.17, AFP was 7.5 and CA 19-9 - 71.5. TNF Alpha – 28.2 / HBsAG Quantification – 415 / HBeAg Reactive / Anti- HB Core reactive and Anti-HBS Non-Reactive. HVPG and Liver biopsy (TJLB) was done on 03/09/2024. HVPG – 12 mmHg.

Biopsy revealed - Chronic Hepatitis with mild activity and possibly coexistent drug-induced liver injury (DILI) with advanced fibrosis (probable cirrhosis). Ishak's Modified HAI- 4/18. Fibrosis – 5 / 6. A review of PET-CT records was done. Reported as status-post distal pancreatectomy and splenectomy with no focal metabolically active lesion in the post-operative bed. Mild metabolically active mediastinal lymph nodes with right-sided mild pleural effusion - likely infective in aetiology. As compared to previous PET CT dated 03/06/2024, there is mild increase in size of mediastinal lymph nodes with no significant change in metabolic activity. A Medical Oncology consult advised no active intervention at present in view of deranged LFTs. He complained of a persistent pain and numbness of his extremities, which had worsened post-chemotherapy. An NCV was done which revealed a mixed (motor + sensory) polyneuropathy. A neurology consult was done and advice followed. The nature of the disease and long term prognosis was explained in great detail to the patient and the patient relatives. Proper diet and mobilisation was followed throughout the course of admission. All vitals, temperature, RBS and other necessary parameters were checked regularly and managed appropriately. He is discharged in hemodynamically stable condition with advice to follow up in OPD.

**PLAN / ADVICE AT DISCHARGE (Including duration of medication if any):**

2000 Kcal/day + 90 grams protein/day

TAB TAF 25 MG PO OD

TAB ENTACAVIR 0.5 MG PO OD

TAB LASILACTONE (20/50) **½ TABLET** PO OD

TAB MIDODRINE 5 MG PO TDS

TAB GABAPENTIN PO HS

TAB CLONAZEPAM 0.25 MG PO SOS

BRANVIO SACHET PO BD

ENSURE DM POWDER 2 SCOOP PO TDS

HEPSURE SACHET PO OD

INJ LANTUS 14 U S/C HS

STRICT RBS MONITORING AND GLYCEMIC CONTROL AS ADVISED

Review in Hepatology OPD with CBC/LFT/KFT/INR reports after 1 week. REVIEW IN VIRTUAL OPD AFTER 4 DAYS.

WEEKLY TEST – HBV DNA AND HBSAG

IN CASE OF DECREASE IN URINE OUTPUT, ALTERED SENSORIUM, BLEEDING, FEVER, NEW ONSET COUGH AND REVIEW IN ILBS EMERGENCY ON SOS BASIS.

### Summary 32

Non Bleeder, No esophageal varices, No signs of PHTN - 27/05/2023

HVPG 5 mmHg - 29/05/2023

CLD - Ethanol related (Last intake - 1 days back ) **(ICD10 - K70.9)**

Alcohol Use Disorder with Alcohol Withdrawal Syndrome

CAP – 251	LSM – 15.3	SSM – 24.2

**Comorbidities:** AUD x 5 years ; Strong Recidivsim

**CURRENT ISSUES:**

- Alcohol Withdrawal Syndrome
- Transaminitis (AST/ALT – 791 / 168)
  - s/p USG guided Liver Biopsy - 26/12/2024 - Report Awaited

**PRESENTING COMPLAINTS:**

- Tremors x 1 days
- Disturbed sleep pattern

**INDICATION FOR ADMISSION:**

Evaluation & management of the presenting complaints

**HISTORY:**

Mr. Patient AF is a 41 years old male with h/ o chronic ethanol use (LI- 1 days back ), with no comorbidity . Patient has h/o seizure previously after a day of abstinence from alcohol, c/o tremors and disturbed sleep pattern. He was admitted outside and managed conservatively as CLD likely ethanol related ( with signs and symptoms of withdrawal). However, alcohol use continued and he has been admitted for further evaluation of ALD. There is no h/o abdominal pain, vomiting, cough, altered bowel habits, hematemesis, malena, burning micturition, or decreased urine output. With the above complaints, he got admitted for further evaluation and management. There is no h/o any intoxications, indigenous medications, blood transfusions or IV drug abuse prior to onset of the disease. There is no h/o HTN/CAD/TB/COPD/Thyroid disorders.

**EXAMINATION:**

Pt. was conscious, oriented, afebrile.

BP 122/ 80 mm Hg Pulse 84/min	RR -20/min

Pallor-, Icterus-, Cyanosis-, Clubbing-, Pedal edema-, LNP-, JVP normal

CVS -S1 S2 normal, no murmurs

Chest- B/L normal air entry & vesicular breathing, no adventitious sounds

CNS -Patient conscious, oriented, no sensorimotor deficit.

P/A ON INSPECTION non-distended, umbilicus centrally located and inverted. No visible superficial veins +, no visible pulsations.

ON PALPATION -soft, non tender, liver - non palpable spleen - non palpable. No guarding, no rigidity, no rebound tenderness.

ON PERCUSSION -tympanic note, with no free fluid present.

ON AUSCULTATION- normal bowel sounds present, no bruits.

**SYSTEMATIC REVIEW:**

Patient AF was admitted with the above mentioned plan. His examination findings were as mentioned above. He was managed with IV antibiotics , IV albumin, nutritional therapy & other supportive measures. Psychiatry opinion was taken in view of AUD and AWS and their advice was followed and subsequently his improved symptomatically. Patient was treated with IV antibiotics, nutritional support and other supportive medication and treatment. Repeat Psychiatry opinion was taken in view of recidivism and their advice was followed. AUD counselling done. He was counselled and planned for FMT and risks and benefits for the same was explained. In view of transaminitis, he underwent a USG guudeed liver biopsy on 27/12/2024 – Report Awaited. He is now planned for nutritional optimization and improvement in performance status. Proper diet and mobilisation was followed throughout the course of admission. All vitals, temperature, RBS and other necessary parameters were checked regularly and managed appropriately. He is now discharged in hemodynamically stable condition with following advice to follow in OPD.

**PLAN / ADVICE AT DISCHARGE (Including duration of medication if any):**

2100 KCAL/DAY + 90 GRAMS PROTEIN/DAY, LOW SALT (<2GRAMS/DAY), DIABETIC DIET

T.  TAXIM O 200MG PO BD	1-0-1 X 5 DAYS THEN STOP

TAB CILACAR 10 MG PO BD (DO NOT GIVE IF BP < 90/60 AND HR < 55)

TAB CIPLAR LA 20 MG PO BD	**2-0-1**	(DO NOT GIVE IF BP < 90/60 AND HR < 55)

TAB ARKAMINE 0.1 MG PO TDS

T. LEVIPIL 500MG PO BD	1-0-1

T. ZOLFRESH 10MG PO HS 0-0-1

T. ALCOMAX 1 TAB PO OD	1-0-0

TAB NALTREXONE 25 MG PO OD HS

TAB LIBRIUM 25 MG PO SOS

C. HENZOVIT 1 CAP PO OD	1-0-0

SYP LACTIHEP 15 ML PO HS	0-0-1 (ENSURE 2-3 BOWEL MOTIONS/DAY)

REVIEW IN HEPATOLOGY / PSYCHIATRY OPD WITH CBC/LFT/KFT/INR REPORTS AFTER 4 WEEKS.

IN CASE OF ALTERED SENSORIUM, DECREASE IN URINE OUTPUT, BLEEDING, FEVER, NEW ONSET COUGH AND REVIEW IN ILBS EMERGENCY ON SOS BASIS.

### Summary 33

PHTN - Non Bleeder, Small Esophaeal Varices – 01/07/2024

Chronic Liver Disease - HVOTO – Chronic BCS (ICD - I82.0)

- Near-complete occlusion of IVC with non-visualization of all 3 Hepatic Veins
- Accessory Hepatic Vein seen with multiple collaterals
- s/p IVC Angioplasty on 05/10/2024

CTP-7, CHILD-B, MELD 8

**PRESENTING COMPLAINTS:**

Abdominal distension x 4 months

**INDICATION FOR ADMISSION:**

Evaluation and the management of presenting symptoms

**HISTORY:**

Mrs. Patient AG is a 32 year old lady with no known comorbidities. She had her index presentation at an outside centre in June 2024 when she presented with a progressive abdominal distension and abdominal pain. On evaluation she was found to have ascites and CECT diagnosed a chronic liver disease. UGIE screening was done on 01/07/2024 at the outside centre and revealed Small Esophaeal Varices. She was referred to ILBS for further management. On 30/09/2024, HVPG + TJLB was attempted but procedure abandoned due to unsuccessful cannulation, due to a suspicious narrowing at IVC – RA junction. There is no complaint of jaundice, fever, cough, loose stools, vomiting, hematemesis, melena, burning micturition or decreased urine output. He is not a case of T2DM, HTN, COPD, CAD, Thyroid or renal disorders.

**GENERAL EXAMINATION:**

Pt. was conscious, oriented, afebrile

BP- 104/78 mm Hg     Pulse - 120 / min	RR - 16 / min

Pallor -, Icterus+, Cyanosis-, Clubbing-, Pedal edema - , LNP-, JVP normal

CVS -S1 S2 normal, no murmurs

Chest- B/L air entry present, no added sounds

CNS -conscious, oriented to time and place

P/A examination

ON INSPECTION - mildly Distended, umbilicus central and inverted, no visible venous prominences,

no visible pulsations.

ON PALPATION - Soft, non-tender, liver and spleen non-palpable. No guarding, no rigidity, no rebound tenderness.

ON PERCUSSION - tympanic note, no free fluid

ON AUSCULTATION - Normal bowel sounds present, no bruits.

**SYSTEMATIC REVIEW:**

A CECT Abdomen was performed with report awaited, and UGI Endoscopy done on 27/08/2024 revealed small high risk oesophageal varices with mild PHG. A complete prothrombotic gene profile was sent (report awaited). TJLB was done on 28/08/2024 and reported as cirrhosis with features suggestive of chronic HVOTO. Right Hepatic Vein Angioplasty was performed on 28/08/2024 and post-procedure RHV and IVC showed normal colour flow. The procedures were tolerated well. There were no post procedural complications. The nature of the disease and long term prognosis was explained in great detail to the patient and the patient relatives. He is now planned for nutritional optimization and improvement in performance status. Proper diet and mobilisation was followed throughout the course of admission. All vitals, temperature, RBS and other necessary parameters were checked regularly and managed appropriately. He recovered symptomatically following the management and is discharged in hemodynamically stable condition with the advice to follow ups in OPD.

**PLAN / ADVICE AT DISCHARGE (Including duration of medication if any):**

1800 Kcal/day + 80 grams protein/day, low salt (<2grams/day)

DO NOT TAKE ANY MEDICINES FROM OUTSIDE WITHOUT CONSULT

**TO MAINTAIN INR BETWEEN 2.5 AND 3**

INJ. CLEXANE 0.4 MG S/C BD TO CONTINUE

SYP LACTIHEP 30ML PO SOS 	(ENSURE 2-3 BOWEL MOTIONS/DAY)

TAB TAXIM-O 200MG BD	1-0-1 X 5 DAYS

ECONORM SACHET PO BD FOR 7 DAYS

TO BE FOLLOWED UP IN HEPATOLOGY OPD WITH **USG ABD DOPPLER** / CBC/LFT/KFT/INR/ CECT REPORTS AFTER **1 MONTH**

**PLAN: USG DOPPLER AFTER 1 MONTH – IF HV NOT PATENT, PLAN HEPATIC VEIN PUNCTURE**

IN CASE OF DECREASE IN URINE OUTPUT, ALTERED SENSORIUM, BLEEDING, FEVER, NEW ONSET COUGH AND REVIEW IN ILBS EMERGENCY ON SOS BASIS.

### Summary 34

Portal hypertension (Non-bleeder/ Small Low Grade Esophageal Varices, Severe PHG, GAVE 26/09/2024)

Cirrhosis - Ethanol related (K70.3) (Relapse with Last Intake - 2 days back)

Decompensated -Ascites (controlled)

Prior ACLF - May 2024

s/p UGIE + Fecal Microbiata Transplantation - 26/09/2024

Liver Biopsy: Severe Alcoholic Steatohepatitis with Bridging Fibrosis with AHHS- 7/9

**Comorbidities:** HTN

**CURRENT ISSUES:**

Alcohol Associated Hepatitis 🡪 Resolving on Conservative management (TB – 3.84🡪 2.63) (Mdf – 32.8)

**PRESENTING COMPLAINTS**

- Generalized Weakness
- Abdominal Distension
- Pain abdomen + Vomiting

**INDICATION FOR ADMISSION:**

Evaluation and management of symptoms

**HISTORY:**

Mr. Patient AH is a 38 years old gentleman with co morbidities of HTN and AUD. He is a known ethanol user with LI - 2 days prior to this admission. He had his index presentation in May 2024 with jaundice and abdominal distension. With this clinical picture he was admitted at ILBS and underwent liver biopsy on 24/05/24 which showed Severe Alcoholic Steatohepatitis with Bridging Fibrosis with AHHS- 7/9 and was managed on nutrition and subsequently discharged in hemodynamically stable condition. He was again admitted with alcohol withdrawal symptoms. Now he has been again admitted in view of alcohol relapse and increasing bilirubin. There is no h/o cough, altered bowel habits, hematemesis, and malena, burning micturition, altered sensorium or decreased urine output. There is no h/o any indigenous medications, major surgeries, blood transfusions or IV drug abuse prior to onset of the disease. There is no h/o CAD/ TB/ COPD/ Thyroid or renal disorders.

**EXAMINATION**

Pt. was conscious, oriented to time place and person.

BP: - 116/70 mm Hg, Pulse: - 86/min, RR: - 14/min and afebrile.

Pallor-, Icterus+ , Cyanosis-, Clubbing-, Pedal edema (pitting type) + , LNP-, JVP normal

On systemic examination

Respiratory system:--B/L vesicular breathing, B/L airway equal air entry, no wheeze

Cardiovascular system: - S1 S2 normal, no murmurs

CNS: - conscious and oriented with no sensorimotor deficit,

Per Abdomen examination

ON INSPECTION: - distended abdomen, no visible swellings or pulsations.

ON PALPATION: - soft, non-tender, Liver and Spleen not palpable. No guarding, no rigidity, no rebound tenderness.

ON PERCUSSION: - Dull note, free fluid present

ON AUSCULTATION normal bowel sounds present

**SYSTEMATIC REVIEW:**

Patient was admitted with above mentioned complaints and baseline investigations were sent. His examination findings were as mentioned above. His initial lab data and latest lab data is included at the end of the summary.Blood and Urine Cultures revealed no growth, and Urine Routine was unremarkable. S. PCT – 0.37. Tap was done and Fluid examination revealed straw coloured fluid with WBC - 59, N – 19% and L – 80%, Protein: 3.08, Glu: 110, Albumin: 1.69, and Gene Xpert: negative, Gram Stain negative. Cultures were negative.Psychiatry opinion was taken in view of AUD and AWS and their advice was followed and subsequently his improved symptomatically. Patient was treated with IV antibiotics, nutritional support and other supportive medication and treatment. Psychiatry opinion was taken in view of recidivism and their advice was followed. AUD counselling done.Conservative management was opted for, and a decreasing trend of bilirubin ensued. The nature of the disease and long term prognosis was explained in great detail to the patient. Proper diet and mobilisation was followed throughout the course of admission. All vitals, temperature, RBS and other necessary parameters were checked regularly and managed appropriately. Strict alcohol abstinence advised. He is discharged in hemodynamically stable condition with advice to follow up in OPD.

**ADVICE ON DISCHARGE (Duration of medications if any):**

2500 Kcal/day + 80 grams protein/day, low salt (<2grams/day), normal diet

TAB TAXIM-O 200MG BD	1-0-1 X 5 DAYS *

TAB CIPLAR LA 20 MG PO BD 1-0-1 (STOP IF HR<55,BP <90/60)

TAB TELMA AM 40 MG PO OD (STOP IF HR<55,BP <90/60)

TAB ALDACTONE 50 MG PO OD (MONITOR KFT WEEKLY)

TAB DYTOR 10 MG PO OD (MONITOR KFT WEEKLY)

TAB LEVIPIL 500MG PO BD 1-0-1

TAB ALCOMAX 300MG PO TDS 1-0-1

SYP LACTIHEP 30ML PO HS  0-0-1 (ENSURE 2-3 BOWEL MOTIONS/DAY)

CAP HEPKART 400 MG PO BD

TAB NUCICAL PLUS PO OD

CAP HENZOVIT 1 TAB PO OD

HEPSURE SACHET PO BD

TAB MELATONIN 1 TAB PO SOS / HS

TAB BACLOFEN 10 MG PO OD HS

HEPAMERZ SACHET 1 TDS 1-1-1

REVIEW IN HEPATOLOGY OPD WITH CBC/LFT/KFT/INR REPORTS AFTER 4 WEEKS.

PSYCHIATRY OPD FOLLOW UP AS ADVISED

IN CASE OF DECREASE IN URINE OUTPUT, ALTERED SENSORIUM, BLEEDING, FEVER, NEW ONSET COUGH AND REVIEW IN ILBS EMERGENCY ON SOS BASIS.

### Summary 35

Portal Hypertension - (Non bleeder, Grade II Esophageal Varices With No RCS - 13/02/23)

Cirrhosis ?AIH – (AMA M2 +++) (ICD - K76.8)

Decompensated With Jaundice

**Co-morbidities**: None

**Current issues: -**

- Persistent Jaundice (T.B./D.B/I.B. – 4.15 / 2 / 2.15)
- Thrombocytopenia, likely due to hypersplenism (Plt – 39,000)
- MRCP – 20/09/2024 -
  - Bilobar IHBRD show irregularity with appearance of beading on both sides (R >L).
  - In view of cirrhotic liver bile duct irregularity (small ducts) cannot be differentiated from primary / secondary cholangiopathy.
- s/p Colonoscopy 01/10/2024	-
  - Coagulopathy related ooze.
  - Normal Mucosa at ileum, caecum and colon.
  - External haemorrhoids seen.
  - Rectal biopsy not taken i/v/o coagulopathy.

**PRESENTING COMPLAINTS:**

Bleeding P/R

**INDICATION FOR ADMISSION:**

Evaluation and the management of the symptoms

**HISTORY:**

Mrs. Patient AI is a 33 years old lady, with no known comorbidities. She had her index presentation in February 2023 at an outside centre when she presented with generalized weakness and easy fatiguibility, and then developed a jaundice. She was evaluated outside and diagnosed with chronic liver disease, and was referred to ILBS. Thrombocytopenia, likely due to hypersplenism was revealed and she was planned for a splenic artery embolization, which was deferred in view of hyperbilirubinemia. She was discharged in satisfactory condition and has been on regular OPD follow up since. In view of persistent bilirubinemia in Spetember 2024, she underwent an MRCP on 20/09/2024 which revealed Bilobar IHBR show irregularity with appearance of beading on both sides (R >L). In view of cirrhotic liver bile duct irregularity (small ducts) cannot be differentiated from primary / secondary cholangiopathy. USG on 20/09/2024 Chronic liver disease with findings suggestive of portal hypertension (splenomegaly with prominent splenoportal axis and collaterals). She has had intermittent episodes of hematochezia – fresh blood. She has been admitted for a colonoscopy and further evaluation. There is no h/o fever, vomiting, cough, abdominal pain, abdominal distension, altered bowel habits, hematemesis, malena, altered sensorium, burning micturition or decreased urine output. With these complains she got admitted for further evaluation and management.There is no h/o any intoxications, indigenous medications, blood transfusions or IV drug abuse prior to onset of the disease. There is no h/o DM/HTN/CAD/TB/COPD/Thyroid disorders.

**EXAMINATION**

Pt. was conscious, oriented to time place and person.

BP: - 102/70 mm Hg, Pulse: - 86/min, RR: - 14/min and afebrile.

Pallor -, Icterus - , Cyanosis - , Clubbing - , Pedal edema- , LNP-, JVP normal

On systemic examination

Respiratory system:--B/L vesicular breathing, B/L airway equal air entry, no wheeze,

Cardiovascular system: - S1 S2 normal, no murmurs

CNS: - conscious and oriented with no sensorimotor deficit,

Per Abdomen examination

ON INSPECTION: - abdomen not distended, umbilicus central and inverted, no visible venous prominences, no visible pulsations.

ON PALPATION: - soft, non-tender, spleen palpable 2 cm below LCM, No guarding, no rigidity.

ON PERCUSSION: - Tympanic note with no fluid present

ON AUSCULTATION: - Normal bowel sounds present

**SYSTEMATIC REVIEW:**

Patient was admitted with above mentioned complaints. Her examination findings were as mentioned above. Her initial lab data and latest lab data is included at last of summary. Serum IgG4 – 0.65, AMA- M2 +++(strong positive). She received 3 Cryo and 3 RDPC, and underwent an UGI Endoscopy and Colonoscopy on 01/10/2024, which revealed a coagulopathy related ooze with normal Mucosa at ileum, caecum and colon. External haemorrhoids were seen. Rectal biopsy was not taken i/v/o coagulopathy. All vitals, temperature, RBS and other necessary parameters were checked regularly and managed appropriately. She is discharged in hemodynamically stable condition with the advice to follow up in OPD.

**PLAN / ADVICE AT DISCHARGE (Including duration of medication if any):**

2100 KCAL/DAY + 90 GRAMS PROTEIN/DAY

TAB TAXIM-O 200MG BD	1-0-1 X 5 DAYS

TAB ZINC 50 MG PO BD

TAB ME-12 1 TAB PO OD

CAP BETADEK 1 CAP PO OD

TAB URSOCOL 450 MG PO BD

CAP HENZOVIT 1 TAB PO OD

HEPSURE SACHET PO BD

**Next follow up –** Hepatology OPD / Virtual OPD after two weeks with CBC / KFT / LFT / INR

IN CASE OF DECREASE IN URINE OUTPUT, ALTERED SENSORIUM, BLEEDING, FEVER, NEW ONSET COUGH AND REVIEW IN ILBS EMERGENCY ON SOS BASIS.

### Summary 36

PHTN - Bleeder, Non-Bandable Esophageal Varices, Mild PHG, Duodenal Ulcer – 14/01/2025

CLD- NASH related

Decompensated with Ascites / Jaundice / HE

Rapid Filling Left Hydrothorax, Prior SBE

CTP-9	CHILD-B	MELD Na-19

**CURRENT ISSUES:-**

- Malena s/p UGIE - Non-Bandable Esophageal Varices, Mild PHG, Duodenal Ulcer – 14/01/2025
- Refilling Left Hydrothorax, Partially Treated SBE, s/p ICD inserted on 16/01/2025, removed on 22/01/2025- C/S: Stenotrophomonas maltophilia
- PET-CT suggestive of Pulmonary TB – decision regarding ATT after review in Pulmonology OPD

**PRESENTING COMPLAINTS:**

- Malena x 2 days
- Breathlessness x 5 days

**INDICATION FOR ADMISSION:**

Evaluation & management

**HISTORY:**

Mrs Patient AJ is a 66 year old lady with comorbidity of HTN who had her index presentation in 2014 when she underwent evaluation for cholelithiasis and incidentally found to have CLD. She was admitted in ILBS with c/o- shortness of breath and on evaluation she was found to have hepatic hydrothorax and underwent pleural tapping 4 times. USG showed CLD with PHTN. Ascitic fluid analysis showed high SAAG, low protein with no SBP. In view of Left Hydrothorax, ICD was inserted and Pleural fluid analysis showed high SPAG, low protein with SBE and negative for malignant cytology. She was discharged in satisfactory condition, and was on OPD follow up. She had one further decompensation in the form of HE, which was conservatively managed. She currently presented with complaints of malena, and was ad mitted for the same. She has now come to ILBS for further evaluation and management. She has a history of fall on 11/01/2025 for which she took analgesics. There is H/O-Hysterectomy in 2006 and cholecystectomy in 2014. There is no h/o- jaundice, vomiting, cough, abdominal pain, altered bowel habits, hematemesis, burning micturition, altered sensorium or decreased urine output. There is no h/o any intoxications, indigenous medications or IV drug abuse prior to onset of the disease. There is no h/o-CAD/TB/COPD/Thyroid disorders.

**EXAMINATION:**

Pt. was conscious, oriented, afebrile.

BP- 136/70 mm Hg	Pulse 80/min	RR 20/min

Pallor-, Icterus-, Cyanosis-, Clubbing-, Pedal edema-, LNP-, JVP normal

CVS S1 S2 normal, no murmurs

Chest - B/L rhonchi +, occasional crepts, left sided decreased air entry

CNS Patient conscious, oriented, no sensorimotor deficit.

P/A ON INSPECTION - distended ,umbilicus central and inverted, no visible venous prominences, no visible pulsations.

ON PALPATION soft, non tender, liver not palpable and Spleen palpable 2cm BCM. No guarding, no rigidity, no rebound tenderness.

ON PERCUSSION dull note, free fluid +

ON AUSCULTATION normal bowel sounds present, no bruits.

**SYSTEMATIC REVIEW:**

Patient was admitted with above mentioned complaints. Her examination findings were as mentioned above. Lab data is attached at the end of the summary. Blood and Urine Cultures revealed no growth, and Urine Routine was unremarkable. S. PCT – 0.21. She was admitted in HDU in view of dyspnea. RSV Panel – Negative. She was started on Oxygen support, IV antibiotics other supportive management. UGIE was done, which revealed Non-Bandable Esophageal Varices, Mild PHG, Duodenal Ulcer – 14/01/2025. She was closely monitored in HDU. Regular Pulmonology review done. ICD was inserted on 16/01/2025 in view of left hydrothorax and daily 1-1.5 litres drained. Pleural fluid analysis was done which revealed a straw coloured fluid with WBC - 282, N – 45% and L – 54%, Protein: 0.57, Glu: 136, SPAG: 1.72, ADA: 1.1 and Gene Xpert: negative, Gram Stain negative. HRCT Chest was done on 16/01/2025 reported

- Left moderate pleural effusion seen with underlying lung collapse and mild mediastinal shift to right side.
- Patchy parenchymal GGOs are seen in right apex as well as right mid zones –paracardiac location involving pneumonitis consolidation – likely infective.

NCCT Head - Mild cerebral and cerebellar atrophy. No significant change. She recovered clinically, and was shifted to the ward on 17/01/2025. Oxygen support was tapered and she underwent a PET-CT on 20/01/2025 –

- Features suggestive of chronic liver disease with portal hypertension, ascites and splenomegaly as described. No focal metabolically active or arterial hyperenhancing lesion noted in the liver parenchyma.
- Left sided pleural effusion.
- Mild Metabolically active tree in bud nodules with surrounding ground glass opacities in right lung upper lobe- infective.
- Metabolically active enhancing lesion in right parotid gland - likely Warthins tumour.

PET-CT suggestive of Pulmonary TB – decision regarding ATT after review in Pulmonology OPD. The nature of the disease and long term prognosis was explained in great detail to the patient and the patient relatives. She recovered symptomatically following the management. Proper diet, purging and mobilisation was followed throughout the course of admission. All vitals, temperature, RBS and other necessary parameters were checked regularly and managed appropriately. She is being discharged in stable hemodynamic state with advice of follow up in OPD.

**PLAN / ADVICE AT DISCHARGE (Including duration of medication if any):**

DIET AS ADVICED BY DIETICIAN

TAB LEVOFLOX 500 MG PO OD X 5 DAYS

T. RIFAGUT 550 MG PO BD	1-0-1 (START ON 6TH DAY)

TAB UDCA 450 MG PO BD

HEPSURE SACHET PO BD

LAXOPEG SACHET BD. 1-0-1 (ENSURE 2-3 BOWEL MOTIONS/DAY)

SYP LACTIHEP 30ML PO HS  0-0-1 (ENSURE 2-3 BOWEL MOTIONS/DAY)

SYP ZINCONIA 10 ML PO BD

STEAM INHALATION

LEVOLIN NEBULIZATION BD 1-0-1 X 3 DAYS THEN SOS

REGULAR SPIROMETRY

REVIEW IN HEPATOLOGY AND PULMONOLOGY OPD WITH CBC/LFT/KFT/INR/CXR-PA REPORTS AFTER 1 WEEK.

IN CASE OF DECREASE IN URINE OUTPUT, ALTERED SENSORIUM, BLEEDING, FEVER, NEW ONSET COUGH AND REVIEW IN ILBS EMERGENCY ON SOS BASIS.

### Summary 37

Portal Hypertension (Prior Bleeder, Small High Risk Esophageal Varices, EVL done, Mild PHG, Partially obturated GOV2 – Glue injected – 21/04/2025)

EHBO (K83.1) with Hilar Mass

- Granulomatous etiology (Biopsy Proven; Culture positive for MTB)
- S/P modified ATT (for extra pulmonary PB - August 2021 🡪 July 2022, Restarted in February 2023 🡪 July 2023)
- SV and SMV Thrombosis; Portal Cavernoma
- on Anticoagulation (Dabigatran) from August 2021 🡪 September 2022

TJLB on 19/01/2023 showed inflow vascular pathology

HVPG - 6mmHg

Prior Bleeder - AVB – 2 episodes in 2023 (s/p EVL) and 2024 (s/p Glue inj)

Planned for Splenectomy > PARTO/PSAE

**COMORBIDITIES**- Symptomatic GSD

**PRESENTING COMPLAINTS:**

Generalized Weakness x 5 days

**INDICATION FOR ADMISSION:**

Further evaluation & management of symptoms

**HISTORY:**

Mr. Patient AK is a 50 years old male, non-ethanolic, non-smoker with no known prior comorbidities. He had his index presentation in September 2021 in the form of pain abdomen and jaundice for which he was evaluated. MRI revealed a perihilar mass with extension to suprapancreatic region. Biopsy of the lesion revelaed granulomatous etiology and culture grew Mycobacterium. He was started on modified ATT and was managed conservatively. In view of SMV thrombosis, he was started on Dabigatran. However, he had 2 episodes of bleed in 2023 and 2024. Currently, he has presented with above mentioned complaints for possibility of PSAE / PARTO. There is no h/o any intoxications, major surgeries, blood transfusions or IV drug abuse prior to onset of the disease. There is no h/o T2DM/HTN/CAD/Thyroid disorders.

**EXAMINATION:**

Pt. was conscious, oriented, afebrile

BP- 124/78 mm Hg	Pulse -68/min	RR -18/min

Pallor-, Icterus++, Cyanosis-, Clubbing-, Pedal edema, LNP-, JVP normal

CVS -S1 S2 normal, no murmurs

Chest- B/L air entry present, no added sounds

CNS -conscious, oriented to time and place

P/A ON INSPECTION -not distended, umbilicus central and inverted, no visible venous prominences, no visible pulsations.

ON PALPATION -soft, non tender, liver and Spleen- not palpable. No guarding, no rigidity, no rebound tenderness.

ON PERCUSSION -tympanic note, no free fluid

ON AUSCULTATION- normal bowel sounds present, no bruits.

**SYSTEMATIC REVIEW:**

Patient was admitted with the above mentioned complaints. His examination findings were as mentioned above. Case was discussed with IR. CECT Abdomen – provisionally portal cavernoma with splenic vein thrombosis and collaterals; gastric varices; GLR shunt of 11 mm. Final report awaited. UGIE was done - Small High Risk Esophageal Varices, EVL done, Mild PHG, Partially obturated GOV2 – Glue injected – 21/04/2025. Case was initially planned for PSAE and PARTO; however, in view of non-feasibility of the same, he was planned for splenectomy. Case was discussed with HPB surgery for splenectomy. Patient was treated with IV antibiotics, nutritional support and other supportive medication and treatment. The nature of the disease and long term prognosis was explained in great detail to the patient and the patient relatives. Patient remained afebrile with no significant complains in the ward. Proper diet, purging and mobilisation was followed throughout the course of admission. All vitals, temperature, RBS and other necessary parameters were checked regularly and managed appropriately. He recovered symptomatically following the management and. He is discharged in haemodynamically stable with the advice to follow up in OPD.

**PLAN / ADVICE AT DISCHARGE (INCLUDING DURATION OF MEDICATION IF ANY):**

2100 KCAL/DAY + 90 GRAMS PROTEIN/DAY, NORMAL DIET

TAB TAXIM-O 200 MG PO BD FOR 5 DAYS

TAB CARDIVAS 6.25 MG PO BD (DO NOT GIVE IF HR<60, BP < 90 MMHG)

TAB PANTOCID 40 MG PO OD BBF X 14 DAYS

SYP SUCRAL 10 ML PO QID X 14 DAYS

TAB ULTRACET PO SOS

TAB MYORIL 4 MG PO SOS FOR BACK PAIN

SYP LACTIHEP 15ML PO SOS (ENSURE 2-3 BOWEL MOTIONS/DAY)

REVIEW IN HEPATOLOGY OPD WITH CBC/LFT/KFT/INR REPORTS AFTER 4 WEEKS.

MRI LUMBOSACRAL SPINE PLANNED FOR BACK PAIN

**UGIE +/- ENDOTHERAPY AFTER 3 WEEKS**

**FOLLOW UP IN HPB – SX OPD FOR SPLENECTOMY**

IN CASE OF DECREASE IN URINE OUTPUT, ALTERED SENSORIUM, BLEEDING, FEVER, NEW ONSET COUGH AND REVIEW IN ILBS EMERGENCY ON SOS BASIS.

### Summary 38

Diagnosis: DIAGNOSIS: Hyper Acute Liver Failure- HBV Related [HBsAg- Reactive, IgM HB core- reactive] (ICD10 - K 72.01) -	Jaundice à HE x 4 days	-	KCH - meeting single criteria	CT liver volumetry 893.66cm2 MELD- 37	SOFA- 14 Comorbidity : nil Current Issues -	1)	Encephalopathy grade 4 (S. NH3 1121); s/p intubated; s/p - CRRT Started on 09/09/2025 2)	Refractory Cerebral edema with B/L Sluggish reactive à non reactive; s/p Mannitol 3)	Coagulopathy with oral bleed (PT/INR- unrecordable); s/p 1 session of PLEX (09/09/2025)	PRESENTING COMPLAINTS	Fever x 5 days Vomiting x 3 episode, 5 days back	Loose stool x 3 episode, 5 days back	Jaundice x 5 days Altered sensorium x 1 day | Admitted For: Evaluation &amp; management	HISTORY: Mr. Patient AL, 41 year old male, with no prior co-morbidities was apparently well 5 day back when he developed fever for 1 days, associated with loose stools and vomiting; and yellowish discoloration of sclera and urine, insidious onset, gradually progressive, not associated with pruritus or clay coloured stool. PCM intake + (dose records NA). Later on he developed worsening of symptom he was admitted in outside and diagnosed as a case of ALF HBV related. Now he presented to ILBS with above mentioned complaints. He was admitted for further evaluation and management. There is no h/o past major surgeries, or IV drug abuse prior to onset of the disease. There is no h/o T2DM/HTN/CAD/ COPD /Thyroid disorder. EXAMINATION: At the time of admission Pt. altered sensorium BP: - 170/80mm Hg, Pulse: 133 /min, RR: - 22/min, Spo2 99% on RA. Pallor -, Icterus +, Cyanosis-, Clubbing-, Pedal edema -, LNP-, JVP Normal	On Systemic Examination	Respiratory System: - B/L vesicular breathing, B/L airway equal air entry, no wheeze, no crepts	Cardiovascular system: - S1 S2 normal, no murmurs	CNS: - altered sensorium	Per Abdomen examination	ON INSPECTION: - Distended abdomen, umbilicus central and everted, skin over the abdomen is stretched with no visible venous prominences, no visible pulsations, . ON PALPATION: - Soft, non tender, liver palpable 2 cm below coastal cartilage, Spleen non palpable. No guarding, no rigidity, no rebound tenderness. ON PERCUSSION: - Dull note, Free fluid is present	ON AUSCULTATION: - Bowel sounds present	SYSTEMATIC REVIEW: Patient was admitted with above mentioned complaints. His examination findings were as mentioned above. His initial lab data revealed Hb/TLC/PLT- 12.5/23.49/133. PT/- &gt;120, BU/S.Creat- 18.8/1.06 with serum Na+/K+ -140.2/4.64. LFT showed S.Bil (total/direct/indirect: 9.37/3.2/6.17, AST/ALT- 3426/3447, SAP/GGTP- 198/44, T.Prt /Alb- 5.59/2.91. Ammonia - 1121. Patient was received in ER. CCM Team review was done and advice followed Immediate IV fluid resuscitation was started. Patient was intubated and put on mechanical ventilator in view of HE grade IV on 08/09/25. Patient was admitted in LC ICU. Patient was started on aggressive management with IV antibiotics, IV anti-fungals, IV fluids, IV albumin and other supportive measures. HD catheter insertion done. After iv fluid resuscitation, he was initiated on CRRT. Urine examination (sugar- nil, alb- nil, rbc- 30-35, leu- 2-3. Sepsis screen is (Blood c/s and urine c/s awaited ).HBsAg reactive, IgM HB core reactive, HBV DNA- not detected/Anti HCV/Anti HEV/HIV /Anti HAV - non Reactive. Mini BAL sample was sent, g/s- no bacteria, KOH- ND, Biofire not detected. NCCT Head normal study. NCCT Abdo- Chronic liver disease with findings suggestive of portal hypertension (dilated SP-axis, splenomegaly and abdominal collaterals) and	overlying diffuse heterogeneous attenuation with hypodensity of liver parenchyma suggestive of ?ACLF. HRCT chest - Impression :- Bilateral lower lobe consolidations.. 2D-ECHO s/o Normal LV size and normal systolic function. Global EF approx- 60%. RV size &amp;contractility appear normal. No pericardial effusion. Colour flow &amp; doppler studies could not be done. Valves could not be assessed properly. IVC-18mm. Pulmonary clearance taken for LT advice followed .neurology clearance was taken and advice followed. cardiology clearance taken in view of LT advice followed. Urgent HPBSx consult for LDLT sent in view of coagulopathy and refractory cerebral edema. He underwent 1 session of high volume PLEX on 09/09/2025. Relatives have been explained in great detail the need for LT as definitive management of disease. LT work up sent urgently and patient was optimized. On 09/09/2025, he developed ICP surges with sluggish reactive pupils, started on mannitol . Repeated HPB Sx opinion sent, patient is being taken up for LDLT. Attendants counseled regarding grave prognosis, nature of disease and need of LT as definitive t/t option. Advised intraoperative and post operative CRRT from Hepatology side. Patient is being discharged from Hepatology side for LDLT to get admitted in surgery side. Current Vitals- Patient sedated and paralyzed on INJ FENTANYL/PROPOFOL/ATRA @ 3ML/HR SpO2-100% ON PSIMV MODE, Fio2 - 40%, Rate - 14, PEEP - 10 BP- 164/88(112), HR- 84/Min	UO- 50 Ml/Hr ABG: PH - 7.54, PaO2 - 197, PaCO2 - 38, HCO3 30.2, Lact 2.7	CURRENT TREATMENT: ON CRRT	-INJ POLYMYXIN B 7.5 LAC MU IV BD -INJ MEROPENAM 1 GM IV TDS -INJ AMPHO B 250 MG IV OD -INJ TARGOCID 400MG IV OD	-INJ TRANEXA 1 GM IV TDS -TAB TAF 25 MG RT OD	-INJ LEVIPIL 750 MG IV BD -NEB COLISTIN TDS	-INJ NAC @2.1 IV	-IVF 25% DEXTROSE @ 10 ML/HR -IVF PLASMOLYTE 100 ML IV

### Summary 39

PHTN - Non-Bleeder, Small Low Risk Esophageal Varices, Mild PHG, Antral Erosions 01/03/2025

Cirrhosis Likely NASH **(ICD - K74.6)**

Decompensated Ascites / HE

LT Explained

CAP 181	LSM 75.0	SSM 83.6

**COMORBIDITIES**:

-          None

**CURRENT ISSUES:**

-	Hepatic Encephalopathy Grade III 🡪 Resolved

-     Grade III Ascites (High SAAG, Low Protein, No SBP)

-     AKI KDIGO II HRS (Creat 1.83 🡪 1.17)

**PRESENTING COMPLAINTS:**

- Abdominal distension x 3 months
- Altered Sensorium x 2 days

**INDICATION FOR ADMISSION:**

Evaluation and the management of the symptoms

**HISTORY:**

Mr Patient AM is a 56 years old gentleman, who had his index presentation in August 2024 at an outside centre when on evaluation of generalized weakness, he was diagnosed to have cirrhosis. Over the past three months, he developed a new onset abdominal distension and pedal edema. LVP was done twice and he was initiated on diuretics, however ascites persisted. He was recently admitted in February 2025, in ILBS, with refilling ascites. UGIE was done and revealed Small Low Risk Esophageal Varices, Mild PHG, Antral Erosions on 01/03/2025. . LT referral done. Now he has presented to ILBS with complaints of altered sensorium and got admitted for further evaluation and management. There is no h/o fever, jaundice, vomiting, cough, abdominal pain, altered bowel habits, hematemesis, and malena, burning micturition or decreased urine output. There is no h/o any intoxications or IV drug abuse prior to onset of the disease.

**EXAMINATION**

Pt. was conscious, disoriented, Grade III HE

BP: - 132/78 mm Hg, Pulse: - 111/min, RR: - 14/min and afebrile.

Pallor+, Icterus-, Cyanosis-, Clubbing-, Pedal edema (pitting type) - , LNP-, JVP normal

On systemic examination

Respiratory system:--B/L vesicular breathing, B/L airway equal air entry, no wheeze,

Cardiovascular system: - S1 S2 normal, no murmurs

CNS: - conscious, disoriented, Grade III HE

Per Abdomen examination

ON INSPECTION: - distended abdomen, umbilicus central and inverted, skin over the abdomen is stretched with no visible venous prominences, no visible pulsations.

ON PALPATION: - soft, non-tender, no organomegaly. No guarding, no rigidity, no rebound tenderness.

ON PERCUSSION: - dull note, free fluid present.

ON AUSCULTATION normal bowel sounds present

**SYSTEMATIC REVIEW:**

Patient was admitted with the above mentioned complaints. His examination findings were as mentioned above. Blood and Urine Cultures revealed no growth, and Urine Routine was unremarkable. S. PCT – 0.79. Plasma Ammonia – 185.6. He was shifted to the ICU. Anti-coma measures initiated. He was started on IV antibiotics, and supportive medications until he symptomatically improved. Sensorium improved and he was symptomatically better. He was shifted to the ward. The nature of the disease and long term prognosis was explained in great detail to the patient and the patient relatives and were also explained regarding the need of liver transplant in view of Decompensated Liver Disease. Ascitic fluid tap was done which revealed a straw coloured fluid with WBC - 98, N – 20% and L – 79%, Protein: 1.63, Glu: 103, Albumin: 0.64, SAAG: 2.22, ADA: 5.8 and Gene Xpert: negative, Gram Stain negative. Cultures were negative. CXR revealed right sided minimal pleural effusion, however, patient remained asymptomatic. He is now planned for nutritional optimization and improvement in performance status. Proper diet and mobilisation was followed throughout the course of admission. All vitals, temperature, RBS and other necessary parameters were checked regularly and managed appropriately. He recovered symptomatically following the management and is discharged in hemodynamically stable condition with following advice to follow in OPD.

**PLAN / ADVICE AT DISCHARGE (Including duration of medication if any):**

2100 KCAL/DAY + 90 GRAMS PROTEIN/DAY, LOW SALT (<2GRAMS/DAY)

**WEIGHT REDUCTION WITH DAILY 30 MIN EXERCISE**

TAB TAXIM-O 200MG BD	1-0-1 X 5 DAYS

T. RIFAGUT 550 MG PO BD	1-0-1 (START ON 6TH DAY)

TAB MIDODRINE 2.5 MG PO TDS

TAB NUHEME 1 TAB PO OD

CAP HENZOVIT 1 TAB PO OD

TAB ME-12 1 TAB PO OD

TAB ZINC 50 MG PO BD

HEPSURE SACHET PO BD

SYP LACTIHEP 30ML PO HS  0-0-1 (ENSURE 2-3 BOWEL MOTIONS/DAY)

LAXOPEG SACHET BD. 1-0-1 (ENSURE 2-3 BOWEL MOTIONS/DAY)

HEPAMERZ SACHET 1 TDS 1-1-1

INJ. ALBUMIN 20% 100 ML IV ONCE A WEEK UNDER MEDICAL SUPERVISION OVER 4-6 HOURS

LVP SOS UNDER ALBUMIN COVER

VIRTUAL OPD AFTER 1 WEEK WITH CBC / LFT / KFT / INR REPORTS

REVIEW IN HEPATOLOGY OPD WITH CBC / LFT / KFT REPORTS AFTER 2 WEEKS

PLAN: LT WORKUP ON OPD BASIS

IN CASE OF DECREASE IN URINE OUTPUT, ALTERED SENSORIUM, BLEEDING.

### Summary 40

EHBO

Etiology - Ca Gall Bladder with Metastasis **(ICD10 - C23)**

S/P- ERCP + EPT + SEMS – SEMS placed, PD stent placed - 25/10/2024

CEA – 35.095, CA19.9 – 216, PIVKA II – 3463.44, AFP - 7

**Co Morbidities** – Hyperthyroid x 5 years

**CURRENT ISSUES:**

- EHBO - Ca Gall Bladder with Metastasis S/P- ERCP + EPT + SEMS - 25/10/2024
- Osteomyelitis – Left Ankle Wound with Tarsal Bone Involvement – Wound Swab growing Staphylococcus aureus sensitive to Teicoplanin

**PRESENTING COMPLAINTS:**

Jaundice x 30 days

Abdominal Pain x 30 days

**INDICATION FOR ADMISSION:**

Evaluation and management of symptoms

**HISTORY:**

Mrs. Patient AN is a 51 year old lady with known co morbidities as above. She had her index presentation at an outside centre 1 month ago when she developed Jaundice and abdominal pain. She was evaluated outside and found to have a lesion ?CA GB. She has presented to ILBS for the same. She first noticed a yellowish discolouration of eyes and urine since 30 days, which was insidious in onset and progressive. It was associated with itching or clay-coloured stools. She also complaints of a dull aching abdominal pain. The pain was insidious in onset, localised to upper abdomen, non-colicky, with no radiation or migration. There is no h/o, cough, altered bowel habits, hematemesis, and malena, burning micturition, altered sensorium or decreased urine output. There is no h/o any intoxications, indigenous medications, other major surgeries, blood transfusions or IV drug abuse prior to onset of the disease. There is no h/o DM/HTN/CAD/TB/COPD.

**EXAMINATION**

Pt. was conscious, oriented to time place and person.

BP: - 106/70 mm Hg, Pulse: - 86/min, RR: - 14/min and afebrile.

Pallor+, Icterus+, Cyanosis-, Clubbing-, Pedal edema (pitting type) - , LNP-, JVP normal

On systemic examination

Respiratory system:--B/L vesicular breathing, B/L airway equal air entry, no wheeze, no crepts

Cardiovascular system: - S1 S2 normal, no murmurs

CNS: - conscious and oriented with no sensorimotor deficit,

Per Abdomen examination

ON INSPECTION: - Non distended abdomen, umbilicus central and inverted, skin over the abdomen is stretched with no visible venous prominences, no visible pulsations.

ON PALPATION: - soft, non-tender, GB palpable. No guarding, no rigidity, no rebound tenderness.

ON PERCUSSION: - Tympanic note with no fluid present

ON AUSCULTATION -normal bowel sounds present,

**SYSTEMIC REVIEW**

Patient was admitted with above mentioned complaints. Her examination findings were as mentioned above. Blood cultures revealed no growth, and Urine Routine was unremarkable. She was initiated on IV antibiotics, nutritional therapy and other supportive medications. MRCP was done (final report awaited). A PET-CT was done on 23/10/20241.

- Metabolically active heterogeneously enhancing lesion in the fundus of gallbladder with extension to adjacent liver parenchyma- probably neoplastic in aetiology (primary carcinoma gallbladder).
- Metabolically active hypodense lesions in both lobes of liver- metastatic.
- Metabolically active heterogeneously enhancing left supraclavicular, mediastinal and upper abdominal lymph nodes- metastatic.
- Metabolically active left heterogeneously enhancing left external iliac lymph nodes- suspicious for metastases.
- Abrupt cut-off of the distal CBD with upstream dilatation of proximal CBD, CHD, cystic duct and bilobar IHBRD. No focal metabolic activity or enhancing thickening noted in the distal CBD- ? Partial stricture- advise MRCP correlation for further evaluation.
- Patchy areas of metabolically active short segment wall thickenings with the proximal dilatation in the entire length of colon- a colonoscopy with biopsy may be advised to rule out Crohn’s disease.
- Non-metabolically active cystic lesion in the proximal body of pancreas- pancreatic cyst.

She underwent ERCP + EPT + SEMS on (25/10/2024) – SEMS placed, PD stent placed. Wound was noted on lower limb with extension to bone and MRI lower limb was done. Orthopaedic consult was done and advice followed. She tolerated the procedure well and post procedure care was provided. Medical Oncology opinion was sought and advice followed. Proper diet and mobilization was followed throughout the course of admission. All vitals, temperature, RBS and other necessary parameters were checked regularly and managed appropriately. The nature of the disease and long term prognosis was explained in great detail to the patient. Now she has improved symptomatically and is being discharged in hemodynamically stable condition with the advice to follow up in OPD.

**ADVICE** **ON DISCHARGE (Duration for medications if any):**

2000 KCAL/DAY + 90 GRAMS PROTEIN/DAY, NORMAL DIET

TAB FAROPENAM 200 MG PO BD FOR 7 DAYS

INJ TEICOPLANIN 400 MG OD FOR 7 DAYS

TAB URSOCOL SR 450 MG PO BD

CAP BECOSULE PO OD
SYP LACTIHEP 30ML PO SOS/HS  0-0-1

TAB ULTRACET 1 TAB PO SOS FOR PAIN

TAB CALPOL SOS FOR FEVER

TAB NEOMERCAZOLE 5 MG PO TDS TO CONTINUE

FOLLOW UP IN HEPATOLOGY / MEDICAL ONCOLOGY OPD WITH CBC, LFT, KFT, MRCP, INR REPORT IN 2 WEEKS

FOLLOW UP AT ORTHOPAEDIC CENTRE FOR MANAGEMENT OF LOWER LIMB OSTEOMYELITIC WOUND

IN CASE OF DECREASE IN URINE OUTPUT, ALTERED SENSORIUM, BLEEDING, FEVER, NEW ONSET COUGH AND REVIEW IN ILBS EMERGENCY ON SOS BASIS.

### Summary 41

PHTN (Bleeder, Residual High Risk Esophageal Varices with clean based ulcers – 05/04/2025)

Cirrhosis – Likely NASH

Decompensated - AVB

Active Post EVL Ulcer Bleed – s/p DE stent insertion – 30/03/2025 - removed on 05/04/2025

S/P TIPSS with Variceal Embolization – 04/04/2025

CTP - 8	CHILD – B	MELD Na – 7

**COMORBIDITIES:** T2DM – Newly Diagnosed

**CURRENT ISSUES:**

- Hematemesis s/p DE stent insertion – 30/03/2025 - removed on 05/04/2025
- S/P TIPSS with Variceal Embolization – 04/04/2025

**PRESENTING COMPLAINTS:**

Hematemesis x 3 episodes – 12 hours ago

**INDICATION FOR ADMISSION:**

Evaluation and the management of the symptoms

**HISTORY:**

Mr Patient AO is a 55 year old gentleman with no known comorbidities or prior surgeries. His index presentation was March 2025 with a history of hematemesis and was diagnosed to have a liver disease EVL was done – glue injected on 10/03 and EVL done on 13/03. Currently, he again had episodes, and has come to ILBS for further management. There is no h/o any intoxications, indigenous medications, major surgeries, blood transfusions or IV drug abuse prior to onset of the disease. There is no h/o CAD/TB/COPD/Thyroid disorders.

**EXAMINATION**

Pt. was conscious, oriented to time place and person.

BP: - 70/30 mm Hg, Pulse: - 97/min, RR: - 14/min and afebrile.

Pallor +, Icterus +, Cyanosis-, Clubbing-, Pedal edema + , LNP-, JVP normal

On systemic examination

Respiratory system:-- B/L vesicular breathing, B/L airway equal air entry, no wheeze, Cardiovascular system: - S1 S2 normal, no murmurs

CNS: - conscious and oriented with no sensorimotor deficit,

Per Abdomen examination

ON INSPECTION: - non-distended abdomen, umbilicus central and inverted, no visible pulsations.

ON PALPATION: - soft, non-tender, Liver palpable 2 cm below RCM, No guarding, no rigidity, no rebound tenderness.

ON PERCUSSION: - tympanic note, free fluid present.

ON AUSCULTATION normal bowel sounds present,

**SYSTEMATIC REVIEW:**

Patient was admitted with above mentioned complaints. His examination findings were as mentioned above. His initial lab data and latest lab data is included at last of summary. Bleeder protocol initiated, and he was started on aggressive management with RT lavage, fluid resuscitation with IV fluids, blood transfusion, iv antibiotics, and other supportive measures. He was started on IV Terlipressin and blood products in the form of PRBC were transfused to correct acute blood loss. Immediate UGI endoscopy was done to investigate the bleed, which showed Large High Risk Esophageal Varices with post EVL scarring; GOV-1 and small GOV-2 seen, Glue injected. He was admitted in ICU in view of shock. Regular CCM consults done. In view of worsening respiratory distress with shock, he was intubated and initiated on MV on 30/03/2025. Relook UGIE was done in view of persistent shock - revealed active post EVL ulcer bleed. Glue was injected in GOV 1 and Danis Ella stent placed on 30/03/2025. Shock improved and he was continued on iv fluids, and inotropes. He was extubated on 31/03/2025. In view of recurrent bleeds, patient and his relatives have been explained about TIPSS And the issues with TIPSS with regards to risk of encephalopathy, and informed consent obtained. TIPSS was done by IR team on 03/04/2025 under all aseptic precautions. Post TIPS there was no episode of HE. Post procedural USG screening showed patent TIPS stent. He was stabilized and shifted back to the ward. Danis Ella stent was removed on 05/03/2025. Proper diet and mobilisation was followed throughout the course of admission. All vitals , temperature, RBS and other necessary parameters were checked regularly and managed appropriately. He recovered symptomatically following the management and is discharged in hemodynamically stable condition with following advice to follow in OPD.

**PLAN / ADVICE AT DISCHARGE (Including duration of medication if any):**

2100 KCAL/DAY + 90 GRAMS PROTEIN/DAY, LOW SALT (<2GRAMS/DAY)

WEIGHT REDUCTION WITH DAILY 30 MIN EXERCISE

STRICTLY AVOID CONSTIPATION

**ENSURE 2-3 BOWEL MOTIONS/DAY**

TAB FAROPENAM 200 MG PO BD FOR 5 DAYS

TAB FLUCOS 200 MG PO OD FOR 5 DAYS

TAB PANTOCID 40 MG PO OD BBF X 14 DAYS

SYP SUCRAL 10 ML PO QID X 14 DAYS

TAB LASILACTONE (20/50) **1/2 TAB** PO OD (MONITOR KFT WEEKLY)

HEPSURE SACHET PO BD

TAB UDCA 450 MG PO BD

TAB THIOTRESS 500 MG PO BD X 14 DAYS

SYP LACTIHEP 30ML PO HS  0-0-1 (ENSURE 2-3 BOWEL MOTIONS/DAY)

LAXOPEG SACHET BD. 1-0-1 (ENSURE 2-3 BOWEL MOTIONS/DAY)

REVIEW IN HEPATOLOGY OPD WITH CBC/LFT/KFT/INR REPORTS AFTER 4 WEEK.

PLAN UGIE AND SCREENING AFTER 4 WEEKS

IN CASE OF DECREASE IN URINE OUTPUT, ALTERED SENSORIUM, BLEEDING, FEVER, NEW ONSET COUGH AND REVIEW IN ILBS EMERGENCY ON SOS BASIS.

### Summary 42

Extrahepatic biliary obstruction: Type 3b block with cholangitis

Etiology- Hilar cholangiocarcinoma with liver metastasis (Segment VI)

- S/P ERCP + Stenting (Outside: 13/11/2024)
- S/P CT: FOLFOX regimen (December 2024🡪 March 2025)
- S/P ERCP + Stricturoplasty + Stent exchange (14/04/2025)
- Spontaneous stent displacement (04/05/2025)
- S/P USG guided PTBD insertion LAHD (05/05/2025)
- S/P ERCP + RPD stenting + PTBD stent exchange (15/05/2025)
- PS-1, CA19.9-32.1, CEA-6.81, AFP-2.0

Portal hypertension- Non bleeder, Grade II esophageal varices, Mild PHG (15/05/2025)

Secondary biliary cirrhosis- Likely metabolic + Infiltration related

Decompensated- Ascites (High SAAG, low protein, SBP+🡪 improving)

MELD Na-16, CTP-8, Child-B

Co morbidity- Type 2 DM, Hypothyroidism

ICD-K75.6/C22.1/E14.9/E03.0

**CHIEF COMPLAINS**

Pain abdomen and progressive abdominal distension from last 1 month

Worsening jaundice and passing clay colored stools from last 6 months

Generalized weakness and loss of appetite

Evaluation and the management of presenting symptoms

**HISTORY:**

Mr. Patient AP is a 58 years old male patient with prior co morbidities as mentioned above. Patient was apparently well 6-7 months back when he noticed pain abdomen insidious onset, localized to right upper abdomen, dull aching, non-colicky, with no radiation or migration. This was soon followed by jaundice painless, with yellowish discoloration of sclera and urine, insidious onset, gradually progressive, associated with pruritus and clay-colored stools. He was evaluated outside and was found to have EHBO. ERCP + Stenting was done for the same. Further evaluation showed hilar cholangiocarcinoma for which he received FOLFOX regimen between December 2024 and March 2025. Following this patient showed improvement but soon developed stent block. Further sessions of ERCP+ Stent exchange were taken followed by PTBD insertion for obstructive jaundice. However from last 1 month patient has developed abdominal distension which was insidious in onset, gradually progressive, painless, non-tender, not accompanied with pedal edema, and not associated with nausea, vomiting, colicky pain, constipation, obstipation, fever, diarrhea, passage of mucous or blood per rectum. With this clinical picture he has been admitted for further management. Patient was planned outside for SEMS placement in view of spontaneous displacement of stent. There is no h/o vomiting, cough, altered bowel habits, hematemesis, and malena, burning micturition, altered sensorium or decreased urine output. There is no h/o any intoxications, indigenous medications, major surgeries, blood transfusions or IV drug abuse prior to onset of the disease. There are no h/o HTN/CAD/TB/COPD/Renal disorders.

**EXAMINATION:**

Pt. was conscious, oriented to time place and person.

BP: 106/70 mmHg, HR- 86/min, RR- 14/min and afebrile. PTBD in situ

Pallor+, Icterus-, Cyanosis-, Clubbing-, Pedal edema (pitting type) - , LNP-, JVP normal

On systemic examination

Respiratory system:--B/L vesicular breathing, B/L airway equal air entry, no wheeze, no crepts

Cardiovascular system: - S1 S2 normal, no murmurs

CNS: - conscious and oriented with no sensorimotor deficit,

Per Abdomen examination

ON INSPECTION: Distended abdomen, umbilicus central and inverted, skin over the abdomen is stretched with no visible venous prominences, no visible pulsations.

ON PALPATION: soft, non-tender, liver non- palpable below right coastal margin. Spleen non- palpable below left coastal margin. No guarding, no rigidity, no rebound tenderness.

ON PERCUSSION: Dull note over all quadrants with free fluid present

ON AUSCULTATION: normal bowel sounds present,

**SYSTEMIC REVIEW:**

Patient was admitted with above mentioned complaints. His examination findings were as mentioned above. His initial lab data has been mentioned at the end of this case summary. Sepsis screening was done. Blood and urine cultures showed no growth. Urine routine showed normal study. Viral screening for hepatitis was non-reactive. Metabolic profile assessment and biomarker screening was done. Available reports have been provided to patient in digital format.

CECT abdomen was done for loco regional assessment. It showed Liver measures approx. 18-cm in craniocaudal span and shows cirrhotic architecture with lobulated outlines, widened interlobar fissure and relatively hypertrophied left and caudate lobes. Subcapsular areas of THAD are noted in the right lobe. Hypodense soft tissue mass is seen in segment-IV measuring approx 6 x 5.5 cm with associated capsular retraction likely infiltration of segment-IV division of LPV and associated involvement of the primary confluence. PTBD catheter are seen in both main ducts with minimal pneumobilia in segment-IV duct? blocked catheter. CBD stent is also seen in situ upto the confluence. Sub capsule 20-mm hypodense nodule with exophytic projection is seen in segment-IV / III –likely deposits. GB is not visualized? post operative? cause.  Prominent hypodense lymph nodes are seen in the porta. Main portal vein measures 17.7-mm, rest of PV divisions are patent and splenic vein measures 18.9-mm. Hepatic veins show normal contrast opacification. Multiple collaterals are seen in the paraesophageal, gastroepiploic, perigastric, splenic-hilar, mesentery and retroperitoneum regions. Thin recanalized para umbilical vein is seen. Dilated LGV (8.3-mm) giving efferent to perigastric and esophageal collaterals. Pancreas has normal outlines and parenchymal enhancement. No definite calcification is seen. MPD is not dilated. Spleen is enlarged in size (span 22-cm). Bilateral kidneys show normal size, outline and contrast enhancement. Adrenals are normal in size and shape. Abdominal aorta, IVC and other major abdominal vessels show normal contrast opacification except for atherocalcific changes. Few subcentimeter lymph nodes are seen in the upper abdomen, mesentery and retroperitoneum. Stomach, visualized small and large bowel loops are unremarkable except portal enteropathy and colopathy. Diffuse peritoneal thickening and enhancement –SBP. Gross free fluid is seen in the abdomen. UB, pelvis are unremarkable. Bilateral mild pleural effusion with basal atelectasis.

After confirmation of duct + stent block he underwent ERCP + Stenting followed by PTBD exchange. Patient tolerated the procedures well and post procedure period was uneventful. Following this in view of grade III ascites he underwent therapeutic paracentesis. Ascitic fluid analysis showed high SAAG low protein with SBP. IV antibiotic regimen was modified accordingly. Furthermore he has been planned for rendezvous stent insertion by ERCP guided technique.

Patient was treated with IV antibiotics, IV albumin, IV terlipressin, nutritional support and other supportive medication and treatment. Proper diet, purging and mobilization was followed throughout the course of admission. All vitals, temperature, RBS and other necessary parameters were checked regularly and managed appropriately

The advanced nature of the disease and long term prognosis was explained in great detail to the patient and the patient’s relatives. Family was also explained regarding the need of liver transplant as a definite option of management.

He has recovered symptomatically following the management and is being discharged (as LAMA) with advice to follow up on OPD basis.

**PLAN / ADVICE AT DISCHARGE (Including duration of medication if any):**

2100 Kcal/day + 80 grams protein/day, low salt (<2grams/day), Full fat, normal diabetic diet as advised

Daily 30 min exercise to be included into routine

Ensure 2-3 bowel movements/day and avoid constipation with good hydration

T. Faronam 200mg PO BD 1-0-1 for 7 days then stop

T. Rifagut 550mg PO BD 1-0-1 to be started from 8^th^ day onwards

T. TAB UDCA 450 MG PO BD 450mg PO BD 1-0-1

C. Viadek PO OD 0-1-0

T. Gabapin 100mg PO HS 0-0-1

Hepsure sachet PO BD 1-0-1

T. Lasilactone 20/50mg PO OD ½ -0-0 (monitor KFT every 5^th^ day)

Syp. Lactifibre 20ml PO HS 0-0-1 (Dose can be modified according to frequency of bowel movements)

Nutrix ultra whey Powder 2 scoops PO BD 1-0-1

To continue same medications for T2DM and hypothyroidism as prescribed outside

Regular blood sugar monitoring and hypoglycemic education done

Inj. Albumin 20% 100 ml IV once a week under medical supervision over 4-6 hours

Review in Hepatology OPD with CBC/LFT/KFT/INR reports after 2 weeks

PLAN: - Titrate the diuretic dose and Oncology follow up

ER SOS

### Summary 43

Portal HTN – Non Bleeder, Small High Risk Esophageal Varices, Mild PHG -12/02/2022   (HVPG – 19 mm Hg)

Cirrhosis of Liver – Burnt out NASH **(ICD10 - K75.8)**

Decompensated:- Ascites / Mild Right Effusion

CTP- 10, CHILD-C, MELD Na-14

**CO-MORBIDITIES**:

- T2DM
- HTN
- Mediastinal Lymphadenopathy (ATT taken in 2022)
- COVID-19 (3 times)-took oral steroids
- Asymptomatic GSD
- Severe HPS

**CURRENT ISSUES:-**

- Severe HPS
  - 2D Echo – 57% EF, Mild MR / TR, Annular Calcification of MV
  - Saline Contrast Echo shows severe pulmonary vascular shunting
  - On domiciliary O2 support
- ? LV Relaxation Failure – HFPrEF
- Advanced Liver Disease 🡪 **LT Explained**

**PRESENTING COMPLAINTS:**

- Shortness of Breath x 2 weeks

**INDICATION FOR ADMISSION:**

Evaluation and the management of presenting symptoms

**HISTORY:**

Mrs. Patient AQ is a 67 years old lady who had her index presentation 3.5 years back in the form of vomiting and decrease appetite on evaluation its diagnosed as cirrhosis (NASH related). Patient was on regular follow up. Biopsy done revealed burnt out NASH. She now presented with c/o shortness of breath over the past 2 weeks, associsated with a cough. Patient now has come to ILBS with above mentioned complains and for further evaluation and management. There is no h/o pain, dysuria, sweating, altered sensorium, burning micturition or decreased urine output. There is no h/o any intoxications, major surgeries, blood transfusions or IV drug abuse prior to onset of the disease. There are no h/o B.Asthma /CAD/Thyroid disorders.

**EXAMINATION:**

Pt. was conscious, oriented, afebrile

BP- 128/78 mm Hg     Pulse 70/min RR-18/min

Pallor -, Icterus+, Cyanosis-, Clubbing-, Pedal edema+, LNP-, JVP-normal

CVS -S1 S2 normal, no murmurs.

Chest- B/L equal air entry present, no added sounds.

CNS -conscious, oriented to time,place & person.

P/A examination:

ON INSPECTION-distended, umbilicus central and inverted, no visible venous prominences,

no visible pulsations.

ON PALPATION-Soft,non tender, liver non palpable and spleen non palpable. No guarding, no rigidity, no rebound tenderness.

ON PERCUSSIONdull note, free fluid present.

ON AUSCULTATION-Normal bowel sounds present, no bruits.

**SYSTEMATIC REVIEW:**

Patient was admitted with the above mentioned complaints. Her examination findings were as mentioned above. Respiratory Virus Panel – Negative. 2 D Echo - 57% EF, Mild MR / TR, Annular Calcification of MV. Saline Contrast Echo shows severe pulmonary vascular shunting. She was initiated on iv fluids, iv antibiotics and O2 support as required. IV Diuretics initiated. Pulmonology consults taken and advice followed. PFT with Diffusion Studies - Mild Restriction with Small Airway Limitation. Post bronchodilator - significant reversibility. Diffusion Study: Impaired Diffusion. FENo - Normal Study. CECT Whole Abdomen and HRCT done – provisionally, no SOL, no PVT, Distended GB with multiple calculi. Mild ascites seen, mild right pleural effusion. Final report awaited. Her Oxygen saturation maintained between 92-94%. The nature of the disease and long term prognosis was explained in great detail to the patient and the patient relatives and were also explained regarding the need of liver transplant in view of Decompensated Liver Disease with Severe HPS. LT referral done and risk of proceeding with LT explained by HPB team. Family needs more time for the decision at present. She recovered symptomatically following the management and is discharged with following advice to follow in OPD.

**PLAN / ADVICE AT DISCHARGE (Including duration of medication if any):**

2000 KCAL/DAY + 90 GRAMS PROTEIN/DAY, LOW SALT DIET

TO AVOID ANY ALTERNATIVE MEDICINE OR THERAPY

DAILY 30 MIN EXERCISE AND MUSCLE STRENGTHENING EXERCISE

INTERMITTENT OXYGEN AT HOME WITH TARGET spO2 >95%

TAB FAROPENEM 200 MG PO BD                  1-0-1 x 5 DAYS

TAB LEVOFLOX 500 MG PO OD                          1-0-0 EVERY ALTERNATE DAY

T. RIFAGUT 550 MG PO BD	1-0-1 (START ON 6TH DAY)

TAB EPTUS 25 MG PO OD (MONITOR KFT WEEKLY)

TAB LASILACTONE (20/50) ½ PO BD	½ - ½ - 0	(MONITOR KFT WEEKLY)

TAB TRENTAL 400 MG PO OD

HEPSURE SACHET PO BD

CAP HENZOVIT 1 TAB PO OD

NEBULIZATIONS LEVOLIN NEB 1 RESPULE BD

NEUBILZATIONS FORACORT 1 RESPULE BD

INCENTIVE SPIROMETRY AS ADVISED

ER SOS

### Summary 44

Portal HTN- Non Bleeder, Small High risk esophageal varices, mild PHG, ? Duodenal varix - S/P EVL (12/12/2024)

Cirrhosis- Metabolic

Decompensated- Ascites / AVB

MELD Na-13, CTP-8, Child-B

Co morbidity- T2DM | Obesity

ICD-K75.6/E14.9

**CURRENT ISSUES:**

- Malena S/P UGIE - Small High risk esophageal varices, mild PHG, ? Duodenal varix - S/P EVL (12/12/2024)
- Left Bells Palsy – on Dexona, Valacyclovir

**CHIEF COMPLAINS**

- Blackish tarry stools x 7 days
- Left sided ear pain x 15 days
- Left sided facial weakness x 15 days

**INDICATION FOR ADMISSION:**

Evaluation and the management of presenting symptoms

**HISTORY:**

Mrs Patient AR is a 58 years old female with prior co morbidity of T2DM for which she is on regular medications. She had her index presentation in 2022 in the form of abdominal pain and abdominal discomfort. She was evaluated for the same and was found to have CLD. She also underwent UGIE screening and has been on primary EVL sessions. From last 2 weeks patient complains of on and off blackish motion. She also complaints of a left sided ear pain and Left sided facial weakness that resolved spontaneously. With this clinical picture she has been admitted for further management. There is no h/o fever, jaundice, vomiting, cough, abdominal pain, hematemesis, burning micturition, altered sensorium or decreased urine output. There is no h/o any intoxications, indigenous medications, major surgeries, blood transfusions or IV drug abuse prior to onset of the disease. There are no h/o HTN/CAD/TB/COPD/Thyroid disorders.

**EXAMINATION**

Pt. was conscious, oriented to time place and person.

BP: - 136/70 mm Hg, Pulse: - 86/min, RR: - 14/min and afebrile.

Pallor-, Icterus-, Cyanosis-, Clubbing-, Pedal edema (pitting type) - , LNP-, JVP normal

On systemic examination

Respiratory system:--B/L vesicular breathing, B/L airway equal air entry, no wheeze, no crepts

Cardiovascular system: - S1 S2 normal, no murmurs

CNS: - conscious and oriented with no sensorimotor deficit,

Per Abdomen examination

ON INSPECTION: - moderately distended abdomen, umbilicus central and inverted, skin over the abdomen is stretched with no visible venous prominences, no visible pulsations.

ON PALPATION: - soft, non-tender, liver palpable 1 cm below right coastal margin. Spleen is non- palpable. No guarding, no rigidity, no rebound tenderness.

ON PERCUSSION: - Dull note with free fluid present

ON AUSCULTATION normal bowel sounds present,

**SYSTEMIC REVIEW:**

Patient was admitted with above mentioned complaints. Her examination findings were as mentioned above. Blood Cultures revealed no growth, and Urine Routine revealed 10-12 leukocytes. She was admitted in view of malena and started on aggressive management with RT lavage, fluid resuscitation with IV fluids, IV antibiotics, and other supportive measures. She was started on IV Terlipressin. UGI endoscopy was done to investigate the bleed, which showed Small High risk esophageal varices, mild PHG, ? Duodenal varix - S/P EVL - 12/12/2024. Ascitic fluid tap was done which revealed a straw coloured fluid with WBC - 111, N – 10% and L – 89%, Protein: 0.79, Glu: 151, Albumin: 0.3, SAAG: 2.3, ADA: 2.2. Neurology and ENT consults were done in view of left LMN facial palsy and advice followed. MRI Brain was done – provisionally mild diffuse cerebral atrophy. Final report awaited. Proper diet and mobilization was followed throughout the course of admission. Nutritional optimization has been done and patient is tolerating orally well. The nature of the disease and long term prognosis was explained in great detail to the patient and the patient relatives. Now she is being discharged in hemodynamically stable state with the advice to follow up in OPD.

**ADVICE ON DISCHARGE (Duration of medications if any):**

1600 KCAL/DAY + 70 GRAMS PROTEIN/DAY, LOW SALT (<2GRAMS/DAY), NORMAL DIABETIC DIET AS ADVISED

WEIGHT REDUCTION WITH DAILY 30 MIN EXERCISE

TAB TAXIM-O 200MG BD	1-0-1 X 5 DAYS

TAB METROGYL 400 MG PO TDS X 5 DAYS

TAB FLUCOS 200 MG PO OD FOR 5 DAYS

T. RIFAGUT 550 MG PO BD	1-0-1 (START ON 6TH DAY)

T. ALDACTONE 50MG PO OD 1-0-0 (MONITOR KFT EVERY 5^TH^ DAY)

TAB DYTOR 5 MG PO BD	1-1-0	(MONITOR KFT WEEKLY)

SYP LACTIHEP 30ML PO HS  0-0-1 (ENSURE 2-3 BOWEL MOTIONS/DAY)

SYP SUCRAL 10 ML PO QID X 14 DAYS

TAB PANTOCID 40 MG PO OD BBF X 14 DAYS

TAB DEXAMETHASONE 8 MG PO TDS X -------

TAB VALACYCLOVIR 1 GM PO BD X --------

TAB MIDODRINE 5 MG PO TDS

C. EVION 400MG OD 1-0-0

C. HENZOVIT PO OD 1-0-0

T. ZINCONIA 50MG PO OD 1-0-0

T. ME12 PO OD 1-0-0

NUTRIX ULTRA WHEY POWDER 2 SCOOPS PO BD 1-0-1 (WITH MILK/WATER)

TO CONTINUE SAME MEDICATIONS FOR T2DM AS PRESCRIBED FROM OUTSIDE

REVIEW IN HEPATOLOGY OPD WITH CBC/LFT/KFT/INR REPORTS AFTER 4 WEEKS

PLAN: RESTART BETA BLOCKERS AND DIURETICS ON OPD BASIS

**IN CASE OF FEVER, JAUNDICE, PAIN , BLEEDING / BLACKISH MOTION - REVIEW IN ILBS EMERGENCY SOS.**

### Summary 45

Portal HTN – Non-Bleeder, Grade II Esophageal Varices – Jan 2025

Cirrhosis- Metabolic + ALD (LI – 2024)	(ICD-K70.9)

Decompensated - Ascites / Jaundice / SBP

CTP	11 C	MELD	24

**CURRENT ISSUES:**

- Worsening ascites – SBP (TLC 5047 🡪 166), High SAAG, Low Protein,
- AKI (S. Creat 1.13 🡪 0.73)

**CO MORBIDITY**

- CAD – s/p PCI
- Obesity
- T2DM
- Cholelithiasis

**CHIEF COMPLAINTS –**

- Worsening Abdominal Distension

**INDICATION FOR ADMISSION:**

Evaluation and the management of presenting symptoms

**HISTORY:**

Mr. Patient AS is a 62 year old gentleman with prior co morbidity of AUD. He had is index presentation at when he was incidentally detected to have a chronic liver disease . He had history of abdominal distension for past 1 months and outside evaluated and admitted as patient having ascites and high MELD and advised for LT. There is no h/o fever, vomiting, cough, altered bowel habits, malena, burning micturition, altered sensorium or decreased urine output. There is no h/o any intoxications, indigenous medications, major surgeries, blood transfusions or IV drug abuse prior to onset of the disease. There are no h/o CAD/TB/COPD/Thyroid disorders.

**EXAMINATION**

Pt. was conscious, oriented to time place and person.

BP: 136/70 mmHg, HR- 86/min, RR- 14/min and afebrile.

Pallor-, Icterus-, Cyanosis-, Clubbing-, Pedal edema (pitting type) - , LNP-, JVP normal

On systemic examination

Respiratory system:--B/L vesicular breathing, B/L airway equal air entry, no wheeze, no crepts

Cardiovascular system: - S1 S2 normal, no murmurs

CNS: - conscious and oriented with no sensorimotor deficit,

Per Abdomen examination

ON INSPECTION: Distended abdomen, umbilicus central and inverted, skin over the abdomen is stretched with no visible venous prominences, no visible pulsations.

ON PALPATION: soft, non-tender, liver palpable 1 cm below right coastal margin. Spleen non- palpable below left coastal margin. No guarding, no rigidity, no rebound tenderness.

ON PERCUSSION: Dul note over all quadrants with no free fluid present

ON AUSCULTATION: normal bowel sounds present,

**SYSTEMIC REVIEW:**

Patient was admitted with above mentioned complaints. His examination findings were as mentioned above. Blood and Urine Cultures revealed no growth, and Urine Routine was unremarkable. S.PCT – 4.26. Ascitic fluid tap was done which revealed a straw coloured fluid with WBC - 5047, N – 93% and L – 06%, Protein: 0.67, Glu: 173, Albumin: 0.28, ADA: 2.9 and Gene Xpert: negative, Gram Stain negative. Cultures were negative. Patient was treated with IV antibiotics, IV albumin, nutritional support and other supportive medication and treatment. Antibiotics upgraded in view of SBP. The nature of the disease and long term prognosis was explained in great detail to the patient and the patient relatives and were also explained regarding the need of liver transplant in view of Decompensated Liver Disease. However, no donor is available at present. LT referral done. Response tap revealed resolution of SBP. Proper diet, purging and mobilization was followed throughout the course of admission. All vitals, temperature, RBS and other necessary parameters were checked regularly and managed appropriately. He has recovered symptomatically following the management. He is discharged**,** in hemodynamically stable condition with following advice to follow in OPD.

**PLAN / ADVICE AT DISCHARGE (Including duration of medication if any):**

1800 KCAL/DAY + 80 GRAMS PROTEIN/DAY

INJ IMIPENEM 500 MG IV TDS X 2 DAYS, FOLLOWED BY

TAB FAROPENEM 200 MG PO BD FOR 5 DAYS

TAB NORFLOX 400 MG PO OD (START ON 7TH DAY)

TAB CARDIVAS 3.125 MG PO OD HS (DO NOT GIVE IF HR < 55 OR BP < 90/60 MMHG)

CAP HENZOVIT 1 TAB PO OD

TAB ME-12 1 TAB PO OD

HEPSURE SACHET PO BD

SYP LACTIHEP 30ML PO HS  0-0-1 (ENSURE 2-3 BOWEL MOTIONS/DAY)

PROHANCE HP POWDER 2 SCOOP PO TDS

INJ. ALBUMIN 20% 100 ML IV ONCE A WEEK UNDER MEDICAL SUPERVISION OVER 4-6 HOURS

REVIEW IN HEPATOLOGY OPD AFTER 2 WEEKS WITH CBC/LFT/KFT/INR REPORTS

IN CASE OF DECREASE IN URINE OUTPUT, ALTERED SENSORIUM, BLEEDING, FEVER, NEW ONSET COUGH AND REVIEW IN ILBS EMERGENCY ON SOS BASIS.

### Summary 46

PHTN (Non - Bleeder – Grade I Esophageal Varices, Mild PHG – 14/05/2025)

Acute-on-Chronic Liver Failure - ----- Acute: Hepatitis B Reactivation (HBV DNA – 1.3 Log 6)

Chronic Hepatiits B **(B18.9)**

AARC Score – 6	Grade I	OF – None	OD – None

CTP - 12, CHILD - C, MELD Na - 29

HVPG 16 mmHg, Biopsy – 14/05/2025 – Awaited

**COMORBIDITIES:** None

**CURRENT ISSUES:**

- Reactivation of HBV (DNA –5.5 log 7 🡪 1.3 log 6)

**PRESENTING COMPLAINTS**

Yellowish discoloration of eyes since 15 days

**INDICATION FOR ADMISSION:**

Evaluation and management of symptoms

**HISTORY:**

Mr Patient AT is a 32 year old gentleman, who had his index presentation at an outside centre in 2017 when he had an episode of fever. On routine evaluation, he was discovered to be Hep B positive. He was started on TAF however, there is poor compliance to medications. Over the past two weeks, he first noticed a yellowish discolouration of his eyes and urine, which was insidious in onset and progressive. It was not associated with itching or clay-coloured stools. He was evaluated at an outside centre and revealed to be Hepatitis B positive, with chronic liver disease, and referred to ILBS. There was no h/o altered sensorium or acute bleeds. There is no h/o any intoxications, major surgeries, blood transfusions or IV drug abuse prior to onset of the disease. There is no history of CAM intake. There is no h/o DM/HTN/CAD/TB/COPD/Thyroid disorder.He has significant family history as his father expired due to HCC.

**EXAMINATION**

Pt. was conscious, oriented to time place and person.

BP: - 116/70 mm Hg, Pulse: - 86/min, RR: - 14/min and afebrile.

Pallor-, Icterus+ , Cyanosis-, Clubbing-, Pedal edema (pitting type) + , LNP-, JVP normal

On systemic examination

Respiratory system:--B/L vesicular breathing, B/L airway equal air entry, no wheeze

Cardiovascular system: - S1 S2 normal, no murmurs

CNS: - conscious and oriented with no sensorimotor deficit,

Per Abdomen examination

ON INSPECTION: - scaphoid abdomen, no visible swellings or pulsations.

ON PALPATION: - soft, non-tender. Spleen palpable 3 cm below RCM. No guarding, no rigidity, no rebound tenderness.

ON PERCUSSION: - Tympanic note, free fluid absent

ON AUSCULTATION: - normal bowel sounds present

**SYSTEMATIC REVIEW:**

Patient was admitted with above mentioned complaints and baseline investigations were sent. His examination findings were as mentioned above. His initial lab data and latest lab data is included at the end of the summary. Anti – HAV IgM – NR, Anti – HEV IgM – NR; Anti HBc total – Reactive, Anti HBe Non-Reactive; HBV DNA - 1.4 log 5; HBsAg – Reactive. CECT W/A was done - Findings consistent with early CLD. Segment II hepatic lesion with up to cm size abdominal nodes - ? small abcess. He was started on iv fluids, iv antibiotics and other supportive treatment. HVPG and Liver biopsy (TJLB) was done on 14/05/2025. HVPG – 16 mmHg. Biopsy is awaited. UGIE for Variceal screening revealed Grade I Esophageal Varices, Mild PHG on 14/05/2025. The nature of the disease and long term prognosis was explained in great detail to the patient and the patient relatives. Proper diet and mobilisation was followed throughout the course of admission. All vitals, temperature, RBS and other necessary parameters were checked regularly and managed appropriately. He is discharged in hemodynamically stable condition with advice to follow up in OPD.

**PLAN / ADVICE AT DISCHARGE (Including duration of medication if any):**

2000 KCAL/DAY + 90 GRAMS PROTEIN/DAY

TAB TAXIM-O 200MG BD	1-0-1 X 5 DAYS

TAB THIOTRESS 500 MG PO BD X 10 DAYS

TAB TAF 25 MG PO OD

TAB ENTACAVIR 0.5 MG PO OD

TAB URSOCOL 450 MG PO BD

SYP LACTIHEP 15 ML PO HS (ENSURE 2-3 MOTIONS PER DAY)

CAP HENZOVIT 1 TAB PO OD

SITZ BATH TDS

REVIEW IN VIRTUAL OPD AFTER 4 DAYS WITH CBC/LFT/KFT/INR.

REVIEW IN HEPATOLOGY OPD AFTER 1 WEEK.

**PLAN: PLEX IF NO IMPROVEMENT IN JAUNDICE**

IN CASE OF DECREASE IN URINE OUTPUT, ALTERED SENSORIUM, BLEEDING, FEVER, NEW ONSET COUGH AND REVIEW IN ILBS EMERGENCY ON SOS BASIS.

### Summary 47

Antral Gastritis

**CURRENT ISSUES:**

Chronic Epigastric Pain AND Weight Loss U/E

- UGIE – 16/10/2024 – Antral Gastritis, RUT positive, D2 Biopsy – Normal Villous Architecture, Chronic Gastritis
- Tumour Markers, Amylase CECT grossly normal

**PRESENTING COMPLAINTS:**

Chronic pain abdomen x 1-1.5 years – on and off

Dyspepsia x 2 weeks

Loss of weight x 2 months

**INDICATION FOR ADMISSION:**

Evaluation and the management of the symptoms

**HISTORY:**

Mrs. Patient AU is a 66 year old lady with index presentation of abdominal pain – on and off since 2 years. Over the past two weeks, she has complained of a burning retrosternal pain with associated nausea. There is no aggravation of pain on exertion or radiation of pain. Pain worsens at night. She presented for further evaluation of the same. There is associated weight loss of 6 kg over the past month. There is no h/o jaundice, vomiting, cough, abdominal pain, altered bowel habits, hematemesis, and malena, burning micturition, altered sensorium or decreased urine output. There is no h/o any intoxications, indigenous medications, blood transfusions or IV drug abuse prior to onset of the disease.

**EXAMINATION**

Pt. was conscious, oriented to time place and person.

BP: - 126/70 mm Hg, Pulse: - 86/min, RR: - 14/min and afebrile.

Pallor -, Icterus - , Cyanosis - , Clubbing - , Pedal edema- , LNP-, JVP normal

On systemic examination

Respiratory system:--B/L vesicular breathing, B/L airway equal air entry, no wheeze,

Cardiovascular system: - S1 S2 normal, no murmurs

CNS: - conscious and oriented with no sensorimotor deficit,

Per Abdomen examination

ON INSPECTION: - abdomen not distended, umbilicus central and inverted, no visible venous prominences, no visible pulsations.

ON PALPATION: - soft, non-tender, no organomegaly. No guarding, no rigidity, no rebound tenderness.

ON PERCUSSION: - Tympanic note with no fluid present

ON AUSCULTATION: - Normal bowel sounds present

**SYSTEMATIC REVIEW:**

Patient was admitted with above mentioned complaints. Her examination findings were as mentioned above. Her initial lab data and latest lab data is included at last of summary. Tumour markers were within normal limits. CECT done on 15/10/2024 – reported normal study. MRCP was done on 17/10/2024 – provisionally normal. Final report awaited. She was started on IV fluids and IV antibiotics. She underwent a UGI Endoscopy on 15/10/2024. UGIE revealed an antral gastritis with RUT positive, and D2 Biopsy – Normal Villous Architecture, Chronic Gastritis. Patient was treated with nutritional support and other supportive medication and treatment. She had an episode of acute urinary retention and was catheterized for the same. Urology consult done and advice followed. All vitals, temperature, RBS and other necessary parameters were checked regularly and managed appropriately. She recovered symptomatically following the management and is discharged in hemodynamically stable condition with the advice to follow up in OPD.

**PLAN / ADVICE AT DISCHARGE (Including duration of medication if any):**

2100 Kcal/day + 90 grams protein/day

TAB TAXIM-O PO BD X 5 DAYS

PANTOCID DSR 1 TAB PO OD BBF X 7 DAYS

TAB DROTIN 1 TAB PO SOS

SYP MUCAINE GEL 2 TSP PO TDS

TAB TRYPTOMER 25 MG PO HS

TAB URIMAX 0.4 MG PO HS

CAP HEPAGRESS PO BD 1-0-1 X 5 DAYS

SYP LACTIFIBRE 30ML PO HS  0-0-1 FOR CONSTIPATION

DIETARY AND LIFESTYLE CHANGES AS ADVISED

PLAN: PET-CT AFTER 1 MONTH IF NO IMPROVEMENT

FOLLOW UP AFTER 2 WEEKS IN HEPATOLOGY OPD

IN CASE OF DECREASE IN URINE OUTPUT, ALTERED SENSORIUM, BLEEDING, FEVER, NEW ONSET COUGH AND REVIEW IN ILBS EMERGENCY ON SOS BASIS.

### Summary 48

PHTN - (Non Bleeder, Large High Risk Esophageal Varices s/p EVL, Large GOV 1 s/p Glue - 23/10/2024)

CLD - NASH related (ICD10 – K75.8)

Compensated

Multifocal Metastatic HCC (ICD - C22.0)

- 9.6 x 7.0 cm lesion - segments VII and VIII
- Few other mildly FDG avid arterial phase hyperenhancing lesions with portovenous washout noted in segments VII, VIII and III of liver
- No PVT

Planned for SBRT on OPD basis

s/p HDVK

PS – 0	BCLC - B

**Comorbidities –** HTN (controlled)

**CURRENT ISSUES –** Multifocal Metastatic HCC - s/p SBRT – 24/10/2024

**CHIEF COMPLAINTS -**

Generalized Weakness

**INDICATION FOR ADMISSION:**

Evaluation and management of the symptoms.

**HISTORY:**

Mr. Patient AV is a 66 year old gentleman who had his index presentation in 2020 with complaints of abdominal pain. He was diagnosed as fatty liver and was on regular follow up. He then developed an abdominal distension which was painless, progressive accompanied by weight gain. He was evaluated and diagnosed to have CLD - ethanol related and HCC. He has now has presented for management of the same. There is no h/o jaundice, fever, vomiting, cough, altered bowel habits, malena, burning micturition, altered sensorium or decreased urine output. There is no h/o any intoxications, indigenous medications, major surgeries, blood transfusions or IV drug abuse prior to onset of the disease. There is no h/o TB/COPD.

**EXAMINATION**

Pt. was conscious, oriented to time place and person.

BP: - 105/70 mm Hg, Pulse: - 86/min, RR: - 14/min and afebrile.

Pallor-, Icterus-, Cyanosis-, Clubbing-, Pedal edema (pitting type) -, LNP-, JVP normal

On systemic examination

Respiratory system: B/L vesicular breathing, B/L airway equal air entry, no wheeze, no crepts

Cardiovascular system: - S1 S2 normal, no murmurs

CNS: - conscious and oriented with no sensorimotor deficit,

Per Abdomen examination

ON INSPECTION: - globoid abdomen, umbilicus central and inverted, skin over the abdomen is normal with no visible venous prominences, no visible pulsations.

ON PALPATION: - soft, non-tender, liver non- palpable. Spleen non- palpable. No guarding, no rigidity, no rebound tenderness.

ON PERCUSSION: - tympanic note with free fluid absent

ON AUSCULTATION: - normal bowel sounds present,

**SYSTEMIC REVIEW**

Patient was admitted with above mentioned complaints. His examination findings were as mentioned above. Patient has been counselled regarding limited treatment options and therefore advised for immunotherapy. AFP – 2.8. PIVKA II – 1.2 Log 5. ELF Score – 12.67. PET – CT – 22/10/2024

- Features suggestive of chronic liver disease with portal hypertension, ascites and splenomegaly as described.
- A large lobulated heterogeneous density mass lesion measuring 9.6 x 7.0 cm noted involving segments VII and VIII of liver with heterogeneously increased FDG uptake (SUVmax- 9.7). The mass shows areas of arterial phase hyperenhancement showing portovenous washout.
- Few other mildly FDG avid arterial phase hyperenhancing lesions with portovenous washout noted in segments VII, VIII and III of liver (largest measuring 3.5 x 3.4 cm; SUVmax- 6.4)
- Bilateral pleural effusion with few mild metabolically active mediastinal lymph nodes-infective/inflammatory.
- No other hypermetabolic lesion noted in rest of the body.

Radiation Oncology referral done and patient was planned for SBRT. Nutritional optimization has been done and patient is tolerating orally well. Now he is being discharged in haemodynamically stable state with the advice to follow up in OPD. The advanced nature of the disease and its long term prognosis and limited treatment options was explained in great detail to the patient and the patients attendant.

**ADVICE ON DISCHARGE (Duration of medications if any):**

1800 Kcal/day + 80 grams protein/day, low salt (<2grams/day)

TAB TAXIM-O 200MG BD 1-0-1 FOR 5DAYS

T. RIFAGUT 550 MG PO BD	1-0-1

TAB CARDIVAS PO BD – AM – 6.250 MG (2 TABS) PM – 9.375 MG (3 TABS)

(DO NOT GIVE IF HR < 55 OR BP < 90/60 MMHG)

TAB TELMISARTAN + CHLORTHALIDONE PO OD

TAB THIOTRESS 500 MG PO BD X 10 DAYS

TAB UDCA 450 MG PO BD

Review in Hepatology OPD with CBC/LFT/KFT/INR reports after 2 weeks.

**PLAN: UGIE + ENDOTHERAPY AFTER 3 WEEKS, TKI AFTER SBRT**

### Summary 49

**DISCHARGE ON REQUEST**

PHTN (Non-Bleeder, Small High Risk Oesophageal Varices, Mild PHG – 29/01/2025)

Acute-on-Chronic Liver Failure syndrome---- Severe Alcoholic Hepatitis, mDF – 35 (ICD - K72.1)

Chronic: Alcoholic Liver Disease (Last Intake – 10 days ago)

**On Steroid Therapy started on 30/01/2025**

Planned for close follow up, as patient is being discharged on request

CTP - 10	CHILD – C	MELD Na - 25

HVPG 12 mmHg,

Liver biopsy (TJLB) – report awaited – 28/01/2025

**CURRENT ISSUES**: Severe Alcoholic Hepatitis mDF – 35

**COMORBIDITIES:** None

**PRESENTING COMPLAINTS**

Yellowish discoloration of eyes since 30 days

**INDICATION FOR ADMISSION:**

Evaluation and management of symptoms

**HISTORY:**

Mr Patient AW is a 33 years old male, who had presented with a history of jaundice. He first noticed a yellowish discolouration of his eyes and urine since 30 days, which was insidious in onset and progressive. It was not associated with itching or clay-coloured stools. However, he complains of a pedal edema since 1 week. He is a known ethanol user – with last intake 10 days ago. There is no h/o vomiting, cough, abdominal pain, altered bowel habits, hematemesis, and malena, burning micturition, altered sensorium or decreased urine output. There is no history of loss of weight or appetite. There is no history of CAM intake. There is no h/o any major surgeries, blood transfusions or IV drug abuse prior to onset of the disease. There is no h/o CAD/COPD/Thyroid disorder.

**EXAMINATION**

Pt. was conscious, oriented to time place and person.

BP: - 116/70 mm Hg, Pulse: - 86/min, RR: - 14/min and afebrile.

Pallor-, Icterus++, Cyanosis-, Clubbing-, Pedal edema (pitting type) + , LNP-, JVP normal

On systemic examination

Respiratory system:--B/L vesicular breathing, B/L airway equal air entry, no wheeze

Cardiovascular system: - S1 S2 normal, no murmurs

CNS: - conscious and oriented with no sensorimotor deficit,

Per Abdomen examination

ON INSPECTION: - abdomen scaphoid, umbilicus central and inverted, no visible swellings or pulsations.

ON PALPATION: - soft, non-tender, Liver palpable 2 cm below right costal margin. Spleen not palpable. No guarding, no rigidity, no rebound tenderness.

ON PERCUSSION: - Tympanic note, free fluid absent

ON AUSCULTATION normal bowel sounds present

**SYSTEMATIC REVIEW:**

Patient was admitted with above mentioned complaints and baseline investigations were sent. His examination findings were as mentioned above. His initial lab data and latest lab data is included at the end of the summary. Blood C/S showed no growth, and Urine RME was unremarkable. Serum PCT – 0.63. He underwent HVPG + TJLB on 28/01/2025 - HVPG 12 mmHg, Liver biopsy (TJLB) – awaited. UGIE for Variceal screening revealed Small High Risk Oesophageal Varices. CECT W/A was done - revealed no SOL, no PVT - provisional report. Final report awaited. In view of severe alcoholic hepatitis, after explaining benefits and cons, a steroid course was opted for and initiated on 30/01/2025. The procedures were tolerated well. The nature of the disease and long term prognosis was explained in great detail to the patient and the patient relatives. Proper diet and mobilisation was followed throughout the course of admission. All vitals, temperature, RBS and other necessary parameters were checked regularly and managed appropriately. Strict alcohol abstinence advised. He recovered symptomatically following the management and is **discharged on request** in hemodynamically stable condition with following advice to follow in OPD. The precautions to be taken, need for very close follow up has been explained.

**PLAN / ADVICE AT DISCHARGE (Including duration of medication if any):**

2200 KCAL/DAY + 90 GRAMS PROTEIN/DAY, LOW SALT (<2GRAMS/DAY)

**PERSONAL PROTECTION AND HYGIENE AS ADVISED.**

**AVOID SOCIAL GATHERINGS, WEAR MASK REGULARLY.**

**STRICT ALCOHOL ABSTINENCE**

TAB TAXIM-O 200MG BD	1-0-1 X 5 DAYS

TAB WYSOLONE 40 MG PO OD, 1-0-0 AT 10 AM FOR TOTAL 28 DAYS, AND THEN STOP.

TAB CARDIVAS 3.125 MG PO BD (DO NOT GIVE IF HR < 55 OR BP < 90/60 MMHG)

CAP ALCOMAX 300 MG PO BD

TAB UDCA 450 MG PO BD

HEPSURE SACHET PO BD

SYP LACTIHEP 30ML PO HS  0-0-1 (ENSURE 2-3 BOWEL MOTIONS/DAY)

LAXOPEG SACHET BD. 1-0-1 (ENSURE 2-3 BOWEL MOTIONS/DAY)

TAB ME-12 1 TAB PO OD

CAP HENZOVIT 1 TAB PO OD

TAB SHELCAL 1 TAB PO OD

CAP LUMIA 60,000 IU WEEKLY

TAB ZINCONIA 50 MG PO OD

REVIEW IN HEPATOLOGY OPD ON MONDAY – 06/01/2025 WITH CBC / LFT / FBS/PPBS charting of 4 days/INR / KFT REPORTS

IN CASE OF DECREASE IN URINE OUTPUT, ALTERED SENSORIUM, BLEEDING, FEVER, NEW ONSET COUGH AND REVIEW IN ILBS EMERGENCY ON SOS BASIS.

### Summary 50

Portal Hypertension - (Non-Bleeder, Small Low Risk Esophageal Varices with Mild PHG – 11/03/2025, FMT done)

Cirrhosis of Liver - Ethanol (Last Intake: 1 Day prior to admission)

Decompensated - Hepatic Encephalopathy / Ascites

FMT single session done on 11/03/2025

MELD Na: 18	CTP: 10 C      **ICD-K70.9**

**CURRENT ISSUES:**

- Alcoholic Hepatitis (mDF - 45)
- AUD with Strong Recidivism – **s/p FMT done on 11/03/2025**
- Alcohol Withdrawal Syndrome
- Bleeding P/R – s/p Sigmoidoscopy – External Haemorrhoids

**CHIEF COMPLAINTS:**

- Bleeding P/R x 1 day

**INDICATION FOR ADMISSION :**

Evaluation and the management of the symptoms

**HISTORY :**

Mr Patient AX, 40 year old male with TUD and AUD, had his index presentation in 2023 in the form of altered sensorium. He was evaluated and was found to have liver disease. He has strong recidivism. He presented to ILBS with the compalinst of generalized weakness and bleeding per rectum x 2 days. There is no h/o fever, jaundice, vomiting, cough, abdominal pain, altered bowel habits, hematemesis, burning micturition, or decreased urine output. There is no h/o any indigenous medications, major surgeries or IV drug abuse prior to onset of the disease. There is no h/o DM/HTN/CAD/TB/COPD/Thyroid disorders.

**EXAMINATION :**

Pt. was conscious, oriented to time place and person.

BP: -140/82 mm Hg, Pulse: -88 /min, RR: - 22/min and afebrile.

Pallor-, Icterus-, Cyanosis-, Clubbing-, Pedal edema (pitting type) - , LNP-, JVP normal

On systemic examination

Respiratory system:--B/L vesicular breathing, B/L airway equal air entry, no wheeze, no crepts

Cardiovascular system: - S1 S2 normal, no murmurs

CNS: - Altered sensorium

Per Abdomen examination

ON INSPECTION: - Scaphoid abdomen, umbilicus central and inverted, skin over the abdomen is stretched with no visible venous prominences, no visible pulsations.

ON PALPATION: - Soft, non-tender, liver non- palpable, Spleen non- palpable. No guarding, no rigidity, no rebound tenderness.

ON PERCUSSION: - Tympanic note with free fluid absent

ON AUSCULTATION: Normal bowel sounds present,

**SYSTEMIC REVIEW:**

Patient was admitted with above mentioned complaints. His examination findings were as mentioned above. His initial lab data and latest lab data is included at last of summary. Psychiatry opinion was taken in view of AUD and AWS and their advice was followed and subsequently his improved symptomatically. Patient was treated with IV antibiotics, nutritional support and other supportive medication and treatment. UGIE was done in view of suspected UGI Bleed - Small Low Risk Esophageal Varices with Mild PHG. Sigmoidoscopy – External haemorrhoids. He was counselled and planned for FMT and risks and benefits for the same was explained. UGIE + D2 biopsy + FMT was done on 11/03/2025 – Small low risk esophageal varices, mild PHG, FMT administered. Strict alcohol abstinence advised. The nature of the disease and long term prognosis was explained in great detail to the patient and the patient relatives. He is now planned for nutritional optimization and improvement in performance status. Proper diet, purging and mobilisation was followed throughout the course of admission. All vitals, temperature, RBS and other necessary parameters were checked regularly and managed appropriately. He recovered symptomatically following the management and is discharged in hemodynamically stable condition with following advice to follow in OPD.

**ADVICE ON DISCHARGE (Duration of medications if any):**

2500 KCAL/DAY + 80 GRAMS PROTEIN/DAY, LOW SALT (<2GRAMS/DAY), NORMAL DIET

TAB CARDIVAS 3.125 MG PO BD (DO NOT GIVE IF HR < 55 OR BP < 90/60 MMHG)

TAB LASILACTONE ½ TABLET (20/50) PO OD (MONITOR KFT WEEKLY)

TAB LEVEPIL 500 MG PO BD

TAB ZOLFRESH 5 MG PO OD HS / SOS

TAB LIBRIUM 10 MG PO TDS X 5 DAYS

LAXOPEG SACHET BD. 1-0-1 (ENSURE 2-3 BOWEL MOTIONS/DAY)

LORHEP SACHET PO BD

TAB SITCOM FORTE PO BD

ANOVATE CREAM L/A

BRANVIO SACHET PO BD

CAP ALCOMAX 300 MG PO BD

TAB HEPKART 400 MG PO DO

CAP HENZOVIT 1 TAB PO OD

TAB ME-12 1 TAB PO OD

SYP ZINCONIA 10 ML PO BD

REVIEW IN HEPATOLOGY OPD WITH CBC/LFT/KFT/INR REPORTS AFTER 2 WEEKS.

PSYCHIATRY OPD FOLLOW UP AS ADVISED

ER SOS
