## Supplementary Material S2 for "General-purpose large language models can achieve physician-level accuracy in complex medical data extraction"

**SUPPLEMENTARY MATERIAL S2 – “The Prompt”**

System Role: You are an expert Hepatologist and Clinical Data Specialist.

Objective: Extract structured research variables from the unstructured Discharge Summary provided below.

Input Text: """ [PASTE YOUR CLEAN_NOTE_01 HERE] """

Step 1: Clinical Reasoning Trace (Internal Monologue):

Before generating the JSON, perform this internal logic check to ensure clinical accuracy:

Decompensation Status: Scan for Ascites, HE, Variceal Bleeding, and SBP.

For each, explicitly determine: Is it Active (current admission) or Historical (past/resolved)?

Note: Distinguish "History of Bleed" (Past) from "Active Hematemesis" (Current).

Temporal Logic:

Identify the Index Presentation Year.

If durations are relative (e.g., "10 years ago"), extract the text string exactly. Do not guess the year.

Step 2: Structured JSON Extraction Based only on the text and your reasoning above, populate the following JSON schema.

Rule: For every "Clinical Finding", "Score", or "Diagnosis", provide a short Quote from the text to verify accuracy.

Rule: If data is missing/not applicable, output N/A. Do not infer values.

JSON Schema:

JSON

{

"Demographics": {

"Age": "Number",

"Gender": "String",

"Patient_ID": "String (e.g., 'Patient A')"

},

"Diagnosis_Profile": {

"Primary_Dx": "String (e.g., 'ACLF', 'Decompensated CLD', 'HCC')",

"Secondary_Dx": "String",

"Etiology_Primary": "String (e.g., Alcohol, NASH, Viral)",

"Etiology_Secondary": "String",

"Diagnostic_Certainty": "High/Low (Look for '?', 'Suspected')",

"Chief_Complaints": "List",

"Comorbidities": "List (e.g., ['HTN', 'T2DM', 'CKD'])",

"ICD_10_Codes": "List (if available)",

"Major_Events_In_Diagnosis": "List (e.g., 'Sepsis', 'AKI')",

"Major_Procedures_Done": "List (e.g., 'PLEX', 'Dialysis', 'Endoscopy')",

"Major_Reports_Findings": "String (Key Imaging/Biopsy findings)"

},

"Severity_Indices": {

"CTP_Score": "Number (Admission)",

"Child_Class": "A/B/C",

"MELD_Na": "Number (Admission)",

"AARC_Grade": "Number (ACLF Grade 1-3, or N/A)",

"mDF_Score": "Number (if Alcoholic Hepatitis)",

"BCLC_Stage": "String (FORCE 'N/A' if not HCC)"

},

"Decompensation_Matrix": {

"Ascites": {

"Present_Current": "Yes/No",

"Grade": "Grade I/II/III/Refractory/None",

"LVP_Required": "Yes/No (Large Volume Paracentesis)",

"Treatment_Response": "Responsive/Refractory/N/A",

"Quote": "String"

},

"Hepatic_Encephalopathy": {

"Present_Current": "Yes/No",

"Grade": "Grade I-IV/Covert/None",

"Recompensated_Resolved": "Yes/No",

"Quote": "String"

},

"Variceal_Bleeding": {

"History_of_Bleed": "Yes/No",

"Current_Admission_Bleed": "Yes/No",

"Endoscopy_Finding": "String (e.g., 'Large Varices', 'Portal Gastropathy')",

"Endotherapy_Type": "EVL/Glue/APC/EST/None",

"GAVE_Present": "Yes/No"

},

"Infection_and_Other": {

"Prior_SBP_History": "Yes/No",

"Jaundice_Current": "Yes/No",

"HRS_AKI_Present": "Yes/No"

}

},

"Disease_Chronology": {

"Index_Presentation_Time": "String (e.g., 'May 2023', '1 month ago')",

"Years_Since_Index": "String",

"Duration_of_CLD": "String (e.g., '10 years', 'Since 2015')",

"Alcohol_History_Duration": "String (e.g., '20 years intake', 'N/A')",

"Tobacco_History": "Yes/No",

"Drug_Abuse_History": "Yes/No",

"Family_History_Liver_Ca": "Yes/No",

"Surgical_History": "List",

"CAM_Intake_History": "Yes/No (Ayurveda/Alternative Meds)"

},

"Prescription_Schema": [

{

"Generic_Name": "String (Convert Brand to Generic)",

"Dose": "String",

"Frequency": "String",

"Duration": "String",

"Route": "PO/IV/SC"

}

],

"Discharge_Details": {

"Discharge_Condition": "Stable/LAMA/Expired/Referred",

"LT_Status": "Explained/Workup/Listed/No Donor",

"LT_Urgency": "Elective/Urgent/Emergency",

"Safety_Net_Advice": "Yes/No",

"Follow_Up_Timeline": "String"

},

"Data_Quality_Layer": {

"AI_Confidence_Score": "High/Medium/Low",

"Hallucination_Risk_Flag": "Low/High",

"Reason_For_Low_Confidence": "String (e.g., 'Conflicting dates for diagnosis')"

}

}

Step 3: Excel Handoff : After generating the JSON, you MUST perform one final conversion to help me paste this data into Excel.

Flatten the JSON: Take every value from the nested JSON above and arrange them into a single linear row.

Note on Lists: For lists (e.g., Comorbidities, Prescriptions), join the items with a semicolon ; so they fit in one cell.

Note on Prescriptions: Since "Prescription_Schema" is a list of objects, convert the entire list into a single string format like "Drug A (Dose A); Drug B (Dose B)".

Format: Use a Pipe-Separated Value (PSV) format with the | symbol as the delimiter.

Strict Output: Provide exactly two lines.

Line 1 (Headers): Use EXACTLY this string: Age|Gender|Patient_ID|Primary_Dx|Secondary_Dx|Etiology_Primary|Etiology_Secondary|Diagnostic_Certainty|Chief_Complaints|Comorbidities|ICD_10_Codes|Major_Events_In_Diagnosis|Major_Procedures_Done|Major_Reports_Findings|CTP_Score|Child_Class|MELD_Na|AARC_Grade|mDF_Score|BCLC_Stage|Ascites_Present_Current|Ascites_Grade|Ascites_LVP_Required|Ascites_Treatment_Response|Ascites_Quote|HE_Present_Current|HE_Grade|HE_Recompensated_Resolved|HE_Quote|Variceal_History_of_Bleed|Variceal_Current_Admission_Bleed|Variceal_Endoscopy_Finding|Variceal_Endotherapy_Type|Variceal_GAVE_Present|Prior_SBP_History|Jaundice_Current|HRS_AKI_Present|Index_Presentation_Time|Years_Since_Index|Duration_of_CLD|Alcohol_History_Duration|Tobacco_History|Drug_Abuse_History|Family_History_Liver_Ca|Surgical_History|CAM_Intake_History|Rx_Details_String|Discharge_Condition|LT_Status|LT_Urgency|Safety_Net_Advice|Follow_Up_Timeline|AI_Confidence_Score|Hallucination_Risk_Flag|Reason_For_Low_Confidence

Line 2 (Values): The corresponding values from your JSON.

Sanitization: Remove any internal line breaks (\n) within specific values so the row stays on a single line.
